# Modelling timelines to elimination of sleeping sickness in the DRC accounting for possible cryptic human and animal transmission

**DOI:** 10.1101/2024.03.19.24304554

**Authors:** Ronald E Crump, Maryam Aliee, Samuel A Sutherland, Ching-I Huang, Emily H Crowley, Simon E F Spencer, Matt J Keeling, Chansy Shampa, Erick Mwamba Miaka, Kat S Rock

**Author notes:** Equal contributor (Ronald E Crump, Maryam Aliee).

## Abstract

Sleeping sickness (gambiense human African trypanosomiasis, gHAT) is a vector-borne disease targeted for global elimination of transmission (EoT) by 2030. There are, however, unknowns that have the potential to hinder the achievement and measurement of this goal. These include asymptomatic gHAT infections (inclusive of the potential to self-cure or harbour skin-only infections) and whether gHAT infection in animals can contribute to the transmission cycle in humans. Using modelling we explore how cryptic (undetected) transmission impacts the monitoring of progress towards as well as the achievement of the EoT goal. We have developed gHAT models that include either asymptomatic or animal transmission, and compare these to a baseline gHAT model without either of these transmission routes, to explore the potential role of cryptic infections on the EoT goal. Each model was independently calibrated using available historic human case data for 2000––2020 (obtained from the World Health Organization’s HAT Atlas) which includes routine data from active and passive screening for five different health zones in the Democratic Republic of the Congo (DRC).

Our results suggest that when matched to past case data, we estimated similar numbers of new human infections between model variants, although human infections were slightly higher in the models with cryptic infections. We simulated the continuation of screen-confirm-and-treat interventions and found that forward projections from the animal and asymptomatic transmission models produced lower probabilities of EoT than the baseline model. Simulation of a (as yet to be available) screen-and-treat strategy found that removing a parasitological confirmation step was predicted to have a more noticeable benefit to transmission reduction under the asymptomatic model compared to the others. Our simulations suggest vector control could greatly impact all transmission routes in all models, although this resource-intensive intervention should be carefully prioritised.

## Introduction

*Gambiense* human African trypanosomiasis (gHAT), commonly referred to as “sleeping sickness”, is a neglected tropical disease that threatens the lives of millions of the poorest populations in West and Central Africa. The disease, which is caused by the parasite *Trypanosoma brucei gambiense*, is transmitted to humans through the bite of tsetse (*Glossina*) [1, 2] and once symptoms develop the untreated disease is usually fatal. The last gHAT epidemic, which endured from the 1970s until the late 1990s, reached its peak in 1998 with 37,385 cases reported across Africa [3]. In response to this epidemic the World Health Organization (WHO), national control programmes and nongovernmental organizations (NGOs) implemented a range of interventions in endemic areas which led to a significant reduction in case numbers [3, 4]. Two approaches were adopted to control the disease: case detection through passive screening (fixed health facilities for patients presenting with gHAT symptoms) and active screening (AS; predominantly mobile teams travelling to at-risk regions and screening using the card agglutination test for trypanosomiasis (CATT) or rapid diagnostic tests (RDTs)) [5]. Additionally, vector control (VC; reduction of tsetse populations in endemic areas) has been used in some settings including geographically contained foci and some of the regions with highest case reporting. The success of these global and national efforts led to fewer than 1000 annual cases being reported in 2019–2022 [3, 6].

In 2012, following a decade of sustained control efforts, the WHO included gHAT in its “roadmap for eradication, elimination and control of neglected tropical diseases” [7]. The goals identified were for the elimination of gHAT as a public health problem by 2020 and zero transmission in humans by 2030. Steady progress has been made towards these goals with Togo and Ĉote d’Ivoire being the first to have elimination as a public health problem validated, followed by Benin, Uganda and Rwanda and other countries are working towards building and submitting dossiers [8]. Even with falling case numbers and the sustained progress made by national programmes and their partners towards achieving the WHO goals, uncertainties remain. Undetected or cryptic hosts, including asymptomatic gHAT infections and animal infections, represent a significant uncertainty, and understanding the role they play in maintaining the human transmission cycle will be critical for elimination efforts [9].

Although the classical gHAT disease course is typically characterised by an early and a late stage [10] it is now clear that there is a range of potential clinical outcomes with some infected individuals displaying no symptoms following infection (asymptomatic) and some able to clear the parasite spontaneously (self-cure) [11]. Healthy carriers of gHAT have been documented for half a century and can remain infected for years, possibly even decades [11, 12]. The parasite, in such cases, can evade detection by parasitological tests used for routine screening of blood, lymph node aspirate or cerebral spinal fluid, by residing in the extravascular space of organs including the heart and the skin [13, 14, 15]. Consequently, these individuals may act as a human maintenance reservoir or at least hinder intervention efforts (although it has yet to be established how infective asymptomatic humans are to tsetse [16]). An indicator of asymptomatic infection is consistently high titres in the CATT [17] used for mass screening in endemic locations. The CATT test, however, may not be sufficiently specific to *T.b. gambiense* infections[18]. Other tests that are more specific to *T.b. gambiense* infections, including immune trypanolysis, may more accurately correlate with infection prevalence, however, these tests are laboratory rather than field tests and therefore they are not routinely used for population screening [18, 19]. Consequently, under the standard “screen-confirm-and-treat” AS algorithm, not all individuals infected with gHAT have a chance of being diagnosed and treated due to the parasite confirmation criterion for the currently available drugs: fexinidazole, pentamidine and nifurtimox-eflornithine combination therapy (NECT) [20]. A “screen-and-treat” (S&T) scenario in which all individuals with a positive screening test (CATT or RDT) may be a feasible option, particularly in light of the ongoing progress in the development of a single-dose oral cure, acoziborole [21].

In addition, and despite an abundance of evidence that *T.b. gambiense* parasites are present in both wildlife and domestic livestock, it is uncertain if and to what extent they contribute to the transmission cycle [9]. It has been suggested by several studies that animals can act as parasite reservoirs [22, 23, 24, 25, 26, 27] however, studying the infectivity and transmissibility to humans is challenging. Mathematical modelling has been used to great effect to predict what impact non-human animal infections may have on transmission and the effectiveness of control measures. In one such study it was shown that although the probability of elimination would be expected to remain high in the presence of animal transmission (at least 77% probability of gHAT elimination as a public health problem in Boffa East, Guinea by 2020), intervention strategies would need to remain in place even after elimination as a public health problem to prevent recrudescence [28]. Another study presented a model of heterogeneous exposure of humans to tsetse with animal populations that differed in their ability to transmit infections [29]; it concluded that increasing the intensity of VC was more likely to eliminate transmission while increasing the intensity of human screening reduced the time to elimination. The latter model, however, was not fitted to gHAT case data. Another cautionary study, that used human and animal case data to quantify how different species and groups of species (domestic and wild animals) contribute to transmission dynamics, indicated that independent transmission cycles are likely in wild animals [30] and that interventions targeting humans alone are likely insufficient for elimination of gHAT. Previous work by Crump *et al.* [31] directly compared whether there was statistical support for a model with animal infection, rather than solely anthroponotic transmission and concluded that there are some health zones (administrative regions of around 150,000 people) of the Democratic Republic of Congo (DRC) where human case data indicated there may be some evidence of this. However, the amount of transmission from animals would not be sufficient to maintain infection in the long term without human transmission. Some health zones had more than a 10% difference in the probability of elimination by 2030 between the model predictions made with and without animal transmission. Some other modelling studies have remained inconclusive on the existence of animal transmission having a substantial contribution to human-tsetse infection cycles, however, optimistically, falling case numbers in many regions have indicated that if they do exist then we might expect negligible to minor delays to EoT [32, 33].

In this study, we explore three alternative model variants which describe the transmission of gHAT in the DRC, which currently has the highest global case burden and also great geographic heterogeneity in case reporting [3]. We ask the following questions for model variants with asymptomatic human transmission and animal transmission: (1) is there statistical evidence – based on routinely collected active and passive case data – to support using a model with asymptomatic human infection, and (2) would we expect predictions of case reporting and elimination to be modified using such a model? Whilst other modelling studies have presented [34, 35] and even fitted [36, 37] models with either self-curing human infections or skin-only infections, none have been conclusive on whether there is more statistical evidence for the use of this more complex model variant or if simpler models without these additions can explain the data as well.

## Methods

### Model variants

In this study, we utilised three previously developed variants of the Warwick gHAT model that can be visualised via the schematic diagram shown in Figure 1. Briefly, the baseline model (known in our previous publication as “Model 4” [38, 32, 39, 33]) is a solely anthroponotic human-tsetse model that includes heterogeneity in people’s exposure to tsetse bites. This model, in which low-risk people may attend AS but high-risk people do not participate, was found to match longitudinal trends in the data well and outperformed other simpler anthroponotic variants (referred to as Models 1, 2 and 3 as detailed in previous work [38, 32, 33]). The model with possible animal transmission (“Model 7” in previous publications) is the same as the baseline model but includes the addition of non-human animal transmission to and from tsetse [31]. This model has two extra parameters – the relative density of animals capable of acquiring and transmitting infection compared to the human population size, *k_A_*, and the probability that a tsetse takes a blood meal on this animal population, *f_A_*. The model with animal transmission has been found to have similar statistical evidence to the baseline model when matched to human case data in various regions [38, 32, 31]. Finally, the asymptomatic model with possible self-curing human infections (“Model 9”) is very similar to the baseline model with a high-low-risk structure but with alternative disease progressions (natural history) possible in humans [34]. In particular, upon passing through the intrinsic incubation period (*E_H_* ), individuals have a probability (*p_bs_*) of developing stage 1 blood infection (*I^b^*_1*h*_) or skin-only infection (*I^a^*_1*h*_). It is assumed that both blood and skin-only infections are infectious to susceptible tsetse, however, skin-only infections have reduced infectiousness to tsetse by a factor (*x*). Unlike the baseline model and model with animal transmission, we also assume that either of these infected types of individuals can self-cure and that this happens at rates *ω^s^* and *ω^b^* for skin-only or stage 1 blood infections respectively. We assume that skin-only infections which do not self-cure become stage 1 blood infections at a rate *θ*. The asymptomatic model therefore has five extra parameters compared to the baseline model. This variant has not previously been fitted to data.

**Figure 1.**
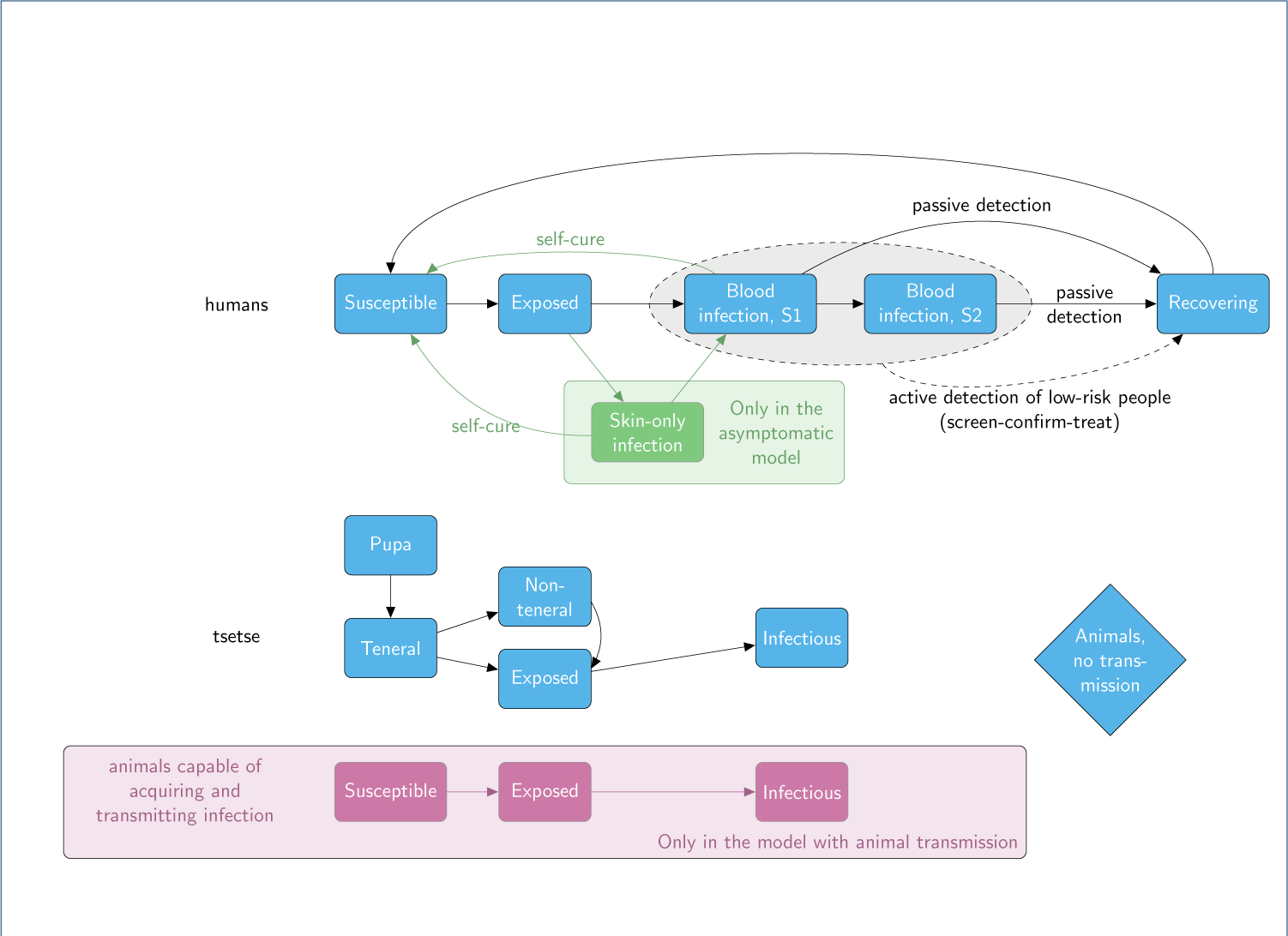
Model diagrams Schematic for the three model variants considered in this study. Blue components form the baseline model and are also included in the other two model variants. The pink boxes and arrows are only found in the animal model and the green box and arrows are only in the asymptomatic model variant. Births, deaths and transmission pathways are not shown to aid readability. Arrows relating to disease/infection progression are shown. The grey oval and dashed lines indicate infection classes assumed to be detectable using a traditional screen-confirm-treat approach in AS (although some infections still may be missed due to imperfect diagnostic sensitivity). An alternative version showing the mathematical notation used in the model and transmission pathways is shown in SI Figure 6.

For each variant, there are both deterministic and stochastic versions. Deterministic models of gHAT, described by systems of ordinary differential equations (see SI), have been predominantly used in the literature due to their quick computational run time and because they are simpler to fit to data. It has also been demonstrated that, despite gHAT being such a low prevalence infection, deterministic and stochastic models of gHAT have very similar mean dynamic behaviour and therefore their ubiquitous use is not unreasonable [40, 41]. Further work has found that fitting deterministic variants of gHAT models but using the analogous stochastic variant for sampling and projections works well to reduce computational challenges but still outputs better estimates for elimination [42]; the main advantage in this instance is for the evaluation of the probability of EoT as it obviates the need for a proxy threshold to determine when the last transmission event occurs. Whilst it is not highly pertinent in the present study, stochastic models of gHAT are also particularly useful when population sizes being modelled are smaller than health zones – for example at village level [43, 44].

In this study, we will use a combined approach to take advantage of both deterministic and stochastic model versions of the three model variants. Fitting of the models to data and assessment of model evidence will be performed using the deterministic model, whilst forward projections to evaluate possible trajectories under different strategies and assess the probability of EoT will be simulated using the equivalent stochastic version, parameterised by the posteriors of the deterministic fit. Specifically, in this work, we are defining EoT as being the first year after the final transmission event occurs in the simulation.

### Data

The WHO define risk thresholds of “high risk” as *>* 1 annual case per 1,000 population, “moderate risk” as 1–10 annual cases per 10,000 population, “low risk” as 1–10 annual cases per 100,000 population, and “very low risk” as 1–10 annual cases per 1,000,000, all averaged for five years [45]. These thresholds apply to spatially smoothed risk areas with 30km radii, which do not align with administrative areas [4]. Although different, one can also look at a similar health-zone-level risk measure of the average number of cases averaged over the last five years per 10,000 people across the health zone – this measure is interesting as *<*1 annual case per year per health zone does correspond with one of the country-level indicators for elimination as a public health problem [6]. Almost all health zones of the DRC were classified as having *<* 10 annual cases on average per 10,000 population during 2011–2015 with only 4 of 516 with *>*10 average annual cases per 10,000. For 2016–2020 cases fell further with only 17 health zones having *>* 1 cases per 10,000 people and none having *>* 10 cases per 10,000. The health zones selected for this model comparison study were chosen as they represent a variety of health-zone-level risk levels observed across the country and they also represent a range of present-day and historical coverages of AS activities. Table 1 shows summary information for each health zone and their geographical position in the country is shown in Figure 2. It is noted that recent (mean) coverage of AS is correlated to risk in two different ways: firstly, one year’s AS is supposed to be dictated by the previous years’ case reporting – according to WHO guidelines, villages reporting cases within the last three years should continue AS, whereas, for those with no reporting in this period and a further year of no case reporting (in year four or five), AS may be stopped [7]. Secondly, if AS is low then there is less chance of finding extant infections, so low coverage will also result in low reporting and therefore lower risk categorisation. We see this played out in the selected health zones – the two health zones with *>*1 annual case per 10,000 for 2011–2015 had higher mean AS, and the two health zones with *<*1 annual case per 100,000 for 2011–2015 had virtually no AS coverage in the last ten years.

**Figure 2.**
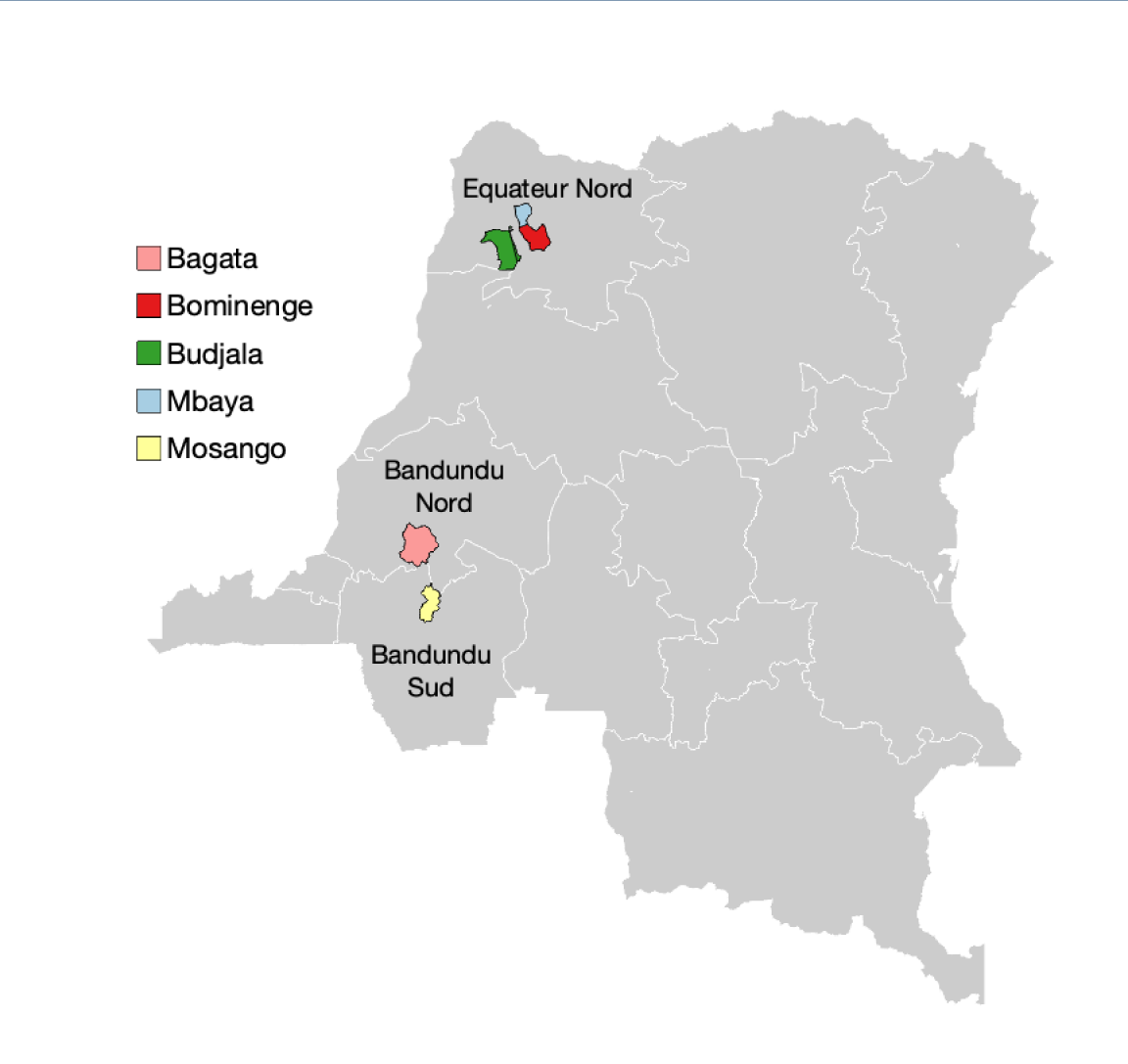
Locations of the health zones considered in this study. The whole DRC is shown in grey with coordination boundaries for gHAT control shown in white. The five health zones under analysis in this study are shown as coloured regions with a black border and the names of the coordinations they are in are labelled.

**Table 1.**
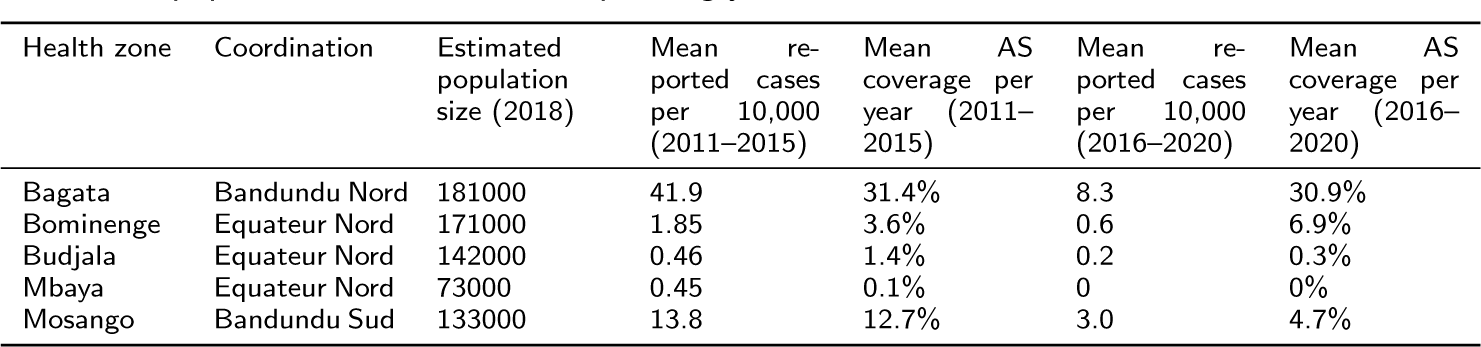
Summary information for the example health zones used in this modelling analysis. The percentages of active screening (AS) coverage are presented here as the mean number out of the health zone population size for the corresponding years.

For each health zone, annual case data for the period 2000–2020 was extracted from the WHO HAT Atlas using geolocation information (where known). The data were aggregated to the health zone level, however, the method of detection (active or passive) and staging information (generally known from 2015) were separated. More information on data extraction can be found in the original fitting paper for the baseline model (Model 4) using 2000–2016 data [39] and in an update paper using 2000–2020 data [46]. None of the five health zones had had large-scale VC implemented before 2020, however, Bagata health zone did commence Tiny Target deployments in mid-2021 and it is possible that there were some minor effects of VC in Mosango health zone due to deployments along a shared river with the neighbouring health zone (Yasa Bonga) since mid-2015. These VC activities have been included in the model simulations – including during the fitting for Mosango, and during projections for 2021–2023 for Bagata (as described in Antillon et al., [46]).

Visualisations for case reporting in the five health zones over time can be found in the Supporting Information.

### Model fitting and evidence

Model fitting of the deterministic baseline (Model 4) and animal (Model 7) variants has already been presented in previous work [39, 31] (using 2000–2016 data) and [46] (using 2000–2020 data), however, the asymptomatic model has not previously been fitted to case data. To directly compare the outcomes of the three model variants we utilise the same adaptive Metropolis-Hastings random walk Markov chain Monte Carlo (MCMC) methodology [47] to fit the asymptomatic model (Model 9) to the same longitudinal case data (2000–2020) for five health zones of the DRC (see Table 1). However, with the asymptomatic model, we follow an additional step called sequential Bayesian updating (SBU) after the initial fitting so that we can share information from health zones that are more informative for this model with health zones with less information. Unlike the animal model, where we believe there should be geographical variation in the extra parameters: the density of non-human animal hosts and the proportion of tsetse feeds taken on them, the five additional asymptomatic model parameters should be intrinsic biological variables and not vary between health zones. Ideally, all health zones would be considered jointly with some form of spatial structure. However, this would be very computationally demanding when performed across the whole of the DRC so SBU was chosen as a more feasible option.

In this approach, we first assess the asymptomatic model fits for the five health zones and rank the health zones in order of how much the asymptomatic parameter posterior distributions have shifted from their priors. We take the health zone where we have learnt the most information (have the highest deviation – measured by the total Kullback-Liebler divergence (*D_KL_*) across the five additional asymptomatic model parameters and reorder this to be our first health zone. We then rank the other four health zones from most to least information learnt using *D_KL_*. After re-ordering we perform refitting. For the health zone with the most information, we can keep the original model fit, however starting from the second highest ranked health zone we first update the five asymptomatic parameter priors to be a multivariate parametric approximation of the posterior parameter distributions from the previous model fit, using a Gaussian mixture model (GMM). Each of the five asymptomatic model-specific parameters was transformed to a (*−∞,* +*∞*) scale and then to have a mean of zero and a standard deviation of one. The GMM was fitted multiple times to the transformed parameters using the MATLAB routine fitgmdist with different numbers of Gaussian distributions and Akaike’s Information Criterion (AIC) was used to select the final number of components in the GMM. After re-fitting the second-ranked health zone using the updated prior we take the new posterior and use this in turn to provide the new prior for the third-ranked health zone. This process of prior updating and re-fitting continues until we have an updated fit for the lowest-ranked health zone. A graphical illustration of this process can be found in Figure 3.

**Figure 3.**
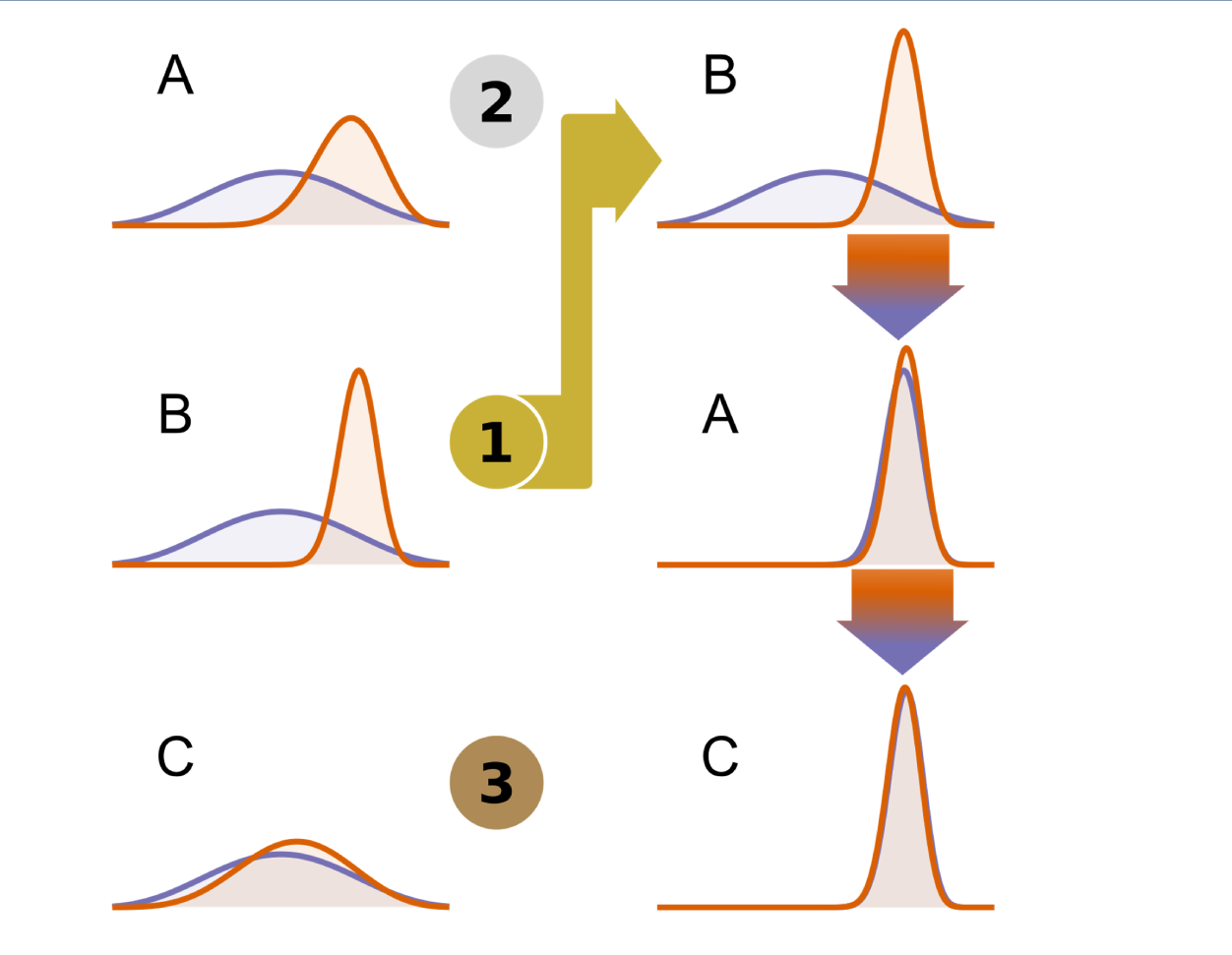
An illustration of the sequential Bayesian updating method used to improve the estimation of the five asymptomatic model parameters. In this example, locations A, B and C have been analysed to estimate a single parameter. In the first round of analyses, the same prior (purple lines) was used in each location. Following this analysis the locations were ranked in terms of how much the posterior distribution (orange lines) diverged from the prior distribution – the more divergence the more information was in the data – such that the order for re-evaluation was B then A then C. Location B did not require re-analysis. Location A was analysed with the posterior for location A acting as the prior. The resulting prior from location A was then used as the prior in a re-analysis of location C. We can see that the posterior parameter distributions for locations A and C are now far more like the ones for the informative location B.

In the MCMC analysis of the baseline and animal transmission models, we ran two chains resulting in 2,000 sets of posterior samples. To facilitate the sequential Bayesian updating we ran 5 MCMC chains in the asymptomatic model analyses to give 5,000 sets of posterior samples, improving the chances of sampling in the extreme tails of the parameter distributions and consequently incorporating this information into our multivariate priors.

Following the fitting of all three model variants to the five health zones we use the model evidence, or marginal likelihood, to compare the fit of the models. Importance sampled estimates of the model evidence [48] were generated, using a defence mixture [49] consisting of a weighted combination of a multivariate Gaussian mixture model fitted to all of the posterior samples (weight=0.95) and the prior distributions of the fitted parameters (weight=0.05) [31]. To create the weighted ensemble model in each health zone, samples were randomly selected from the individual model posterior samples in proportion to their relative model evidence to give an ensemble of 2,000 posterior parameter samples. All of the projected samples associated with each of these posteriors from the individual model runs, 10 for each posterior set, were taken to produce the ensembled projection results, i.e. 20,000 ensemble realisations.

### Model projections

To assess the impact of possible cryptic transmission on future case reporting and elimination, we utilise the posteriors from fitting the different model variants and project forwards using the stochastic model version.

Many different strategies may be considered to control gHAT, comprised of passive screening in fixed health facilities, AS at different levels of coverage, the introduction of safer, single-dose drugs, and the use of VC. Here we will consider only a few strategies, all of which assume that present-day passive screening continues to operate at the same level (see Table 2).

**Table 2.** Strategies considered for forward projections (2024–2050). AS = active screening. S&T = screen-and-treat. VC = vector control. Our default strategy, *MeanAS*, represents a “continuation” strategy based on recent activity in the health zone.

| Strategy name | Passive screening | AS coverage | AS diagnostic algorithm | Vector control‡ |
| --- | --- | --- | --- | --- |
| MeanAS* | Continues at present levels | Mean of 2016–2020 | Parasite confirmation needed for treatment. Assumed 91% sensitivity and 100% specificity† | None |
| MeanAS + VC | Continues at present levels | Mean of 2016–2020 | Parasite confirmation needed for treatment. Assumed 91% sensitivity and 100% specificity† | 80% tsetse reduction after 1 year starting in 2024 |
| MeanS&T | Continues at present levels | Mean of 2016–2020 | Treatment may be given with positive RDT from 2028. This is assumed to increase the algorithm sensitivity to 95% and decrease specificity to 99.5%. | None |

\*default strategy
†The active screening diagnostic algorithm specificity is fitted to 2000–2020 data, and is assumed to increase to 100% from 2015 in Mosango, 2018 in Bagata, and 2024 in the other health zones simulated.
‡ Vector control began in Bagata in mid-2021 and Mosango had low-level inadvertent benefit from vector control along shared rivers with a neighbouring health zone from mid-2015. We assume this stops from 2024 in the *MeanAS* and *MeanS&T* strategies.

Under each model variant, AS is assumed to be conducted by mobile teams which attend villages and screen a certain number of people each year. All of our models have a high-/low-risk structure and assume that only low-risk people participate in AS. A key difference between the strategies for AS is which algorithm is used. In *MeanAS* and *MeanAS+VC*, the current algorithm based on an initial screening test (CATT or RDT) followed by parasite confirmation is simulated. This algorithm is assumed to have a sensitivity of 91% and the specificity is fitted. Whilst this sensitivity is relatively high, some true infections are likely to get missed from AS, especially if only the low-risk group is repeatedly screened each year. We assume that 100% specificity is possible to achieve with additional measures in place including video confirmation of the parasite via computer tablets which can be validated by others. We assume this happened in 2018 in the Bandundu Nord and Sud coordinations based on the historical availability of computer tablets for this activity and would happen elsewhere when case detections are very low (see SI).

The S&T algorithm simulates the treatment of all individuals presenting to screening with a positive screening test (CATT or RDT). At present this is not possible, however, it is hoped that the introduction of a single-dose oral cure, acoziborole, could make this option feasible in the future. In these simulations, we assume that removing the need for a confirmation test before treatment would lead to higher sensitivity (95%) but sacrifice specificity (99.5%). This is expected to result in some “over-treatment” (treatment of false positives) but has the potential to reduce transmission more quickly by treating more of the truly infected individuals. We assume that post hoc laboratory testing via trypanolysis (or similar) would be performed after serosuspects are treated and that only laboratory-confirmed infections would count towards case reporting – this assumption around post hoc confirmation has no direct impact on transmission in the model, however, could make a large difference in number of “cases” reported. We also assume that those serosuspects (who test CATT or RDT positive) but with non-detectable blood parasiteamia would be equally likely to be confirmed as cases using the highly-specific immune trypanolysis as those with detectable blood infections.

Under the asymptomatic model, the *MeanS&T* algorithm is particularly appealing as it has the potential to detect and enable treatment of skin-only infection where there is no detectable blood-parasitaemia [50]. Under the standard AS algorithm, only those with blood infections have a chance of being diagnosed and treated due to the parasite confirmation criterion. The asymptomatic model still has a high-/low-risk structure so *MeanS&T* cannot directly combat high-risk individuals not presenting to screening.

Infected people in high-risk groups are assumed to be detected only through passive screening based on self-presentation after symptoms develop and are severe enough to seek medical attention. In all models, we assume that late-stage infections (stage 2) are more likely to be detected than early-stage infections and the relative detection rate from early and late-stage infections is fitted to the human case data. We assume that passive case detection rates remain the same even if acoziborole becomes available. Under the asymptomatic model, we assume that skin-only infections have no discernible symptoms that could result in a gHAT diagnosis and treatment.

In this study, we simulate the introduction of VC through Tiny Targets for strategy *MeanAS+VC*. We assume that Tiny Targets are deployed twice a year and achieve an 80% reduction in total tsetse population after one year, which is slightly more conservative compared to previous reductions observed in the field in several locations (e.g. *>*85% in Yasa Bonga health zone in the DRC [51], *>*90% in Uganda [52], *>*99% in the Mandoul focus of Chad [32]), *>*95% in Ĉote d’Ivoire [53], and comparable to the reduction observed in the Boffa focus of Guinea [54]. The detailed model formulation we use can be seen in the Supplementary Information and is presented elsewhere [39, 55].

For the period 2021–2023 for which we do not have data but has already occurred we simulate the continuation of the current strategy in all simulations.

By drawing samples from the posterior parameter set for each health zone and for each model variant, we run each projection strategy from 2024–2053. We run the model 10 times for each posterior sample, giving a total of 20,000 realisations for the baseline and animal transmission models and 50,000 realisations for the asymptomatic human transmission model and therefore we incorporate both parameter and stochastic (chance) uncertainty in our predictions.

We track the number of new human infections occurring each year in the model and use this to assess when EoT has been met for each iteration. We say that EoT has been met when 10 consecutive years with no new human infections are produced in the iteration and so can assess the probability of EoT up until 2040. This method was previously used to assess the likelihood that EoT had been or would be met in three health zones of the Equateur Nord coordination (Budjala, Bominenge and Mbaya) using the baseline model elsewhere [56].

Across all three models and all five health zones under the continuation strategy, we found that 99.95% of simulations which had the last year of transmission between 2000 and 2040 achieved elimination of infection within 13 years of reaching EoT. This being all realisations under the baseline and animal models, and 99.91% under the asymptomatic model. For simulations of the asymptomatic model that achieved elimination of infection under the continuation strategy, the average time between the last year of transmission and elimination of infection was 4.03 years (95% PI: 0– 14 years), with the delay being less than or equal to 13 years in 97.4% of realisations. For the baseline and animal models, the means were 1.50 (95% PI: 0–4 years) and 1.33 years (95% PI: 0–4 years). Therefore, we believe that using the last year of transmission in our simulations as the EoT year is reasonable even for EoT in 2040. This approach is a little different to the modelled cut-off for EoT presented in Castano et al., [41] which used the first year after which there were five consecutive years of no transmission as the metric of EoT in the simulations; this approach in Castano et al. will slightly overestimate EoT probabilities. The approach in the present study will give more accurate probabilities, especially for the earlier years but there is a very marginal bias in the later years towards overestimated EoT probabilities.

## Results

### Model fitting

The model captures the different trends in case reporting well across the 21 years despite the qualitative differences between health zones (see Figure 4 and SI Figures 13–17). All model variant fits to the data look very similar and, visually, there are no obviously better or worse fits for each health zone. The main difference we see is the difference in the number of new annual human infections with slightly higher credible intervals for the asymptomatic model compared to the baseline model or model with animal transmission. We use the relative model evidence to assess support for the different models, despite their apparent similarity. Figure 5 shows the results of this for the five different health zones. We notice that (a) all models have *<*13% support for the animal model, (b) Mosango, Budjala, Bagata and Mbaya all have somewhat similar support for the asymptomatic and baseline models, and (c) Bominenge, the most informative health zone for the asymptomatic model based on *D_KL_*, has almost all of its support for the baseline model. Despite the inconclusive nature of this model evidence analysis, by looking at the five asymptomatic model-specific parameter posteriors for each health zone (see SI Figures 8–12) we notice that some of them pull away from the prior in the direction that means less contribution from asymptomatics to transmission (i.e. lower relative infectiousness (*x*), higher probability of developing blood infection (*p_bs_*) and higher rate of self-cure from skin-only infection (*ω^s^* )).

**Figure 4.**
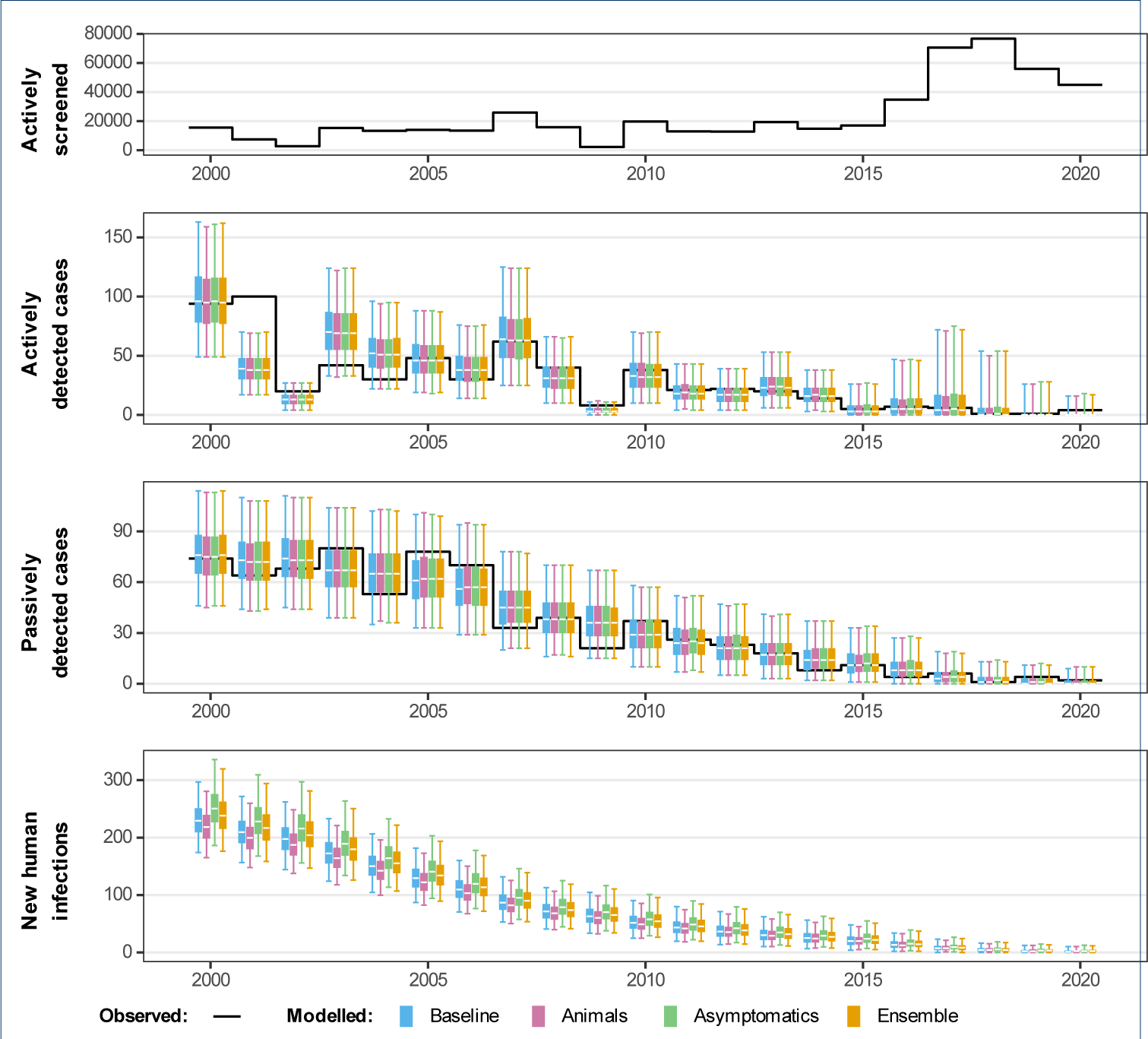
Comparison of fits in Mosango. The deterministic model was used to perform fitting and sampling was conducted by using the stochastic model with the fitted posterior distributions. Blue, pink and green box and whisker plots show the baseline model, model with animal transmission and asymptomatic model fits respectively. The orange boxes represent the ensemble model outputs. The central line of each box is the median, the box is the 50% credible interval (CI) and the whiskers show the 95% CI. Case data are shown as a black line. New infections are estimated through the model fit, however, there is no way to directly observe this so there are no corresponding observational data.

**Figure 5.**
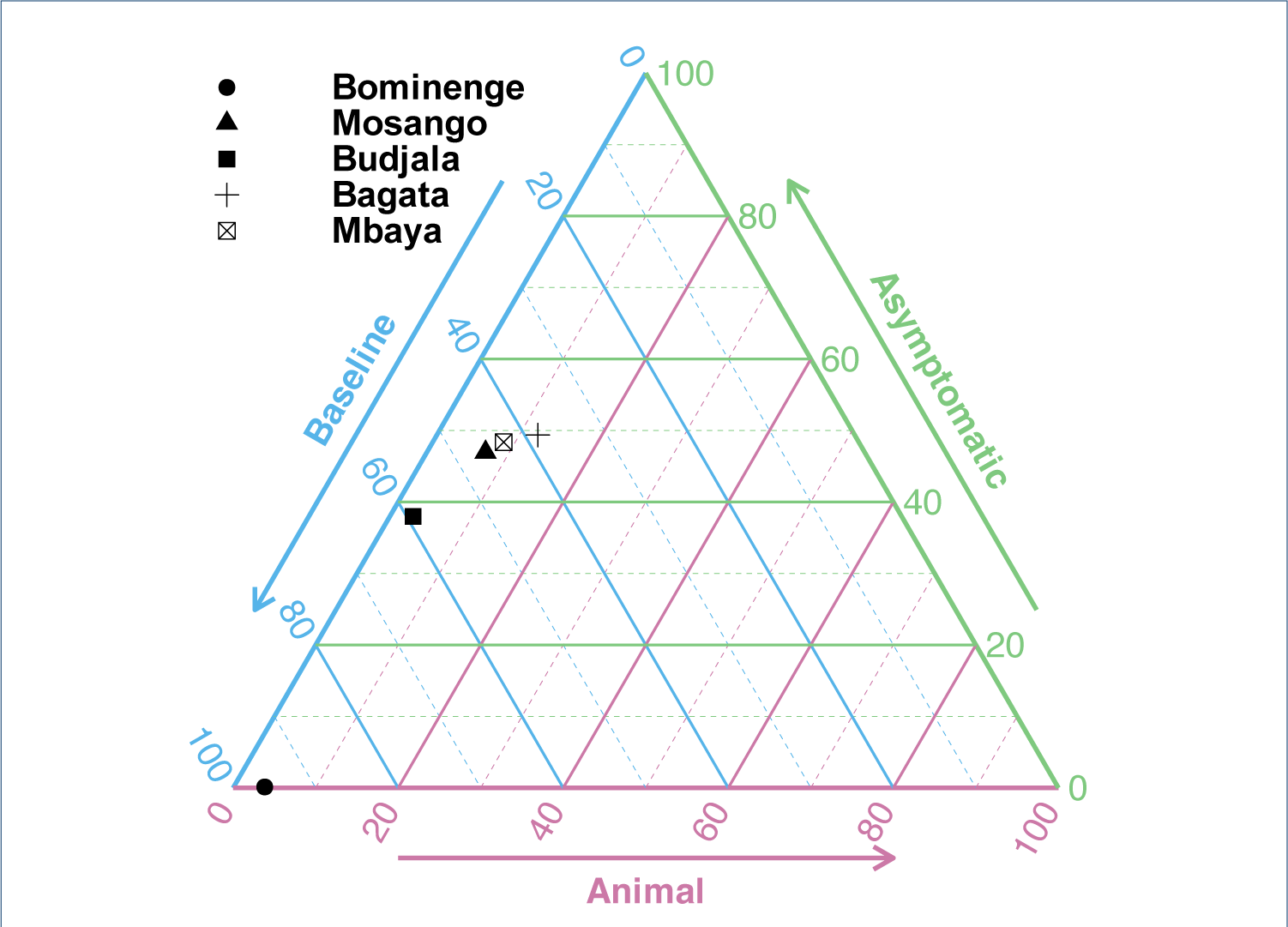
Ternary plot of relative evidence from the three models. The model evidence for the baseline model and the models including animals or asymptomatic human infections contributing to transmission are scaled such that the sum is 100%. These three values are transformed and plotted on an equilateral triangle with edges forming the axis for each model. To read off the relative support for one model variant pick the axis for that model and trace the corresponding coloured line back to the point; e.g. for Bominenge (circle), there is 4% support for the animal model (this is sitting on the animal axis), 0% support for the asymptomatic model and 96% support for the baseline model. For Bagata (cross) there are 12%, 49% and 38% for the animal, asymptomatic and baseline models respectively.

In our initial fit of the asymptomatic model, we found that, whilst in some health zones the additional five asymptomatic posteriors pulled away from the priors in a direction indicating smaller contributions from asymptomatics to transmission than our initial belief (lower relative transmissibility, quicker self-cure rates and a higher probability of being a blood-detectable infection), for other health zones the asymptomatic model parameter posteriors followed the priors more closely – meaning little information had been learnt about them during fitting (see SI Figure 7). By ranking the health zones based on the total Kullback-Liebler divergence of posterior distributions from prior distributions for the five parameters specific to the asymptomatic model (see SI Table 3) and then refitting using SBU we passed some of the information present in Bominenge’s data, and sequentially in other health zones, down to the least informative health zones (Bagata and Mbaya) and this meant that the SBU posteriors for Bagata and Mbaya no longer followed the original prior closely, see SI figures 8–13. There was little information in any of the data sets on the transition rate from skin infections to blood infections (*θ*) and the self-curing rate in blood infections (*ω^b^* ), the posterior distributions for these parameters very closely followed the prior distributions in all health zones and all analyses, before and with SBU.

### Model evidence and ensemble model outputs

The model evidence for each model was converted into relative model evidence values, such that the within-health zone sum of the model evidence was 100, and then used principally in two ways: to compare the statistical support for each model in each health zone and to produce an ensemble model. The calculation of the relative model evidence assumes equal prior weight on each of the three models, in which case our ensemble model is a Bayesian model average. The relative model evidence is presented as a ternary plot in Figure 5. Alternatively, the log model evidence can be summed across health zones within a model before normalising to give an across-health zone relative posterior probability of each model. These values are 99.9%, 0.002% and 0.126% for the baseline, animal and asymptomatic human transmission models.

Given the similarity of the individual model fits to the historical data, the ensemble model’s case reporting during the fitted period is also very similar to the individual model outcomes whereas the estimated new infections lie between the baseline and asymptomatic model in most health zones (see Figure 4 and SI Figures 13–17).

### Model projections under a continuation strategy (*MeanAS* )

Next, we used our model projections under a continuation strategy (*MeanAS*) to assess how the use of the different model variants affects the model predictions. Comparing the case reporting of the three model variants for 2021–2035 from the stochastic model (see Figure 6 and SI Figures 18–32) shows that they all overlap, however, there are some noticeable differences for the three models.

**Figure 6.**
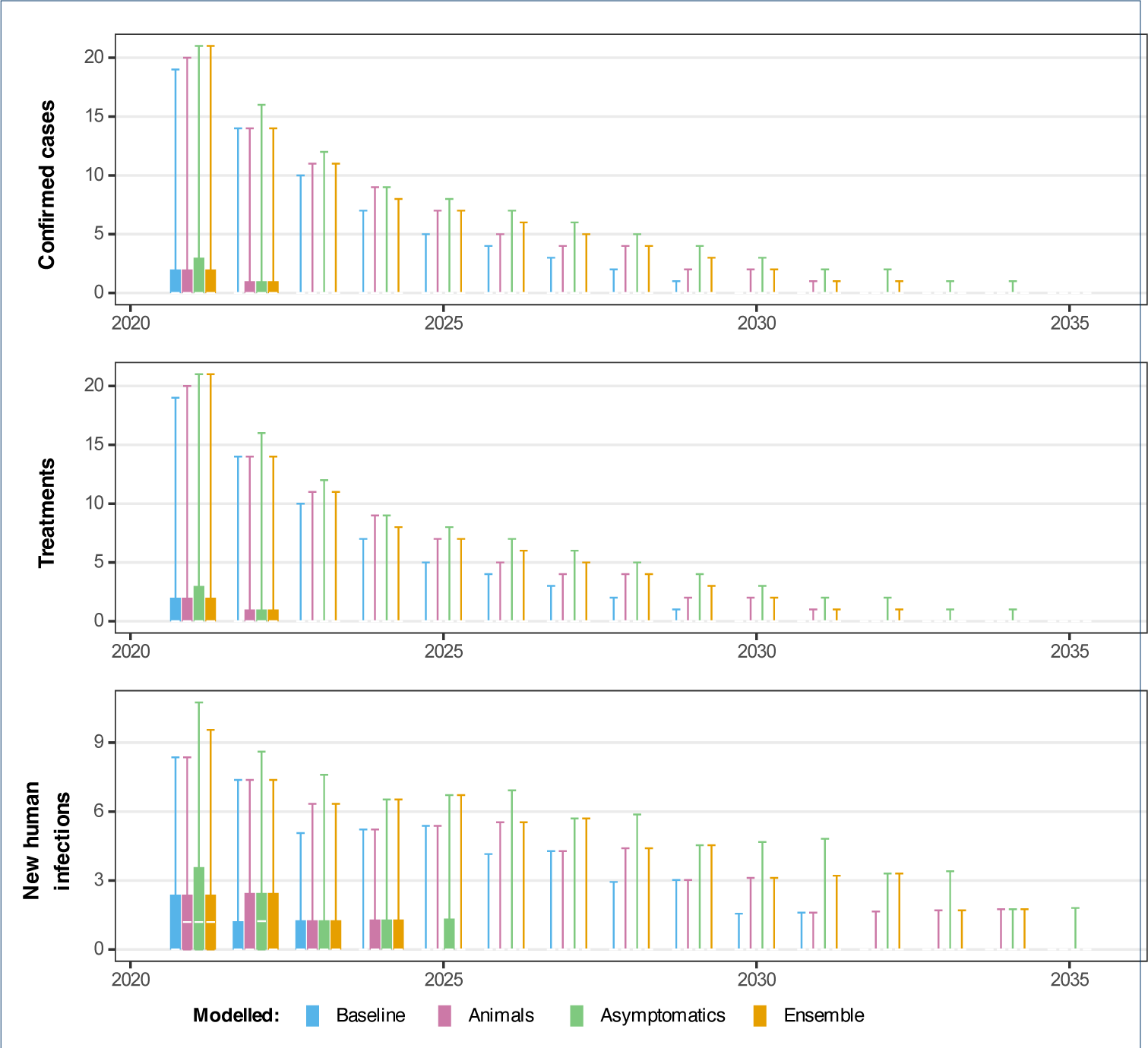
Projected dynamics in Mosango health zone in Bandundu Sud coordination under Mean active screening strategy. Comparing three model variants using the stochastic model including projections for 2021–2035 under a MeanAS strategy (using AS coverage for Mosango from 2016–2020). Blue, pink, green and orange box and whisker plots show the baseline model, model with animal transmission, asymptomatic model and ensemble model projections respectively. The central line of each box is the median, the box is the 50% prediction interval (PI) and the whiskers show the 95% PI.

The differences in 2021–2035 are very small in all health zones although Bagata and Mosango are expected to have marginally higher case reporting under the animal and asymptomatic model variants. The differences in estimated new human infections are also most visible in Bagata and Mosango, but this is not so clear in the other health zones.

It is clear that for Mosango, the baseline model produced the most optimistic probability of EoT by 2030 (96%), followed by the model with animal transmission (90%) and finally the asymptomatic model is the least optimistic (84%) (Figure 7. Bominenge and Budjala also have the asymptomatic model as the most pessimistic but with no difference between the baseline and animal models, whereas Bagata and Mbaya have the animal model as being more pessimistic than the asymptomatic model (see SI Figure 33).

**Figure 7.**
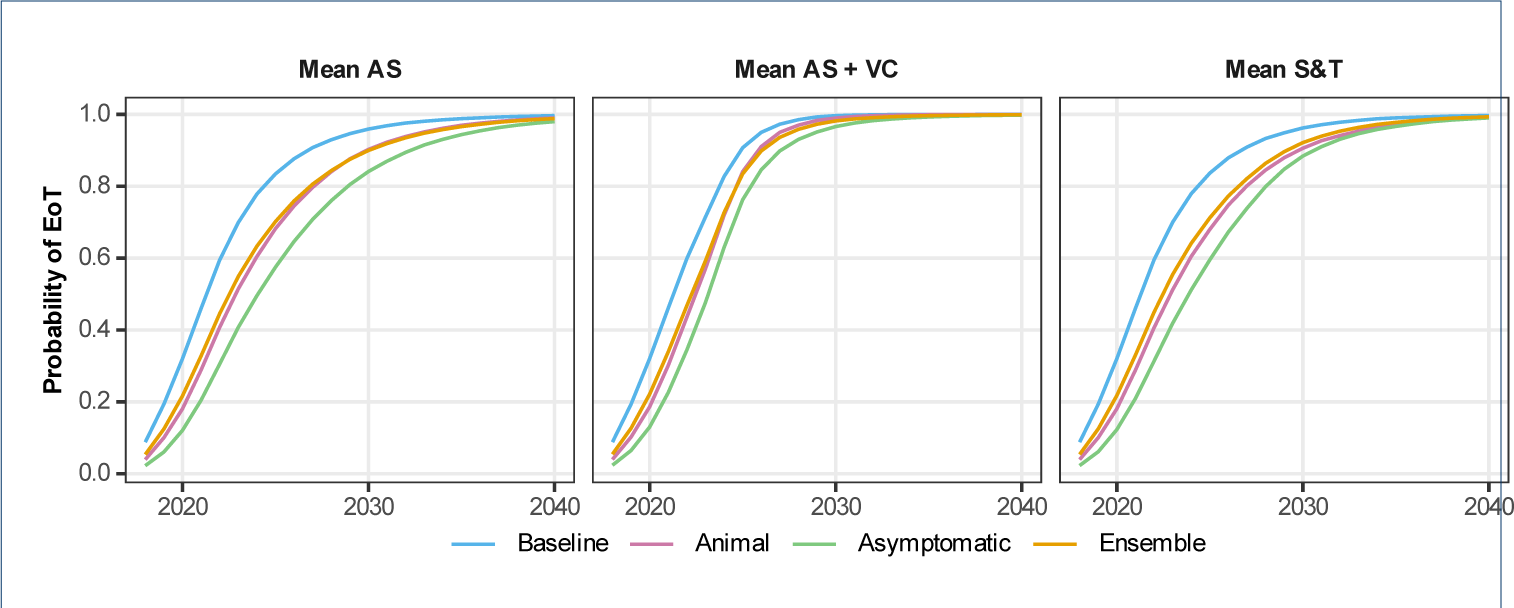
Comparing the probability of EoT in Mosango health zone, under each model variant and three different strategies. The blue, pink and green curves represent the model-estimated probability of EoT by each year, calculated by taking the number of realisations where there are no new infections to humans in or after that year until the end of the simulation and diving by the total number of realisations (20,000 for the baseline and animal models and 50,000 for the asymptomatic model) The ensemble results are given by the orange curve which is computed as the weighted average of the probability of EoT from the individual model variants. As per Table 2, for the second strategy with vector control (VC), we assume this novel intervention begins in 2024, and for the third strategy using screen-and-treat (S&T) we assume this novel intervention begins in 2028.

Unsurprisingly, Bagata, which is one of the health zones classified as high riskbased on 2011–2015 reporting, has lower probabilities of EoT (all model variants are under 93% by 2030 for the *MeanAS* strategy. Mosango (the other high riskhealth zone) has around 96% probability of EoT by 2030 under the baseline model, whereas the health zones classified as moderate or low riskfor 2011–2015 all have a high probability of EoT (around 99%) under the baseline model for the same *MeanAS* strategy.

### Model projections under other strategies

In line with previously published work, the addition of VC through relatively rapid and high levels of tsetse reduction has a dramatic impact in turn on new infections to humans (Figure 7 and SI Figure 33). In all model variants, we predict that the simulated 80% reduction of tsetse would quickly curtail transmission via all possible hosts, including putative non-human animals and/or asymptomatic humans. This intense intervention may not be needed in all settings, however, it may prove a powerful tool in regions of persistent transmission, whatever the cause.

This is the first time the Warwick gHAT model variants have been used to predict the impact of an S&T strategy on infection dynamics. Figure 7 shows how the outcome may be more noticeable for the asymptomatic model compared to the other variants. In the baseline model and the model with animal transmission, *MeanS&T* does little to the projected probability of EoT. The main difference between *MeanAS* and *MeanS&T* in those models is the slight improvement to AS algorithm sensitivity (91% to 95%), however, the model assumption that high-risk people still do not present in AS overwhelms this small improvement. In contrast, in the asymptomatic model, the change to S&T also allows for the treatment of infections which are not detectable in the blood and would not have met the confirmation threshold in current AS algorithms. For Bominenge, Budjala and Mbaya, it is predicted that there is already a high probability that the health zone has achieved EoT in the simulations when S&T begins in 2028 so we don’t see an impact of this novel screening approach (see SI Figure 33).

## Discussion

In this study, we have fitted a gHAT model with asymptomatic transmission to data from the DRC for the first time and compared it to models without asymptomatic and with or without animal transmission. Using human case data from five health zones of the DRC from 2000––2020 we have concluded that there is minimal statistical evidence for animal transmission in these locations, however, when we create our ensemble model for these five health zones there is between 0% (in Bominenge) and 49% (in Bagata) of these samples selected from the model with asymptomatic transmission. By using sequential Bayesian updating to learn information about asymptomatic parameters between different health zones, our results indicate that whilst the model results with asymptomatics are more pessimistic about the elimination of transmission compared to the baseline model with continued medical strategies, the probability of elimination by 2030 in each location is only a little lower.

The relative posterior probability of the models highly favours the baseline model (99.9% probability). This is, of course, dependent on both our choice of health zones and our small set of models (even within the scope of this study we rejected the alternative model in which the asymptomatic parameters vary between locations). There may be alternative models that better represent disease transmission, and the results may vary in other locations (the current result is heavily influenced by the outcome in Bominenge health zone) if there are informative locations that give more support to the animal or symptomatic human transmission models.

The ensemble model represents appropriately weighted results taking account of uncertainty about which model to favour appropriately. The use of an ensemble model here makes our outcomes slightly more pessimistic compared to the baseline model. Fortunately, this appears to suggest the worst-case scenario of the asymptomatic model having a lower endemic equilibrium but being prevented from reaching zero cases, discussed in previous modelling work by Aliee et al. [34] through a model sensitivity analysis, does not appear to be the case in these regions of the DRC and the impact of asymptomatic transmission is to slightly delay elimination rather than prevent it. Furthermore, as we believe the asymptomatic model parameters will be the same across the DRC since they relate to the parasite-human interaction and are not dependent on the local geography, this modelling indicates that we should expect qualitatively similar results across health zones for the DRC with the asymptomatic model if we can assume that the most informative health zones will continue to be those that do not favour the asymptomatic model.

In future work, we suggest that this model could be used for fitting in other locations in the DRC and for other endemic countries to capture this possible hindrance to interventions happening across these settings targeted at elimination. It could be that human-parasite interactions in West Africa are sufficiently different that these relatively optimistic results will not extrapolate to countries like Guinea or Ĉote d’Ivoire. More region-specific data on asymptomatic and skin infections like that collected in Guinea [50] could improve our priors on some fitted parameters and would supplement routinely collected case data.

### Model assumptions

The model presented here was fitted to health zone level data, where we assumed independence between neighbouring health zones. Whilst there could be some movement of people and therefore cross-infection between different locations, we believe this effect to be very small. Previously a small between-village importation rate was estimated for gHAT in the DRC [43] and we would expect movement to reduce further for larger spatial scales. Ideally, this model could be used at the health area level (around 10,000 people) to provide more targeted predictions on a similar scale to intervention planning by the national programme. Recent work has demonstrated that the baseline model framework presented here would be suitable for use with health area fitting [42].

It is noted that in the previous model fitting and comparison of the baseline and animal transmission models (but with fewer years of data), Mbaya health zone had strong support for animal transmission, and Bagata had weak support for animal transmission. Budjala, Bominenge and Mosango had substantial or strong support for the model without animal transmission [31]. It is unclear whether previous support for the animal model variant will correlate with support for the asymptomatic model variant, however, it is possible that in these health zones case reporting has not fallen as much as would be expected and one explanation is cryptic transmission by either undetected (or even undetectable) human infections or by animals.

The present study does not aim to make specific policy recommendations, but to explore structural, parameter and stochastic uncertainty for a variety of settings in the highest-burden country for gHAT (see SI Table 4 for details on the ways this research meets the Policy-relevant items for reporting models in epidemiology of neglected tropical diseases (NTD-PRIME) [57]). Future strategies are selected for illustration of the general potential impact on reporting and new infections. To use this framework for guiding policy, the fitting would need to be performed for all health zones (or health areas) using the latest available data.

### Interventions

We have assumed that S&T will become possible from 2028, however, in practice, we do not yet know if and when acoziborole will be able to be rolled out in such a manner. Our results here are illustrative of the type of effect we might expect to see across different regions with this type of intervention. Even after acoziborole is rolled out, it is possible that some groups in the population (e.g. infants and pregnant people) may still require parasitological confirmation before treatment. Conversely, we did not explicitly simulate S&T in passive screening in this analysis, but this intervention could reduce attribution between initial screening and receiving treatment and consequently, it could improve passive detection rates.

As routine data on skin infections is not collected, we have a large amount of uncertainty about how good the CATT and RDTs are at detecting skin infections (sensitivity), although data from Guinea suggest there is very good correspondence between RDTs and confirmed skin infections [50]. More data on this link could improve our assumptions which are currently that the screening test sensitivity is the same for both skin-only and blood-detectable infections.

VC interventions target transmission to and from all host types and could be particularly effective at reducing transmission if there are animal contributions or asymptomatic human infections. However, widespread VC is infeasible over a short time period. Health zones like Bagata could conceivably add VC to their strategy as other nearby health zones have recently put this in place, but the extra resources (both financial and for trained personnel) are non-negligible for this kind of scale-up. Currently, there are no health zones in Equateur Nord coordination province with large-scale VC in place. It may not represent a cost-effective use of resources to deploy Tiny Targets in very low-burden settings in terms of $*/DALY* given the opportunity costs (i.e. the DALYs that could be averted by spending on other higher-burden health zones or other diseases) [58]. As with many diseases at the end game, pushing to zero is likely to represent fairly large costs for minimal DALY reduction, but has the advantage of being able to (eventually) scale back programmes in the long term. The cost-effectiveness of EoT is a complex issue [59]. Recently the Food and Agriculture Organization of the United Nations (FAO) and WHO convened an expert meeting on VC against gHAT and concluded that more criteria and approaches are needed to prioritise regions for VC [60]; we hope that the present study and other modelling and health economic analyses can support this aim through quantification of benefits and costs in different locations.

In the present study, we have simulated “continue forever” strategies but we will need a cessation of vertical interventions at some point – particularly we need a confirmation method if switching to S&T strategy as otherwise we will have false positive reporting forever with high screening coverage. Our group’s other work which is designed to support specific decision making does factor in cessation [58, 46], however, this was not the focus of this analysis.

## Conclusion

Whilst recent evidence suggests that some people can harbour *gambiense* trypanosomes in the skin and have undetectable blood parasitemia, the modelling work presented here suggests that such infections do not play a large role in transmission, if any. We cannot rule out some level of asymptomatic transmission but we expect the impact of this on elimination targets to be relatively small. Likewise, there is some small predicted delay to elimination if we simulate animal transmission in the model, however, in these five health zones of the DRC, it appears relatively unlikely that non-human animals are contributing to transmission.

If there is some asymptomatic transmission, a screen-and-treat strategy with a safer new drug would be expected to be more beneficial compared to if there is no asymptomatic transmission. For infections arising from asymptomatics, nonhuman animals or people not participating in screening, vector control could help to reduce transmission quickly although it should be coupled with suitable detection and treatment and will not be necessary in all settings.

## Ethical approval

Ethics approval was granted by the University of Warwick Biomedical and Scientific Research Ethics Committee (application number BSREC 80/21-22) to use the previously collected DRC country HAT data, provided through the framework of the WHO HAT Atlas [6], in this secondary modelling analysis. No new data collection took place within the scope of this modelling study.

## Availability of data and materials

Data cannot be shared publicly because they were aggregated from the World Health Organization’s HAT Atlas which is under the stewardship of the WHO. Data are available from the WHO (contact or visit https://www.who.int/trypanosomiasis_african/country/foci_AFRO/en/) for researchers who meet the criteria for access. Model code and outputs produced from this study are available through Open Science Framework https://osf.io/73ytc/.

## Author’s contributions

- Conceptualisation KSR
- Methodology MA, REC, SEFS, KSR
- Software MA, REC, SAS, KSR, CH
- Validation
- Formal Analysis MA, REC, KSR
- Investigation
- Data Curation REC, CS, EMM
- Writing Original Draft KSR
- Writing Review and editing KSR, REC, CH, EHC, SAS, SEFS, MA, MJK, EMM, CS
- Visualisation MA, REC, SAS, KSR
- Supervision MJK, KSR
- Project administration KSR, EHC
- Funding acquisition KSR, MJK, SEFS

## Acknowledgments

The authors thank PNLTHA-DRC for original data collection and WHO for data access (in the framework of the WHO HAT Atlas [45]).

## Rights retention statement

For the purpose of open access, the author has applied a Creative Commons Attribution (CC-BY) licence to any Author Accepted Manuscript version arising from this submission.

## Additional Files

Additional file 1 — Supplementary methods, results and NTD-PRIME checklist

This pdf contains further details of modelling methods and additional results graphics. It also includes the NTD-PRIME checklist for good practice in reporting modelling for policy for NTDs.

