## Additional File 1 for "Modelling timelines to elimination of sleeping sickness in the DRC accounting for possible cryptic human and animal transmission"

#### Additional information on methods

##### Data

The data used in this study were extracted from the WHO HAT Atlas for DRC. Extraction was performed previously for 2000–2016 data and was described in the original baseline model fitting publication [2]. The same approach was used for 2000–2020 data. In summary, the individual records of data were aggregated at the health zone level and for each year based on recorded geolocations (where known) or location descriptions. Cases were classified as active or passive and each was labelled as "stage 1", "stage 2" or "stage unknown". In general, before 2015, staging information is unknown but is present in most data for 2015 and 2016. We also record the total number of people listed as tested by the active screening teams for each health zone for each year.

Figures 4–5 show the extracted and aggregated data for the five health zones of this study that were used for fitting.

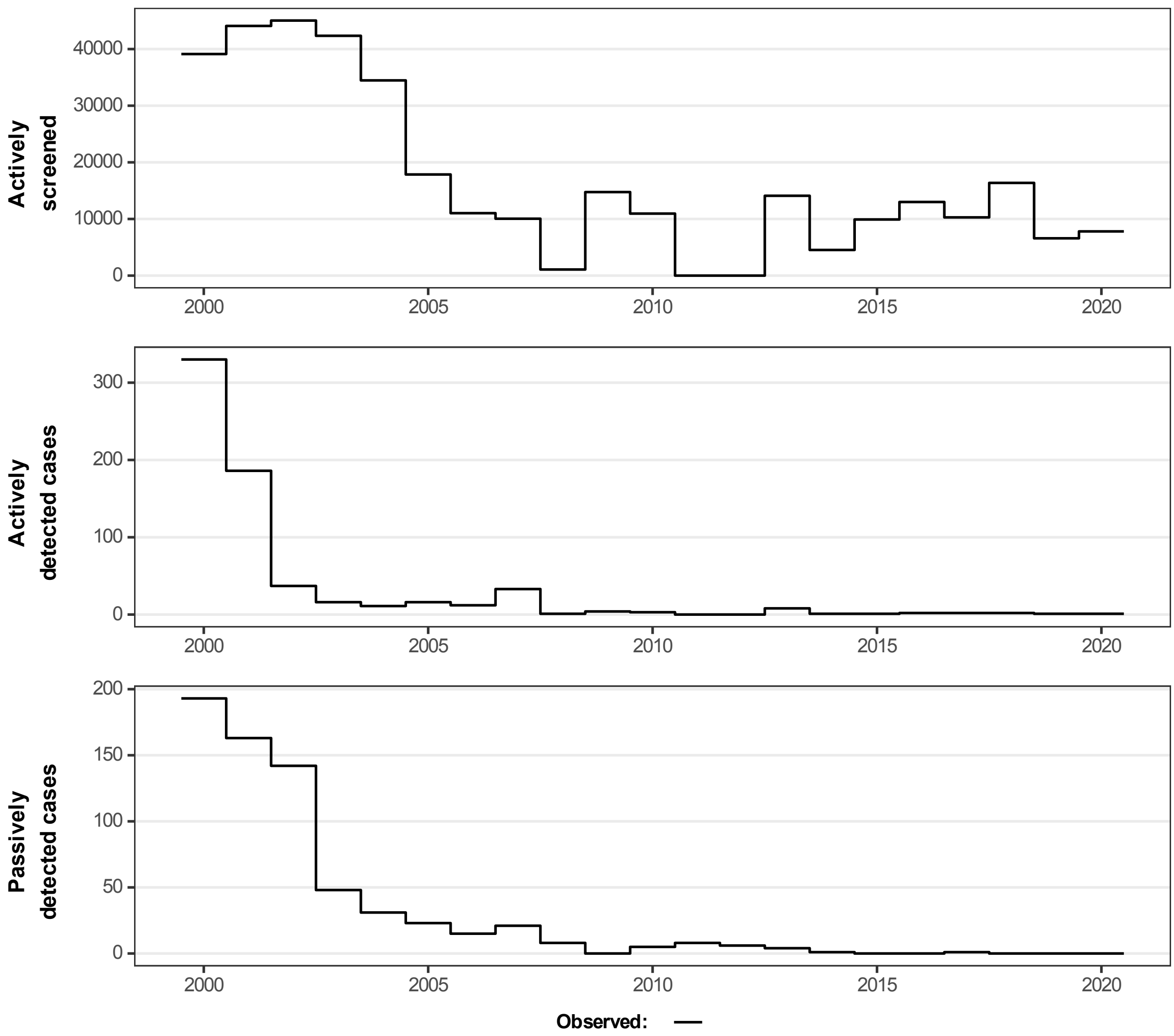


Fig 1: Aggregated data used for model fitting in Bominenge.

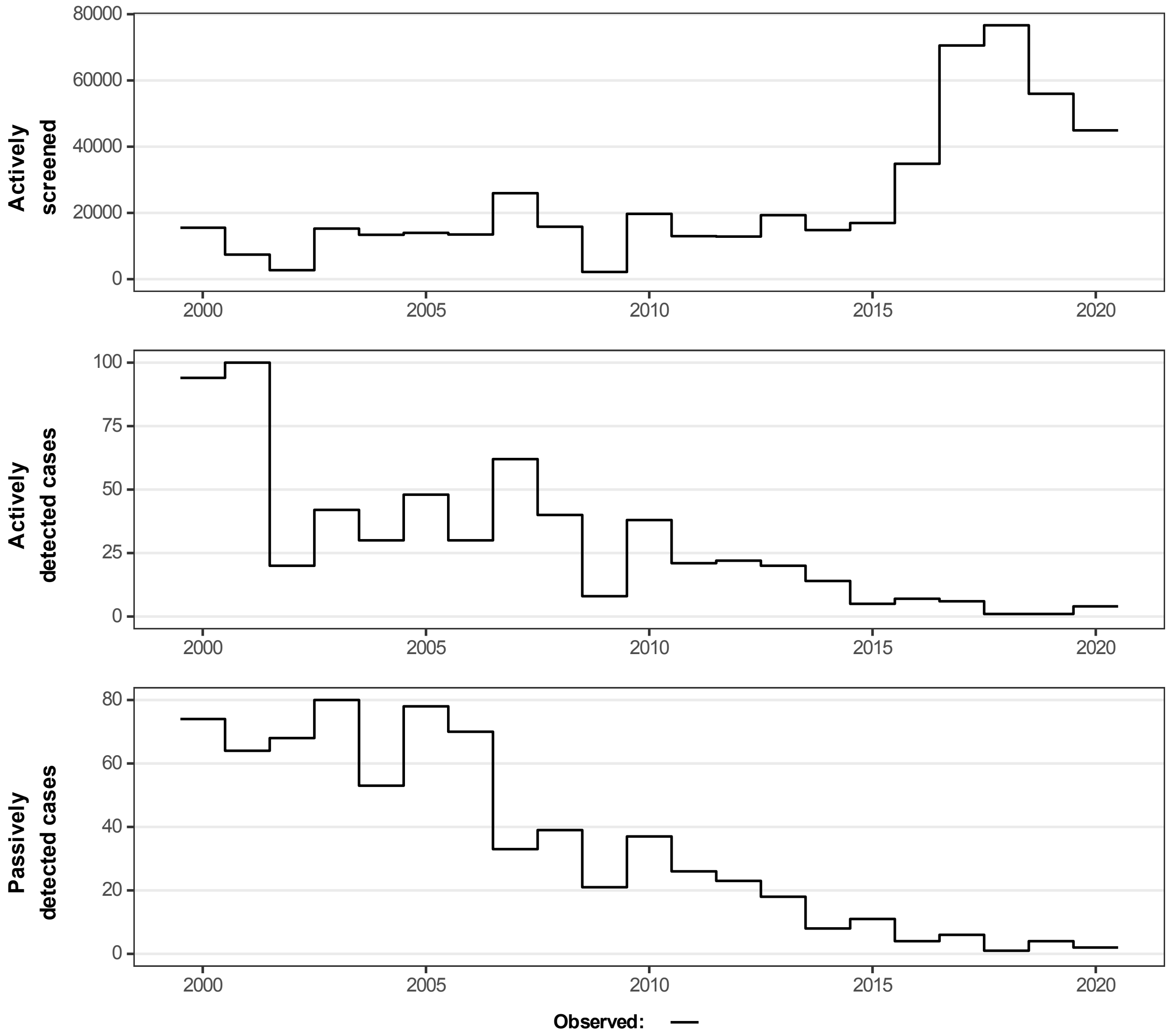


Fig 2: Aggregated data used for model fitting in Mosango.

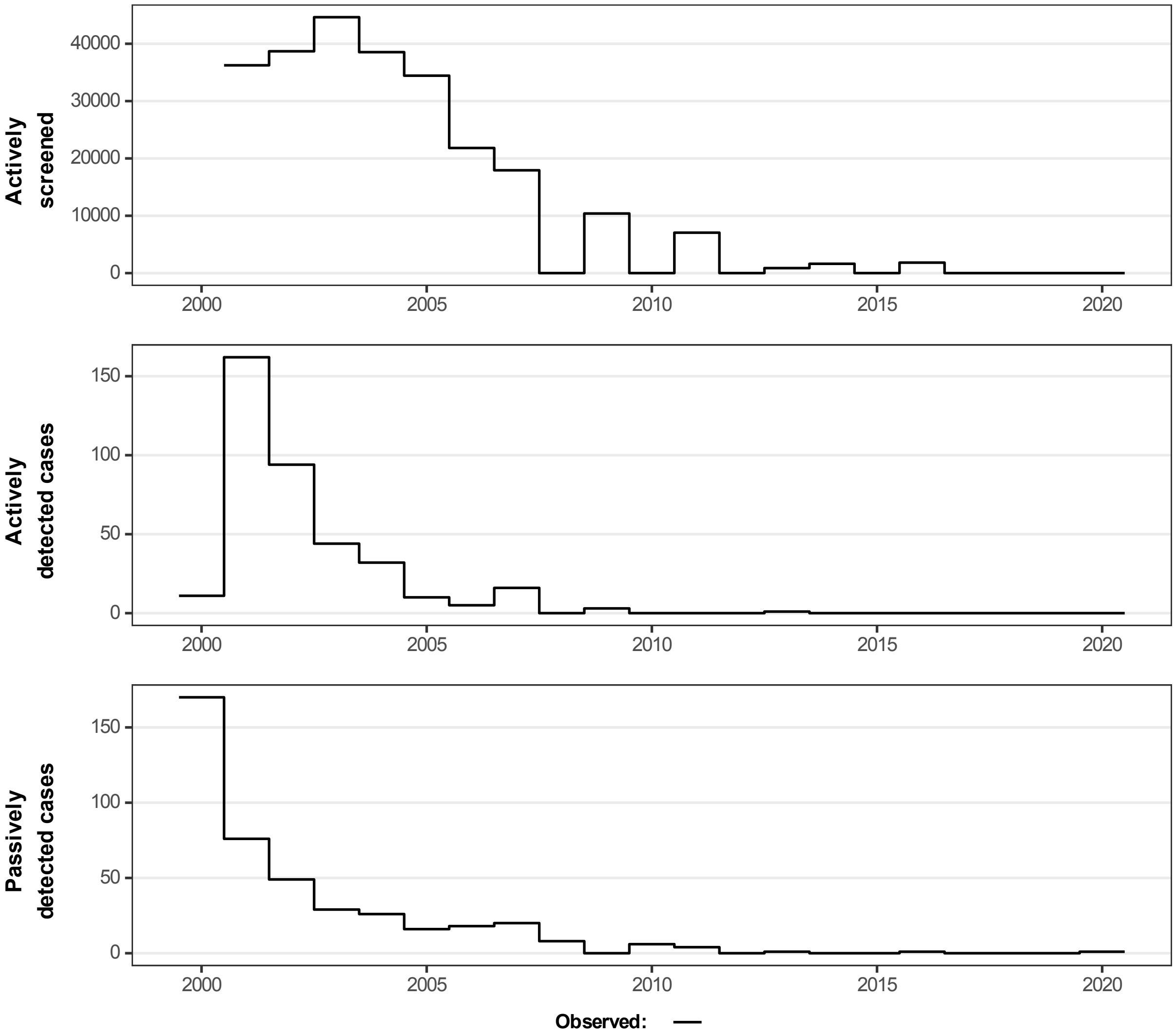


Fig 3: Aggregated data used for model fitting in Budjala.

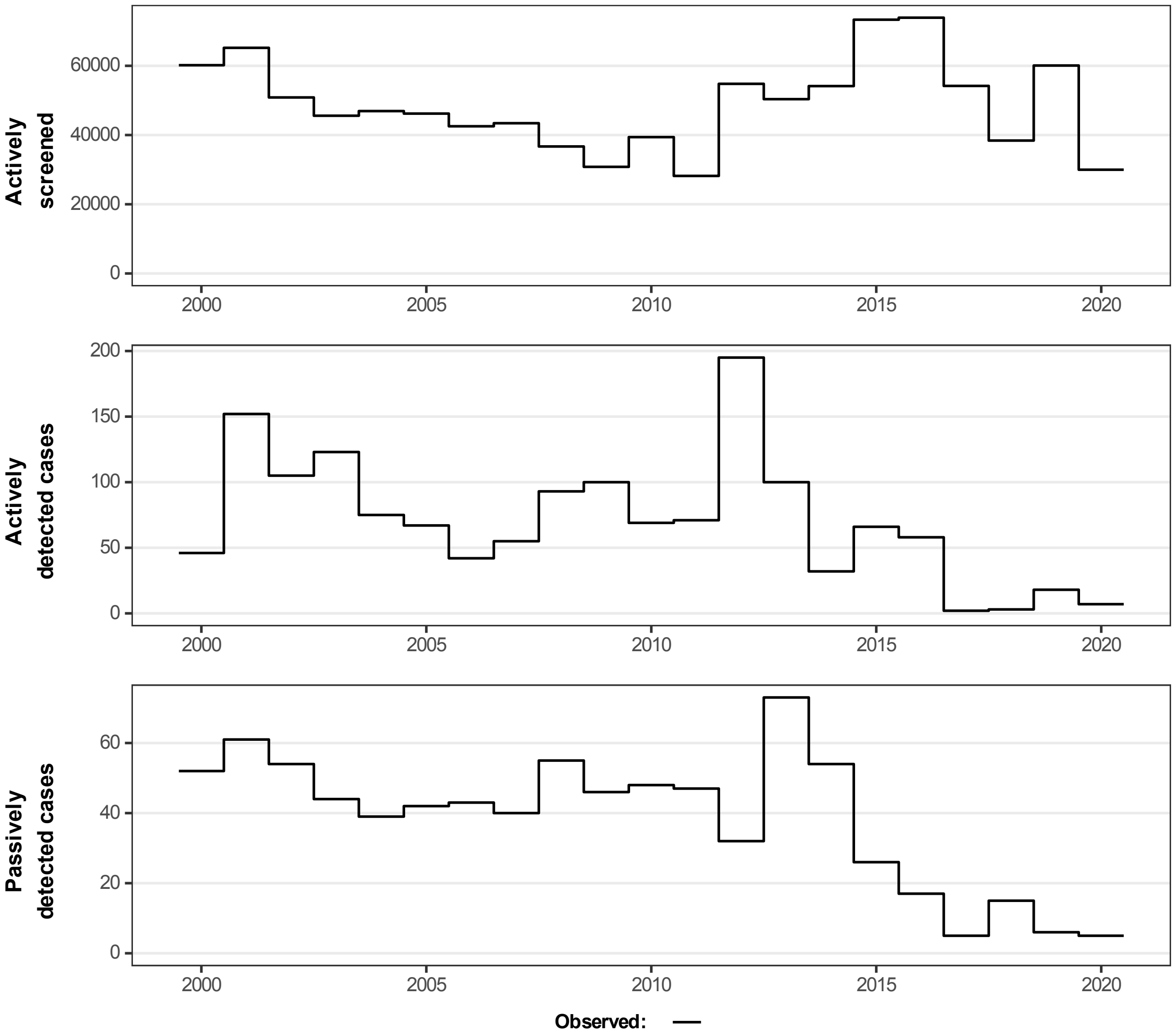


Fig 4: Aggregated data used for model fitting in Bagata.

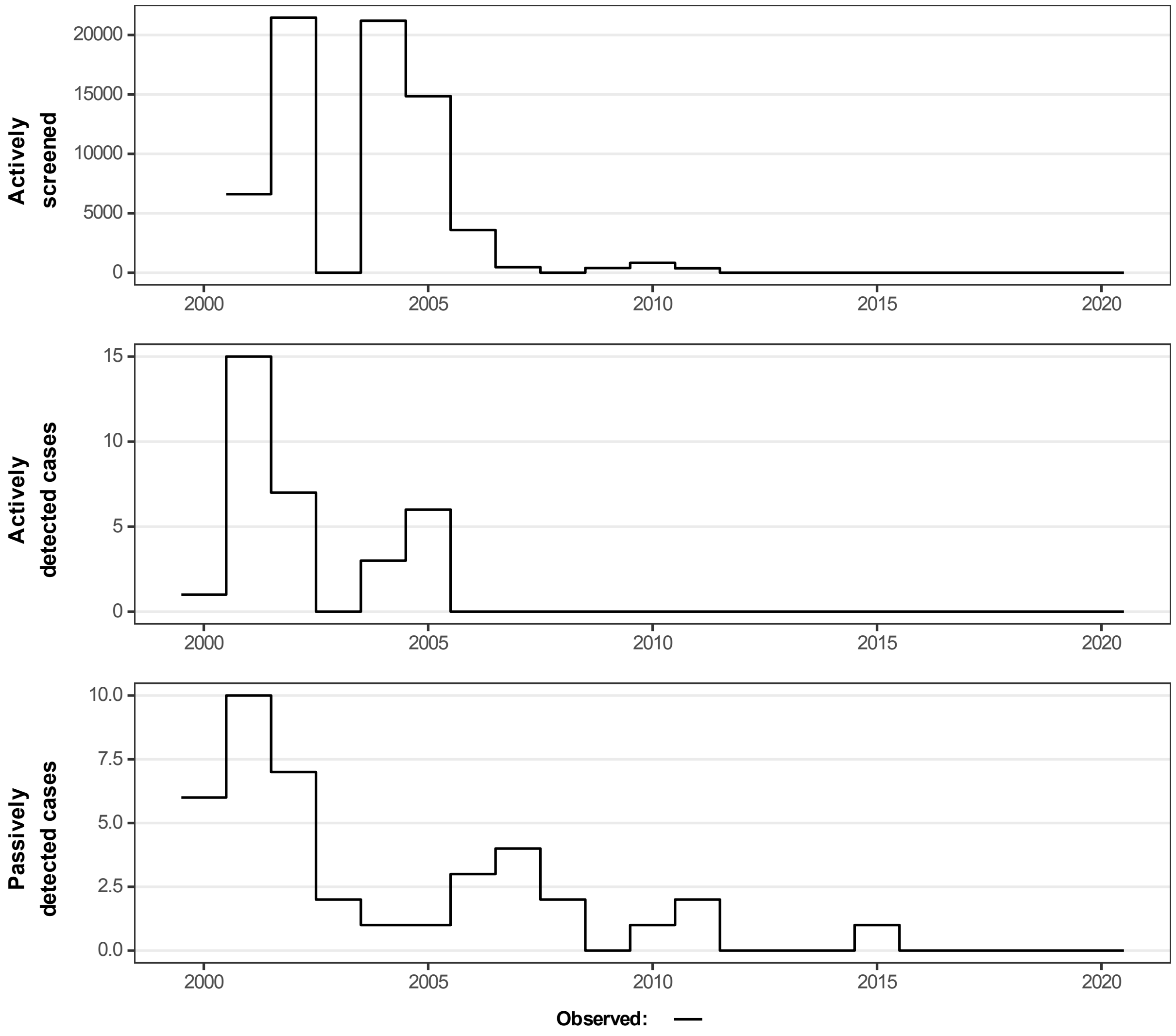


Fig 5: Aggregated data used for model fitting in Mbaya.

#### Model variants

The compartmental gHAT infection model (Fig 6) and its equations (Eqn S.0.1 have been presented elsewhere previously [9, 10, 1] for either the baseline model and model with animal transmission or the asymptomatic model, but not all together. Descriptions of model parameters can be found in Tables A and B.

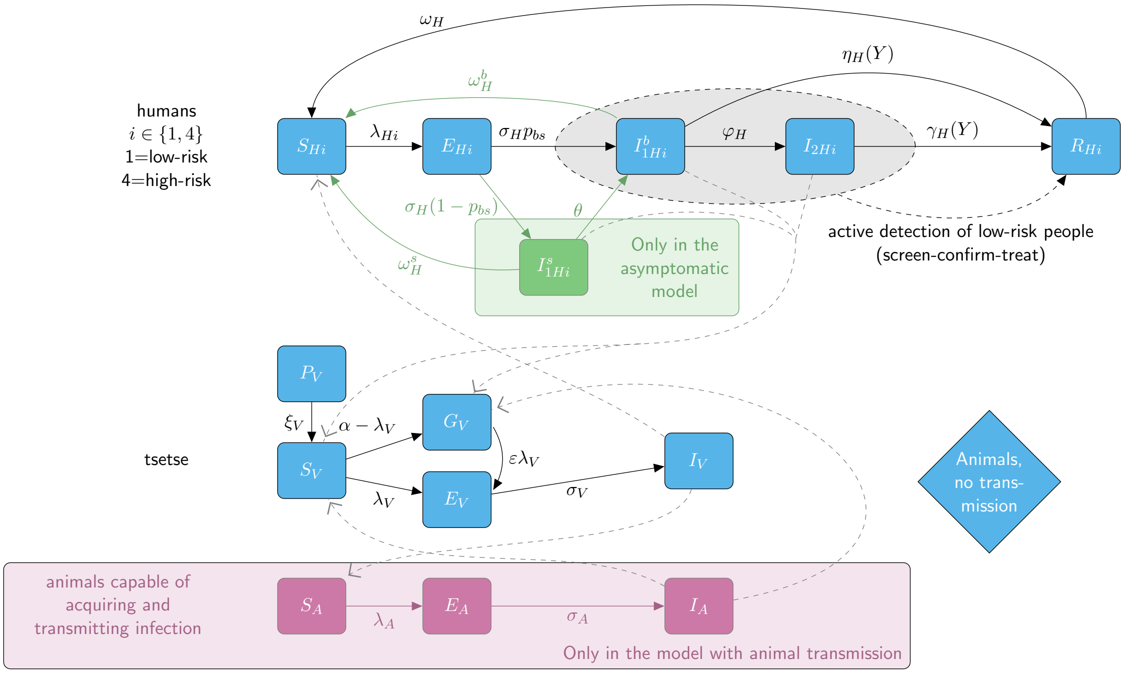


Fig 6: Compartmental model schematic using Greek notation for rates and capital Roman characters for different host and vector infection states. Blue components form the baseline model and are also included in the other two model variants. The pink boxes and arrows are only found in the animal model and the green box and arrows are only in the asymptomatic model variant. Transmission pathways are shown as dashed grey lines. Births and deaths are included but are not shown here to aid readability. The grey oval and dashed black lines indicate infection classes assumed to be detectable using a traditional screen-confirm-treat approach in active screening (although some infections still may be missed due to imperfect diagnostic sensitivity).

$$
\text{Humans}\begin{cases}
\frac{\mathrm{d}S_{Hi}}{\mathrm{d}t} &= \mu_H N_{Hi} + \omega_H^s I_{1Hi}^s + \omega_H^b I_{1Hi}^b + \omega_H R_{Hi} - \alpha m_{\text{eff}} f_i \frac{S_{Hi}}{N_{Hi}} I_V - \mu_H S_{Hi} \\
\frac{\mathrm{d}E_{Hi}}{\mathrm{d}t} &= \alpha m_{\text{eff}} f_i \frac{S_{Hi}}{N_{Hi}} I_V - (\sigma_H + \mu_H) E_{Hi} \\
\frac{\mathrm{d}I_{1Hi}^s}{\mathrm{d}t} &= \sigma_H (1 - p_{bs}) E_{Hi} - (\theta + \omega_H^s + \mu_H) I_{1Hi}^s \\
\frac{\mathrm{d}I_{1Hi}^b}{\mathrm{d}t} &= \sigma_H p_{bs} E_{Hi} - (\varphi_H + \omega_H^b + \eta_H(Y) + \mu_H) I_{1Hi}^b \\
\frac{\mathrm{d}I_{2Hi}}{\mathrm{d}t} &= \varphi_H I_{1Hi}^b - (\gamma_H(Y) + \mu_H) I_{2Hi} \\
\frac{\mathrm{d}R_{Hi}}{\mathrm{d}t} &= \eta_H(Y) I_{1Hi}^b + \gamma_H(Y) I_{2Hi} - (\omega_H + \mu_H) R_{Hi}
\end{cases}
$$

$$
\text{Animals}\begin{cases}
\frac{\mathrm{d}S_A}{\mathrm{d}t} &= \mu_A N_A - \alpha m_{\text{eff}} f_A \frac{S_A}{N_A} I_V - \mu_A S_A \\
\frac{\mathrm{d}E_A}{\mathrm{d}t} &= \alpha m_{\text{eff}} f_A \frac{S_A}{N_A} I_V - (\sigma_A + \mu_A) E_A \\
\frac{\mathrm{d}I_A}{\mathrm{d}t} &= \sigma_A E_A - \mu_A I_A
\end{cases}
$$

$$
\text{Tsetse}\begin{cases}
\frac{\mathrm{d}P_V}{\mathrm{d}t} &= B_V N_H - (\xi_V + \frac{P_V}{K}) P_V \\
\frac{\mathrm{d}S_V}{\mathrm{d}t} &= \xi_V \mathbb{P}(\text{pupating}) P_V - \alpha S_V - \mu_V S_V \\
\frac{\mathrm{d}E_{1V}}{\mathrm{d}t} &= \alpha(1 - f_T(t)) p_V \big( \sum_i f_i \frac{(I_{1Hi}^b + x I_{1Hi}^s + I_{2Hi})}{N_{Hi}} + f_A \frac{I_A}{N_A} \big)(S_V + \varepsilon G_V) \\
&\quad -(3\sigma_V + \mu_V + \alpha f_T(t)) E_{1V} \\
\frac{\mathrm{d}E_{2V}}{\mathrm{d}t} &= 3\sigma_V E_{1V} - (3\sigma_V + \mu_V + \alpha f_T(t)) E_{2V} \\
\frac{\mathrm{d}E_{3V}}{\mathrm{d}t} &= 3\sigma_V E_{2V} - (3\sigma_V + \mu_V + \alpha f_T(t)) E_{3V} \\
\frac{\mathrm{d}I_V}{\mathrm{d}t} &= 3\sigma_V E_{3V} - (\mu_V + \alpha f_T(t)) I_V \\
\frac{\mathrm{d}G_V}{\mathrm{d}t} &= \alpha(1 - f_T(t)) \Big( 1 - p_V \big( \sum_i f_i \frac{(I_{1Hi}^b + x I_{1Hi}^s + I_{2Hi})}{N_{Hi}} + f_A \frac{I_A}{N_A} \big) \Big) S_V \\
&\quad -\alpha \Big( f_T(t) + (1 - f_T(t)) p_V \varepsilon \big( \sum_i f_i \frac{(I_{1Hi}^b + x I_{1Hi}^s + I_{2Hi})}{N_{Hi}} + f_A \frac{I_A}{N_A} \big) \Big) G_V \\
&\quad -\mu_V G_V
\end{cases}
\tag{S.0.1}
$$

$$
f_T(t) = f_{\text{max}} \left( 1 - \frac{1}{1 + \exp(-0.068(\text{mod}(t, 182.5) - 127.75))} \right) \tag{S.0.2}
$$

#### Improvements to active screening diagnostic specificity

The active screening diagnostic algorithm characteristics are dependent on the series of tests performed before someone can be considered a confirmed case and treatment can be administered. Except for the modelled "screen-and-treat" (S&T) strategy, all serological suspects are assumed to need a parasitological confirmation test to receive treatment due to current drug administration guidelines for either pentamidine, NECT or fexinidazole [12].

Parasitological confirmation is made through microscopy, requiring trained personnel to make the diagnosis. Visualisation of the parasite in this manner should theoretically enable perfect specificity of the confirmation test (and therefore of the full diagnostic algorithm), however, the increasing rarity of the infection means that most technicians have limited recent experience in making confirmations. In our fitting procedure, we estimate the health-zone-specific active screening specificity based on historical case reporting between 2000–

Table A: **Model parameterisation (fixed parameters).** Notation, a brief description, and the values used for fixed parameters.

| Model | Notation | Description | Value | |
|---|---|---|---|---|
| All | $N_H$ | Total human population size in 2015 | Fixed for each health zone | [16] |
| All | $\mu_H$ | Natural human mortality rate | 5.4795×$10^{-5}$ days$^{-1}$ | [22] |
| All | $B_H$ | Total human birth rate | = $\mu_H N_H$ | |
| All | $\sigma_H$ | Human incubation rate | 0.0833 days$^{-1}$ | [21] |
| All | $\varphi_H$ | Stage 1 to 2 progression rate | 0.0019 days$^{-1}$ | [5, 7] |
| All | $\omega_H$ | Recovery rate or waning-immunity rate | 0.006 days$^{-1}$ | [15] |
| All | Sens(AS) | Active screening algorithm diagnostic sensitivity | 0.91 | [6] |
| All | Sens(TT) | Initial screening diagnostic sensitivity | 0.95 | Assumed |
| All | Spec(TT) | Initial screening diagnostic specificity | 0.995 | Assumed |
| All | $B_V$ | Tsetse birth rate (per capita rate of depositing new pupae) | 0.0505 days$^{-1}$ | [20] |
| All | $\xi_V$ | Rate of pupal development to adult flies | 0.037 days $^{-1}$ | |
| All | $K$ | Pupal carrying capacity | = $111.09N_H$ | [20] |
| All | $\mathbb{P}$(pupating) | Probability of a pupa surviving to emerge as an adult fly | 0.75 | |
| All | $\mu_V$ | Tsetse mortality rate | 0.03 days$^{-1}$ | [21] |
| All | $\sigma_V$ | Tsetse incubation rate | 0.034 days$^{-1}$ | [11, 17] |
| All | $\alpha$ | Tsetse bite rate | 0.333 days$^{-1}$ | [24] |
| All | $p_V$ | Probability of tsetse infection per single infective bite | 0.065 | [21] |
| All | $\varepsilon$ | Reduced susceptibility factor for non-teneral (previously fed) flies | 0.05 | [19] |
| All | $f_H$ | Proportion of blood-meals on humans | 0.09 | [8] |
| All | $\text{disp}_{\text{act}}$ | Overdispersion parameter for active detection | $4 \times 10^{-4}$ | – |
| All | $\text{disp}_{\text{pass}}$ | Overdispersion parameter for passive detection | $2.8 \times 10^{-5}$ | – |
| All | $f$max | Probability of a tsetse contacting a Tiny Target and dying per blood meal to yield a 80% population reduction after one year | 0.0525 | – |
| Animal | $\mu_A$ | Natural animal mortality rate | 0.0014 days$^{-1}$ | Assumed |
| Animal | $\sigma_A$ | Animal incubation rate | 0.0833 days$^{-1}$ | [21] |

The value of $B_V$ was chosen to maintain constant population size in the absence of vector control interventions. The value of $K$ was chosen to reflect a plausible bounce-back rate.

Table B: **Model parameterisation (fitted parameters).** Notation, brief description, and information on the prior distributions for fitted parameters.

| Notation | Description | Prior distribution* | Percentiles of prior distribution [2.5, 50 & 97.5%] | Unit |
|---|---|---|---|---|
| $R_0$ | Basic reproduction number (NGM approach) | $1 + \text{Exp}(10)$ | [1.003, 1.069, 1.369] | - |
| $r$ | Relative bites taken on high-risk humans | $1 + \Gamma(3.68, 1.09)$ | [2.015, 4.654, 10.028] | - |
| $k_1$ | Proportion of low-risk people | $\text{B}(16.97, 3.23)$ | [0.6564, 0.8514, 0.9609] | - |
| $\eta_H^{\text{post}}$ | Treatment rate from stage 1, 1998 onwards | $\Gamma(3.54, 5.32 \times 10^{-5})^\dagger$ <br> $\Gamma(4.92, 4.51 \times 10^{-5})^\ddagger$ | [4.59, 17.1, 42.9]$\times 10^{-5}$ <br> $[7.12, 20.7, 45.7] \times 10^{-5}$ | days$^{-1}$ |
| $\gamma_H^{\text{post}}$ | Combined treatment and disease-induced death rate from stage 2, 1998 onwards | $\Gamma(2.45, 0.00192)$ | [7.59, 40.7, 121]$\times 10^{-4}$ | days$^{-1}$ |
| $b_{\gamma_H^{\text{pre}}}$ | Relative treatment/death rate from stage 2 factor, pre-1998 | $\text{B}(1, 1)$ | [0.025, 0.500, 0.975] | - |
| Spec(AS) | Active screening diagnostic specificity | $0.998 + (1 - 0.998)\,\text{B}(7.23, 2.41)$ | [0.9989, 0.9995, 0.9999] | - |
| $u$ | Proportion of stage 2 passive cases reported | $\text{B}(20, 40)$ | [0.2208, 0.3315, 0.4564] | - |
| $d_{\text{change}}$ | Midpoint year for passive improvement | $2000 + (2020 - 2000)\,\text{B}(5, 6)^\dagger$ <br> $2000 + (2020 - 2000)\,\text{B}(2.79, 23.1)^\ddagger$ | [2003.2, 2007.7, 2012.5] <br> $[2000.4, 2002, 2005]$ | Year |
| $\eta_{H_{\text{amp}}}$ | Relative improvement in passive stage 1 detection rate | $\Gamma(2.013, 1.049)^\dagger$ <br> $\Gamma(1, 2.17)^\ddagger$ | [0.258, 1.775, 5.870] <br> $[0.055, 1.510, 8.010]$ | - |
| $\gamma_{H_{\text{amp}}}$ | Relative improvement in passive stage 2 detection rate | $\Gamma(1.001, 5)^\dagger$ <br> $\Gamma(1, 1.0014)^\ddagger$ | [0.127, 3.471, 18.455] <br> $[0.0254, 0.6943.69]$ | - |
| $d_{\text{steep}}$ | Speed of improvement in passive detection rate | $\Gamma(39.57, 0.0270)^\dagger$ <br> $\Gamma(15.7, 0.51)^\ddagger$ | [0.761, 1.058, 1.424] <br> $[4.55, 7.84, 12.4]$ | years$^{-1}$ |
| **Parameters specific to the animal model...** | | | | |
| $f_A$ | Proportion of blood meals on reservoir animals | $\text{B}(1.3, 1.3)$ | [0.046, 0.500, 0.954] | - |
| $k_A$ | Relative size of animal reservoir population | $\Gamma(1.26, 19.3)$ | [1.18, 18.3, 19.3] | - |
| **Parameters specific to the asymptomatic model...** | | | | |
| $p_{bs}$ | Proportion of human exposures resulting in initial blood infection | $\text{B}(10, 4)$ | [0.462, 0.725, 0.909] | - |
| $\omega_H^b$ | Self-cure rate for stage 1 blood infections | $\Gamma(4, 1.5 \times 10^{-6})$ | [0.163, 0.551, 1.32] $\times 10^{-5}$ | days$^{-1}$ |
| $\omega_H^s$ | Self-cure rate for skin-only infections | $\Gamma(4, 10^{-4})$ | [1.09, 3.67, 8.77] $\times 10^{-4}$ | days$^{-1}$ |
| $\theta$ | Transition rate from skin-only to blood infection | $\Gamma(4, 10^{-5})$ | [1.09, 3.67, 8.77]$\times 10^{-5}$ | days$^{-1}$ |
| $x$ | Relative infectiousness of skin-only infection compared to a blood infection | $\text{B}(3, 3)$ | [0.147, 0.500, 0.853] | days$^{-1}$ |

*Where Exp(.), $\Gamma$(.) and B(.) are the exponential, gamma (parameterised with shape and scale) and beta distributions, respectively. †Prior used in Bagata and Mosango health zones (former province of Bandundu) ‡Prior used in Bominenge, Budjala and Mbaya health zones (former province of Equateur).

2016. Whilst specificity is assumed to be very high (>99.8%) the high coverage of active screening in many health zones means that it is very likely that some false positives were incorrectly confirmed.

To combat the challenge of accurately making confirmations in the field in the context of falling cases, mobile teams in former Bandundu province now also utilise video confirmation which records the parasites for quality assurance by a second person. This technology was rolled out in Mosango health zone in 2015 and in most other health zones in the former Bandundu province in 2018, therefore we increased our modelled active screening algorithm specificity to 100% in the corresponding year.

In former Equateur province video tablets are not being used by active screening teams. However, we do assume that if large screenings take place and case reporting is low, there would be more careful follow-up of case confirmations. We therefore perfect active screening algorithm specificity from 2024 (the beginning of our projections).

#### Improvements to passive screening

We assume, based on a substantial shift in the provincial-level stage 1 to stage 2 case ratio between 2000–2016 [13], that there is an improvement in passive case detection rates in the former Bandundu province (including Mosango and Bagata health zones) [4]. An improvement to passive screening was also fitted for Bominenge, Budjala and Mbaya health zones in the former province of Equateur but using different priors on the parameters involved. The priors were based on modelling of former-province level data for 2000–2012, augmented with WHO HAT Atlas data to make a 2000–2016 data set. An arbitrary amount of additional variation was incorporated into the priors. Note that the province-level data contained staging which is not available in this period in the WHO HAT Atlas but which was expected to be informative for this process.

$$\eta_H(Y) = \eta_H^{\text{post}} \left[1 + \frac{\eta_{H_{\text{amp}}}}{1 + \exp\left(-d_{\text{steep}}(Y - d_{\text{change}})\right)}\right] \tag{S.0.3}$$

$$\gamma_H(Y) = \gamma_H^{\text{post}} \left[1 + \frac{\gamma_{H_{\text{amp}}}}{1 + \exp\left(-d_{\text{steep}}(Y - d_{\text{change}})\right)}\right] \tag{S.0.4}$$

We assume that all stage 1 cases are reported, but that some of the exits from stage 2 are due to death from gHAT disease outside healthcare. In 1998 the reporting probability for an exit from stage 2 is given by $u$, however as the exit rate from stage 2 increases this reporting probability does not stay constant, but increases (proportionally more people would be detected and treated with higher exit rates). When we compute reporting rates from stage 2 we therefore use the following:

$$\text{Death rate} = (1 - u)\gamma_H^{\text{post}} \tag{S.0.5}$$

$$\text{Stage 2 reporting incidence} = (\gamma_H(Y) - \text{Death rate})(I_{2H1} + I_{2H4}) \tag{S.0.6}$$

#### Vector Control

The function which describes the probability of a host-seeking tsetse both hitting a Tiny Target and dying as a result, $f_T$, is time-dependent ($t$, in days) from when the targets were first deployed:

$$f_T(t) = f_{\text{max}} \left(1 - \frac{1}{1 + \exp(-0.068(\text{mod}(t, 182.5) - 127.75))}\right) \tag{S.0.7}$$

and $f_{\text{max}}$ – the maximum daily probability of contacting a Tiny Target and dying as a result. $f_T$ modifies all the bite rates $\alpha$ in our tsetse equations to produce an additional Tiny-Target-induced mortality for tsetse. $f_{\text{max}}$ is chosen such that the tsetse population after one year is at the observed/assumed percentage reduction. For this model of $f_T$ this is given by $f_{\text{max}} = 0.0525$ for an assumed annual tsetse population density reduction of 80%. The value 182.5 reflects twice-yearly deployments of Tiny Targets, as used in DRC [23].

#### Fitting procedure

##### Model initialisation

For each model for a particular parameter set, we analytically compute pre-1998 initial conditions assuming that the system was at endemic equilibrium. In 1998 we assume that passive detection rates increased with the introduction of the CATT test and that active screening began at the same level as seen in the data for 2000.

##### Likelihood and Markov chain Monte Carlo

We describe the fitting procedure for all models below, with text adapted from our previous fitting publications [9, 10].

Twelve parameters; $R_0$, $r$, $\eta_H$, $\gamma_H$, $b_{\gamma_{H0}}$, $k_1$, $u$, $\mathrm{Spec}$, $d_{\mathrm{change}}$, $\eta_{H_{\mathrm{amp}}}$, $\gamma_{H_{\mathrm{amp}}}$, and $d_{\mathrm{steep}}$ were fitted in all health zones for all models. A further two parameters; $k_A$ and $f_A$, were fitted for the model with animal transmission and five parameters: $x$, $\theta$, $\omega_H^s$, $\omega_H^b$ and $p_{bs}$ were fitted for the asymptomatic model.

For fitting the model variants to case data we transform model ODE solutions (for Equations S.0.1) into annual case reporting denoted $A_{M1}$, $A_{M2}$, for active stage 1 and stage 2 and $P_{M1}$, $P_{M2}$, for passive stage 1 and 2. Since we always know the stage (1 or 2) in the model simulations there is no requirement for a "U" (unknown stage) category for the model. These are computed using solutions to the ODEs for the given set of parameters aggregated across a year.

The new annual reported case incidence is either by passive detection from stage 1 for the year $Y$:

$$P_{M1}(Y) = \int_Y^{Y+1} \eta_H(Y)\big(I_{1H1}(t) + I_{1H4}(t)\big)\,\mathrm{d}t, \tag{S.0.8}$$

passive detection from stage 2

$$P_{M2}(Y) = \int_Y^{Y+1} (\gamma_H(Y) - \text{Death rate})\big((I_{2H1}(t) + I_{2H4}(t)\big)\,\mathrm{d}t, \tag{S.0.9}$$

or by active screening from the low-risk ($H1$) group in year $Y$

$$A_{M1}(Y) = z(Y)\mathrm{Sens}I_{1H1}(Y) + z(Y)(1 - \mathrm{Spec})\big(k_1 N_H - I_{1H1}(Y) - I_{2H1}(Y)\big) \tag{S.0.10}$$

and

$$A_{M2}(Y) = z(Y) \times \mathrm{Sens} \times I_{2H1}(Y) \tag{S.0.11}$$

with variable active screening coverage by year, $z(Y)$ and fixed diagnostic sensitivity. $A_{M1}$ also contains any false positives that may be been incorrectly identified from non-infected people based on the high but imperfect specificity of the active screening algorithm. We assume in DRC that all false positives would be assigned as stage 1.

The log-likelihood function used in the adaptive Metropolis-Hastings MCMC contained two terms in each year for which reported case numbers were available for each source of reported cases (active or passive screening). These were:

- a beta-binomial probability that the total number of cases reported in that year for that source came from the available population (either the reported number of people actively screened for active screening or the health zone population for passive screening) with probability calculated from solving the ODE for the current set of parameters, and
- a binomial probability that the reported stage 1 cases came from the total number of reported staged cases where the probability parameter again comes from the solution of the ODE. In many years staging is unknown so this part of the log-likelihood will return zero and not contribute to our calculation. In some years, we only partially know staging information.

This formulation allowed over-dispersion in the observed cases to be included, via the beta-binomial distribution, and any proportion of cases with reported disease stage to be appropriately accounted for (assuming that the reporting of staging information is independent of the disease stage). The log-likelihood function was as follows:

$$
\begin{aligned}
LL(\theta|x) \;\; = \;\; & \log(P(x|\theta)) \\
\propto \;\; & \textstyle\sum_{i=2000}^{2016} \Bigg( \log \left[ \text{BetaBin} \left( A_{D1}(i) + A_{D2}(i) + A_{DU}(i); z(i), \frac{A_{M1}(i) + A_{M2}(i)}{z(i)}, \text{disp}_{\text{act}} \right) \right] \\
& + \log \left[ \text{Bin} \left( A_{D1}(i); A_{D1}(i) + A_{D2}(i), \frac{A_{M1}(i)}{A_{M1}(i) + A_{M2}(i)} \right) \right] \\
& + \log \left[ \text{BetaBin} \left( P_{D1}(i) + P_{D2}(i) + P_{DU}(i); N_H, \frac{P_{M1}(i) + P_{M2}(i)}{N_H}, \text{disp}_{\text{pass}} \right) \right] \\
& + \log \left[ \text{Bin} \left( P_{D1}(i); P_{D1}(i) + P_{D2}(i), \frac{P_{M1}(i)}{P_{M1}(i) + P_{M2}(i)} \right) \right] \Bigg)
\end{aligned}
\tag{S.0.12}
$$

The model takes parameterisation $\theta$, $x$ is the data, $P_{Dj}(i)$ and $A_{Dj}(i)$ are the number of cases detected by passive or active screening (of stage $j$, which may be 1, 2 or unknown, $U$) in year $i$ of the data. $P_{Mj}(i)$ and $A_{Mj}(i)$ are the number of cases detected by passive or active screening (of stage $j$) in year $i$ of the model, and $z(i)$ is the number of people screened in year $i$. $\text{BetaBin}(m; n, p, \rho)$ gives the probability of obtaining $m$ successes out of $n$ trials with probability $p$ and overdispersion parameter $\rho$. The overdispersion accounts for a larger variance than under the binomial. The probability density function of this distribution is given by:

$$
\text{BetaBin}(m; n, p, \rho) = \frac{\Gamma(n+1)\Gamma(m+a)\Gamma(n-m+b)\Gamma(a+b)}{\Gamma(n-m+1)\Gamma(n+a+b)\Gamma(a)\Gamma(b)} \tag{S.0.13}
$$

where $a = p(1/\rho - 1)$ and $b = a(1-p)/p$.

#### Sequential MCMC fitting of the asymptomatic model to health zones, with Bayesian updating of priors

There are five parameters which are unique to our model with transmission of HAT to and from a reservoir of asymptomatic human infections:

1. the proportion of human exposures resulting in initial blood infection ($p_{bs}$);
2. the self-cure rate for stage 1 blood infections ();
3. the self-cure rate for skin-only infections;
4. the transition rate from skin-only to blood infection; and
5. the relative infectiousness of skin-only infection compared to a blood infection.

We regard these parameters as being innate, biological values and assume that they should be invariant across geographies. This is in contrast to the parameters which are unique to the model with transmission to and from a non-specific animal source, which is liable to vary as a result of differences in the animal species, variability in the amount of animal and tsetse habitats and interactions between these hosts and tsetse.

The best way to fit the asymptomatic model would be to consider all health zones together in a hierarchical model, however, this would be computationally very demanding especially if using this method to fit the model to many more regions e.g. all endemic health zones in the whole of the DRC (>150) or all endemic health areas (regions of around 10,000 people) of the DRC (>1,000). We have therefore opted to perform Bayesian updating of priors such that posterior parameter distributions from health zones that are informative for these parameters provide the prior distributions used for the asymptomatic model-specific parameters in other health zones.

The procedure used is as follows:

1. All health zones were analysed with the same univariate prior distributions (Table B), five MCMC chains were run in each analysis with 1,000 posterior samples taken from each chain, to give 5,000 posterior samples overall with a minimum effective sample size of 2,500.

2. The Kullback-Liebler divergence of the realised marginal posterior distributions from the univariate, independent prior distributions, $\mathrm{KL}(X|Y)$, were calculated and used to assign a rank to the health zones $j = 1, \ldots, 5$.

3. Health zones $j = 2, \ldots, 5$ were re-analysed using a prior based on the posterior distribution of the five asymptomatic specific parameters from the analysis of health zone $j - 1$. A Gaussian mixture model (GMM, a weighted mixture of multivariate normal distributions) was fitted to the samples from the posterior distribution from the analysis of health zone $j - 1$.

##### GMM fitting

For the MCMC analysis of health zone $j$, the 5,000 posterior samples from the analysis of HZ $j-1$ were read in. Each of the five asymptomatic model-specific parameters was first transformed to a $(-\infty, +\infty)$ scale and then to have a mean of zero and a standard deviation of one. The transformation to convert to a $(-\infty, +\infty)$ scale and the centring and scaling values are retained to create the Jacobian for the scaled GMM prior.

A Gaussian Mixture Model was fitted to the transformed parameters using the MATLAB routine `fitgmdist`. Akaike's Information Criterion (AIC) was used to select the number of Gaussian distributions to include in the mixture.

A MATLAB structure was created containing the final GMM, identifiers for the parameters, and transformation information and this was saved to act as the multivariate five-parameter prior for the asymptomatic model-specific parameters in the analysis of the $j^{\mathrm{th}}$ health zone.

### Additional results

#### Model fitting – posteriors

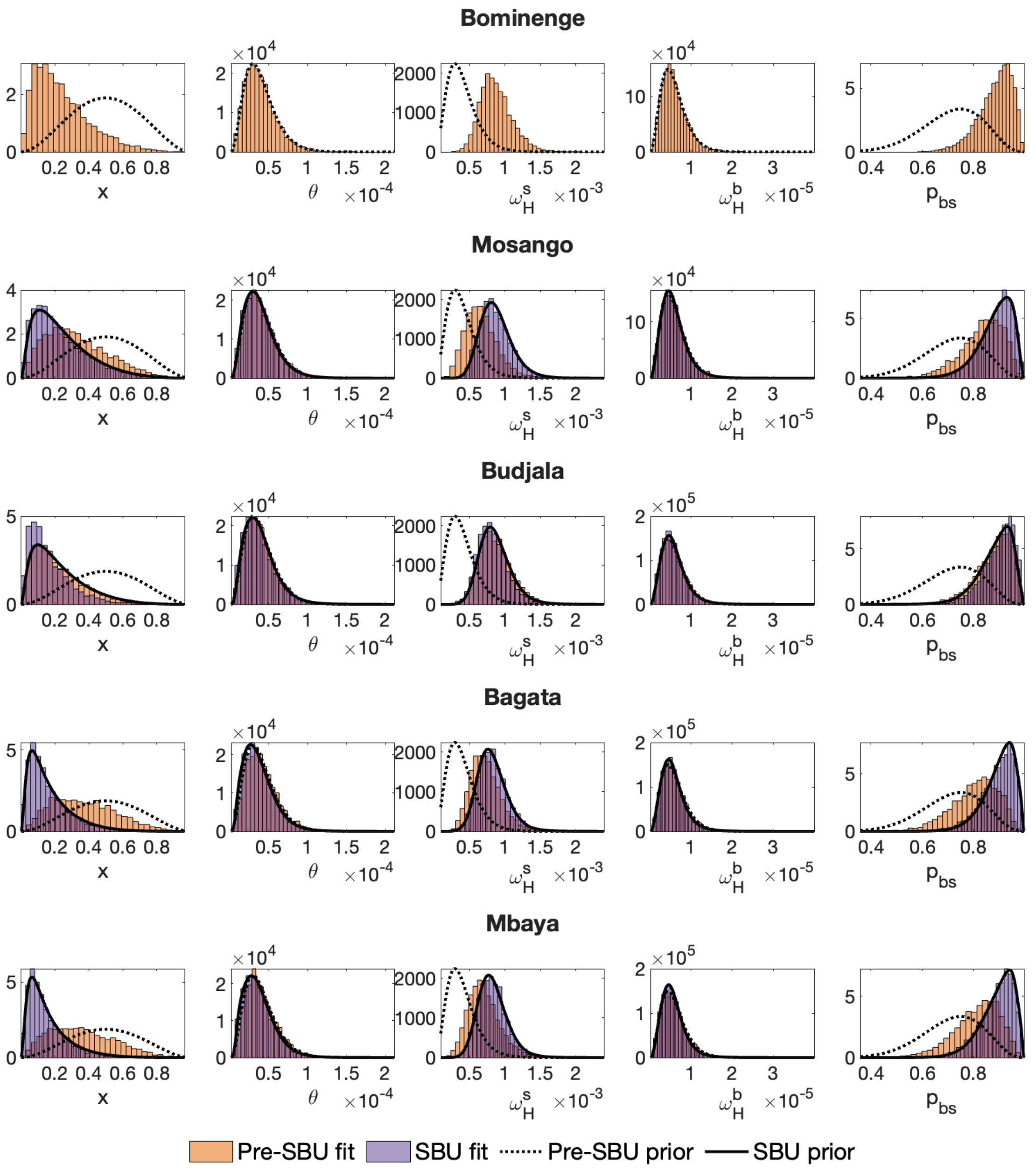


Fig 7: **Posterior distributions of asymptomatic model specific parameters before and after sequential Bayesian (SBU).** Histograms of posterior samples from before and with SBU updating and their prior distributions. Rows are ordered by total Kullback-Liebler divergence, hence Bominenge only has pre-SBU outputs (see Table C).

Table C: Approximate Kullback-Liebler divergence between the prior and posterior distributions for each of the parameters specific to the asymptomatic human transmission model and their total, which was used to decide the order of running the sequential Bayesian updating.

| Health zone | Kullback-Liebler divergence | | | | | |
|---|---|---|---|---|---|---|
| | $x$ | $\theta$ | $\omega_H^s$ | $\omega_H^b$ | $p_{bs}$ | Total |
| Bominenge | 7.0 | 5.7 | 7.0 | 5.3 | 12.7 | 37.7 |
| Mosango | 6.3 | 9.6 | 11.4 | 4.7 | 3.3 | 35.3 |
| Budjala | 5.8 | 4.7 | 8.1 | 5.7 | 10.7 | 35.0 |
| Bagata | 2.5 | 5.5 | 7.6 | 5.6 | 3.3 | 24.6 |
| Mbaya | 2.0 | 4.4 | 7.5 | 4.3 | 3.4 | 21.8 |

Table D: Relative model evidence (%) after sequential Bayesian updating of the asymptomatic human transmission model – weights in the ensemble model. See main text Figure 5.

| Health zone | Baseline | Animal | Asymptomatic |
|---|---|---|---|
| Bominenge | 96.1 | 3.8 | 0.1 |
| Mosango | 45.9 | 7.1 | 47.0 |
| Budjala | 59.2 | 2.8 | 38.0 |
| Bagata | 38.4 | 12.2 | 49.4 |
| Mbaya | 43.0 | 8.6 | 48.4 |

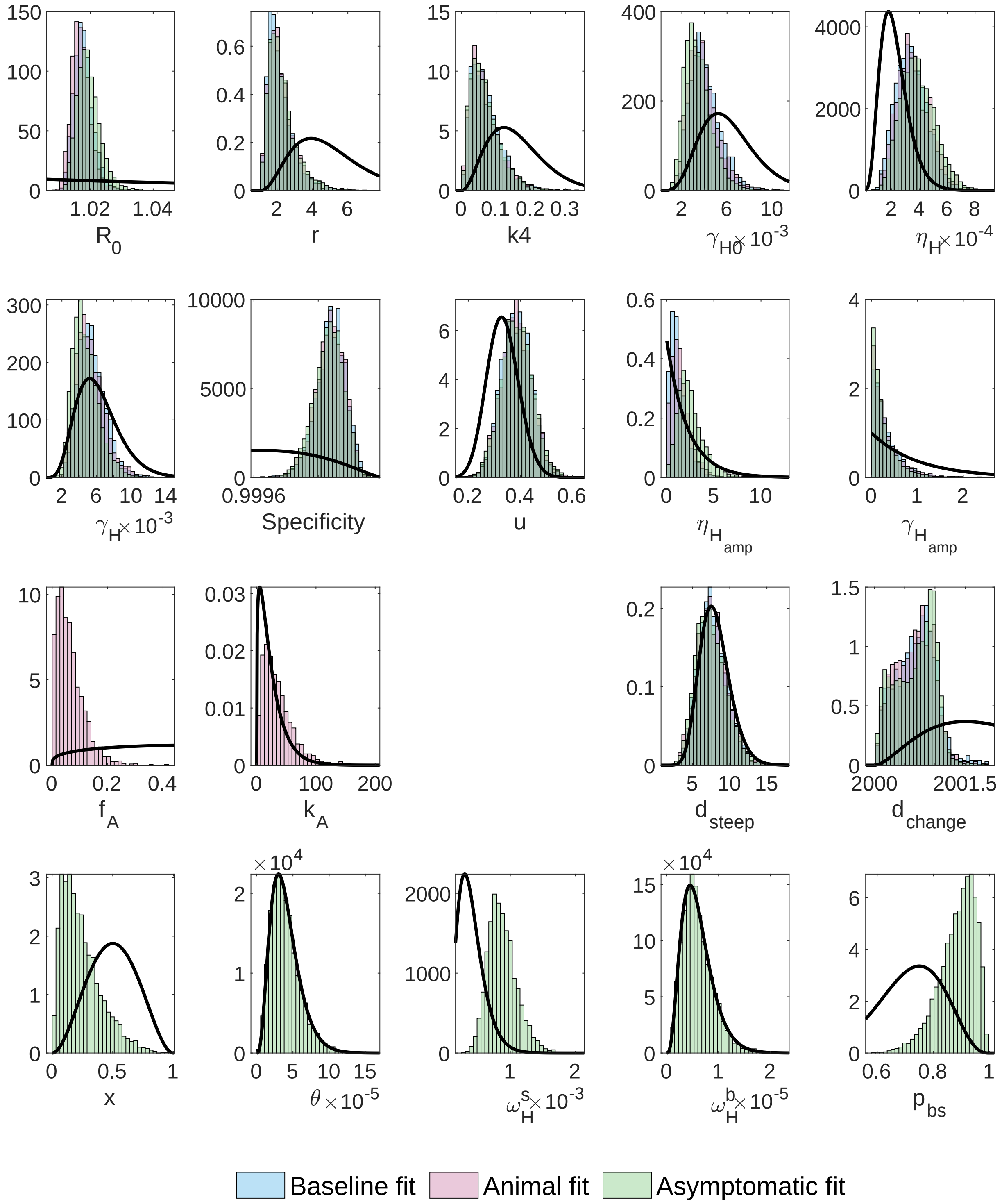


Fig 8: **Posterior parameter distributions for Bominenge (after sequential Bayesian (SBU))** using the deterministic model for fitting each of the three model variants to case data. There are two fitted parameters which only appear in the animal model, and five which only appear in the asymptomatic model

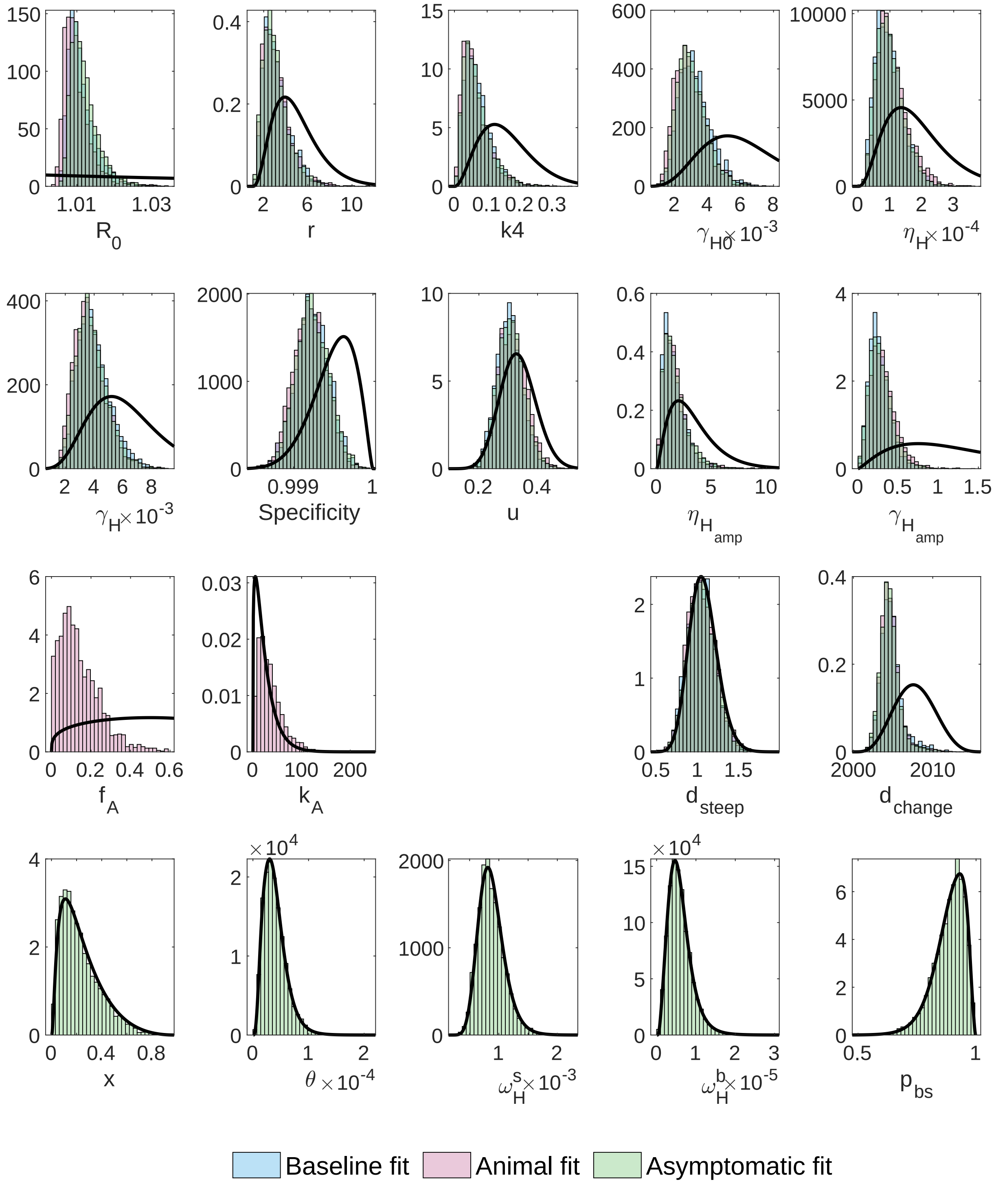


Fig 9: **Posterior parameter distributions for Mosango (after sequential Bayesian (SBU))** using the deterministic model for fitting each of the three model variants to case data. There are two fitted parameters which only appear in the animal model, and five which only appear in the asymptomatic model

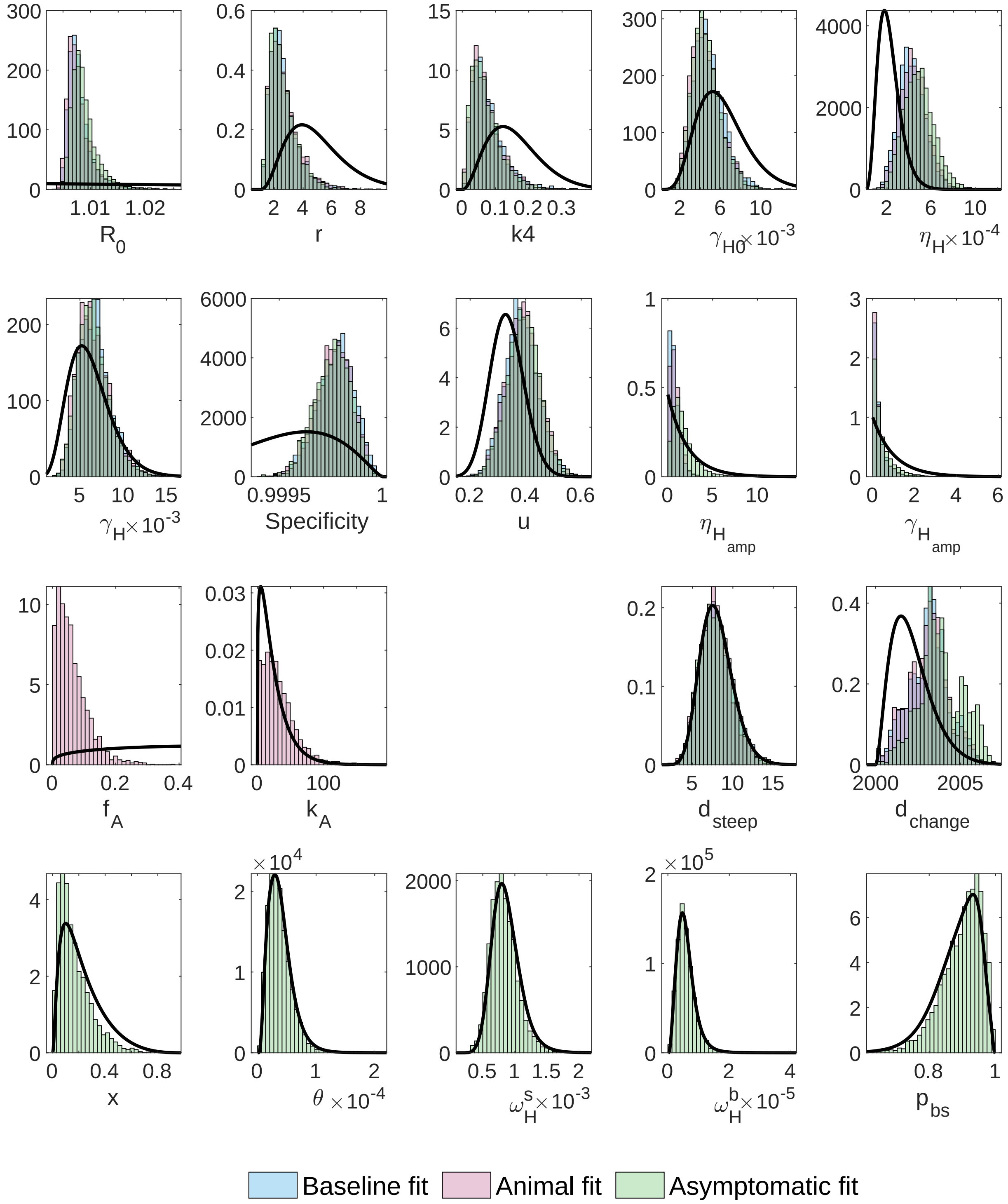


Fig 10: **Posterior parameter distributions for Budjala (after sequential Bayesian (SBU))** using the deterministic model for fitting each of the three model variants to case data. There are two fitted parameters which only appear in the animal model, and five which only appear in the asymptomatic model

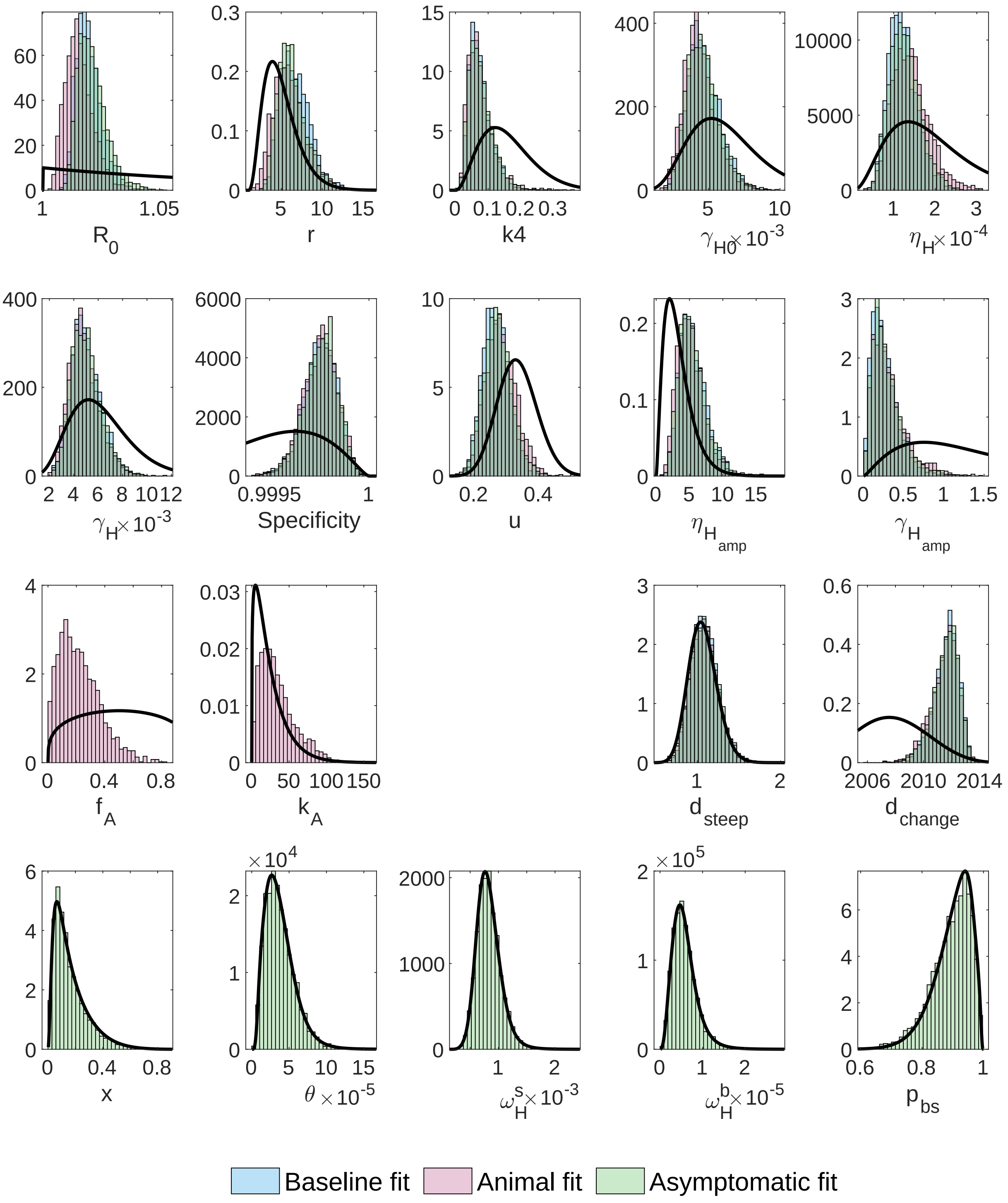


Fig 11: **Posterior parameter distributions for Bagata (after sequential Bayesian (SBU))** using the deterministic model for fitting each of the three model variants to case data. There are two fitted parameters which only appear in the animal model, and five which only appear in the asymptomatic model

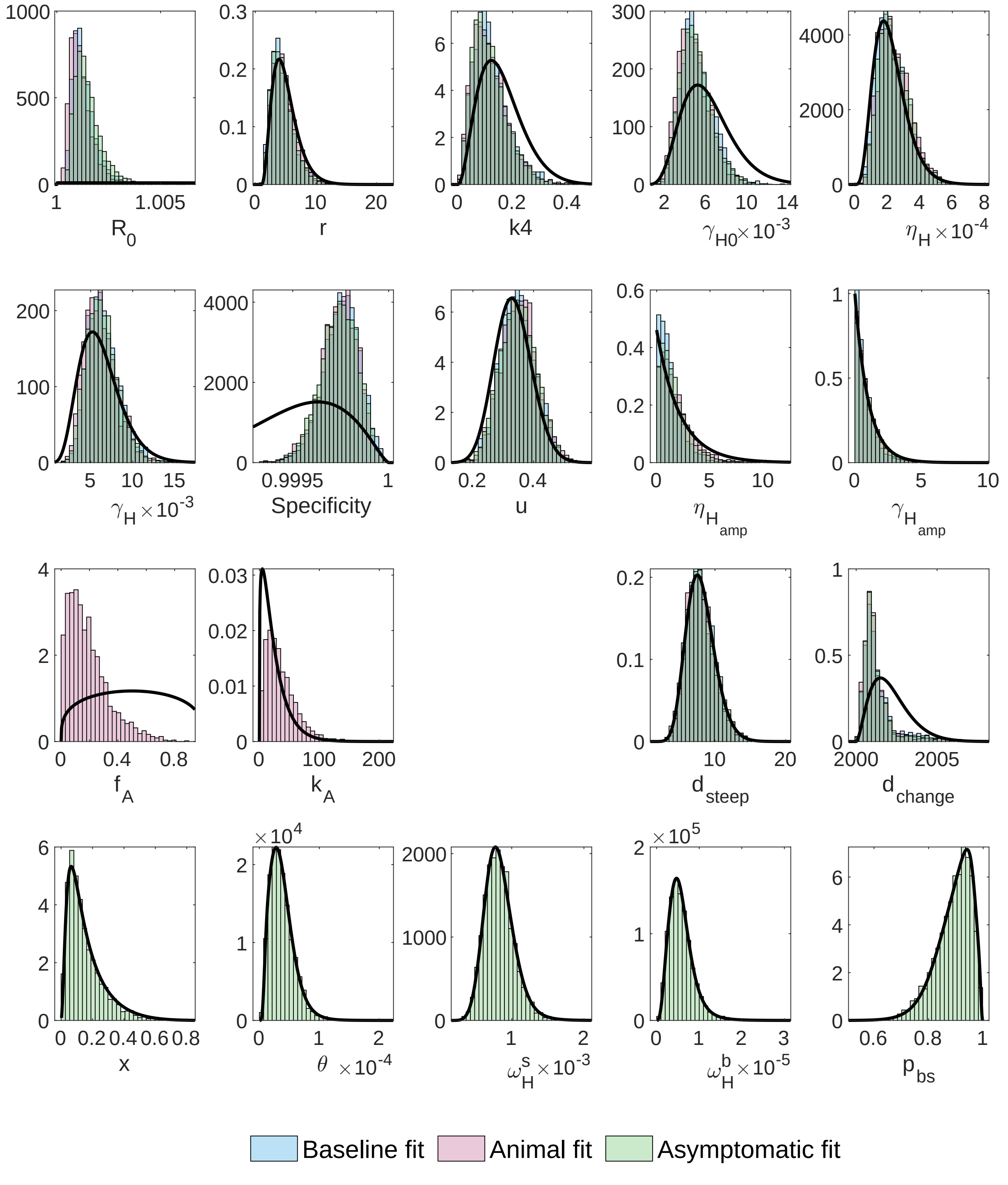


Fig 12: **Posterior parameter distributions for Mbaya (after sequential Bayesian (SBU))** using the deterministic model for fitting each of the three model variants to case data. There are two fitted parameters which only appear in the animal model, and five which only appear in the asymptomatic model

#### Model fitting – time series plots

The following graphs show a comparison of our model outputs to data and include the estimated new human infection incidence.

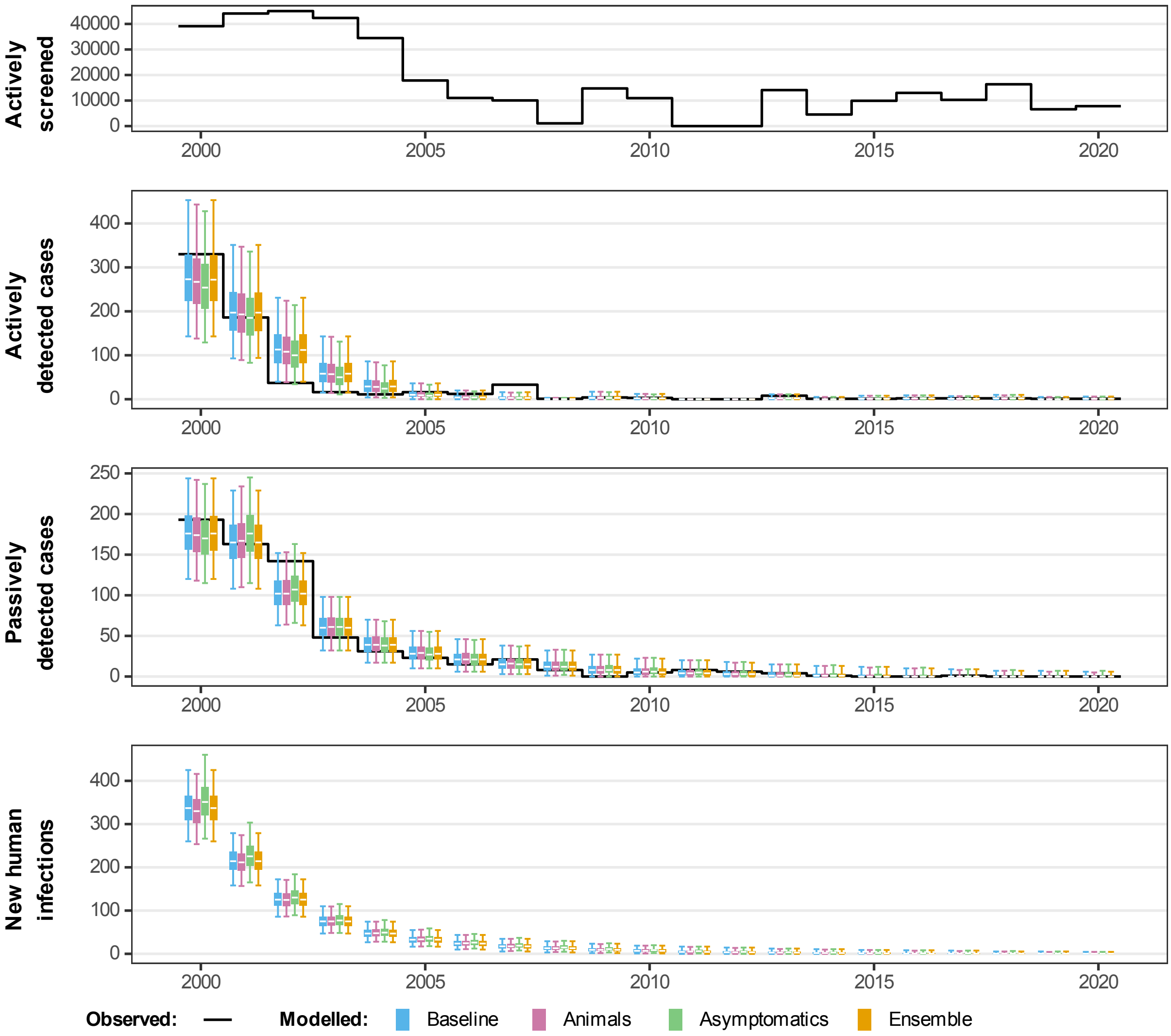


Fig 13: **Comparison of fits in Bominenge.** The deterministic model was used to perform fitting and sampling was conducted by using the stochastic model with the fitted posterior distributions. Blue, pink and green box and whisker plots show the baseline model, model with animal transmission and asymptomatic model fits respectively. The orange boxes represent the ensemble model outputs. The central line of each box is the median, the box is the 50% credible interval (CI) and the whiskers show the 95% CI. Case data are shown as a black line. New infections are estimated through the model fit, however, there is no way to directly observe this so there are no corresponding observational data.

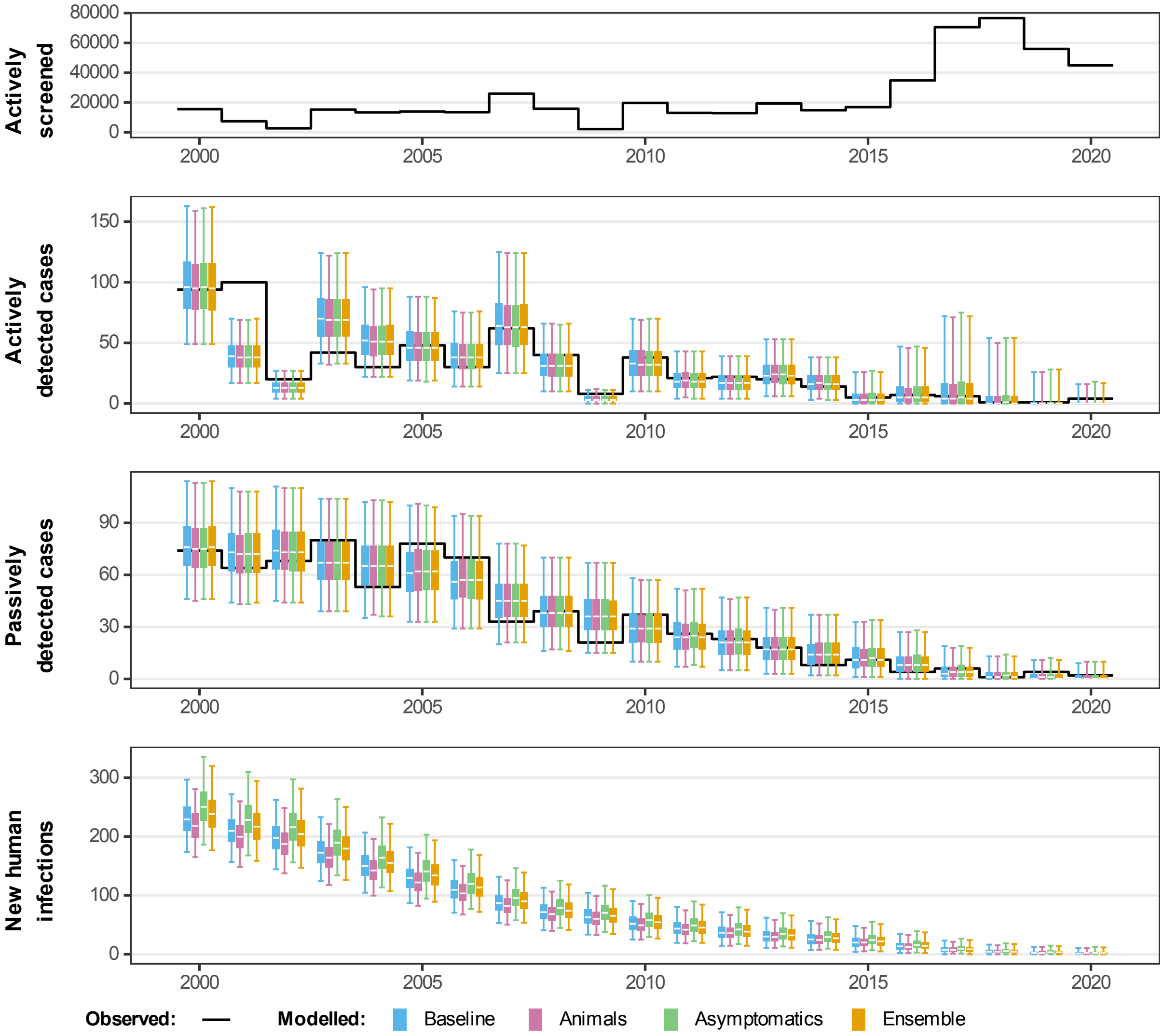


Fig 14: **Comparison of fits in Mosango.** The deterministic model was used to perform fitting and sampling was conducted by using the stochastic model with the fitted posterior distributions. Blue, pink and green box and whisker plots show the baseline model, model with animal transmission and asymptomatic model fits respectively. The orange boxes represent the ensemble model outputs. The central line of each box is the median, the box is the 50% credible interval (CI) and the whiskers show the 95% CI. Case data are shown as a black line. New infections are estimated through the model fit, however, there is no way to directly observe this so there are no corresponding observational data.

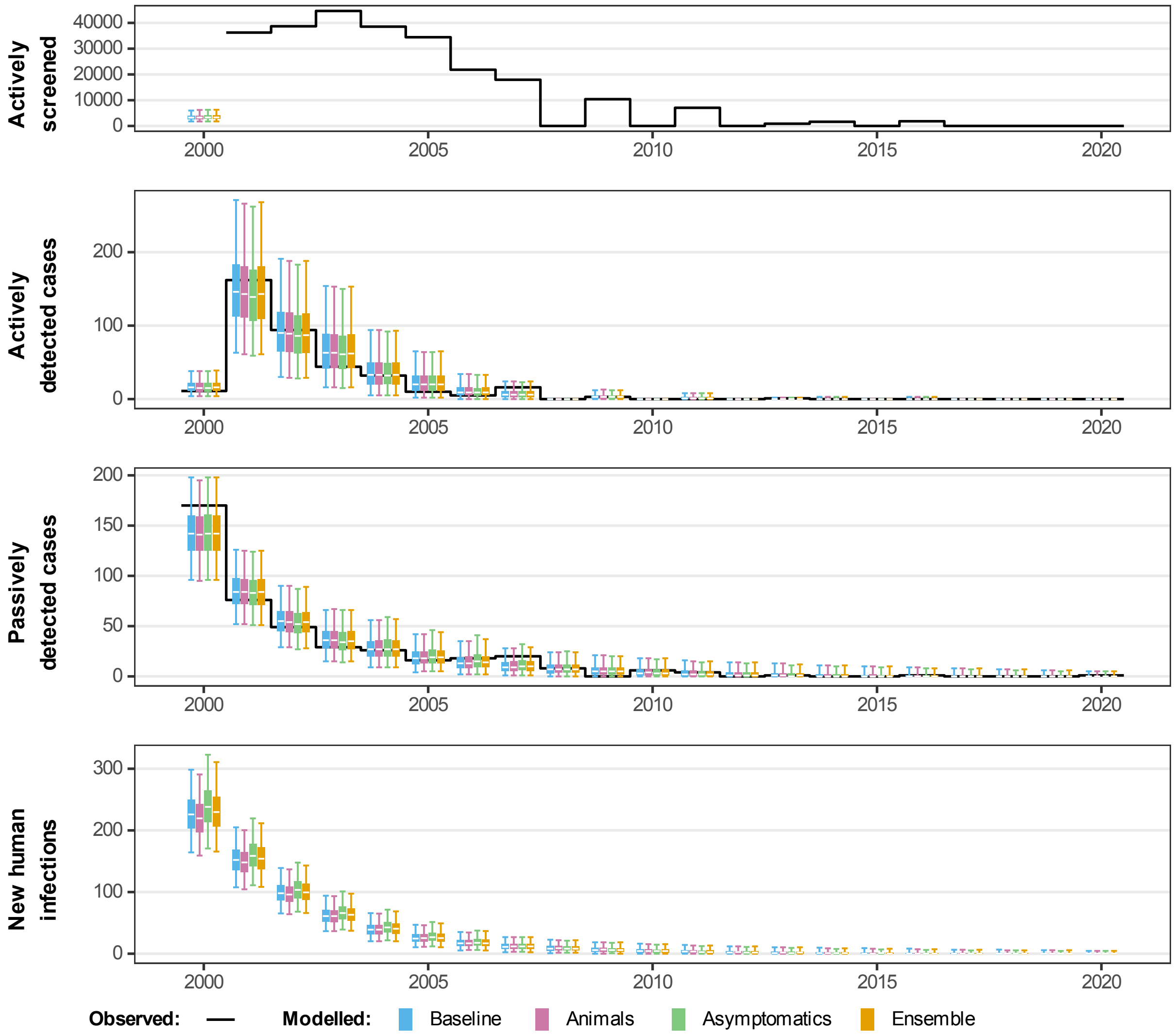


Fig 15: **Comparison of fits in Budjala.** The deterministic model was used to perform fitting and sampling was conducted by using the stochastic model with the fitted posterior distributions. Blue, pink and green box and whisker plots show the baseline model, model with animal transmission and asymptomatic model fits respectively. The orange boxes represent the ensemble model outputs. The central line of each box is the median, the box is the 50% credible interval (CI) and the whiskers show the 95% CI. Case data are shown as a black line. New infections are estimated through the model fit, however, there is no way to directly observe this so there are no corresponding observational data.

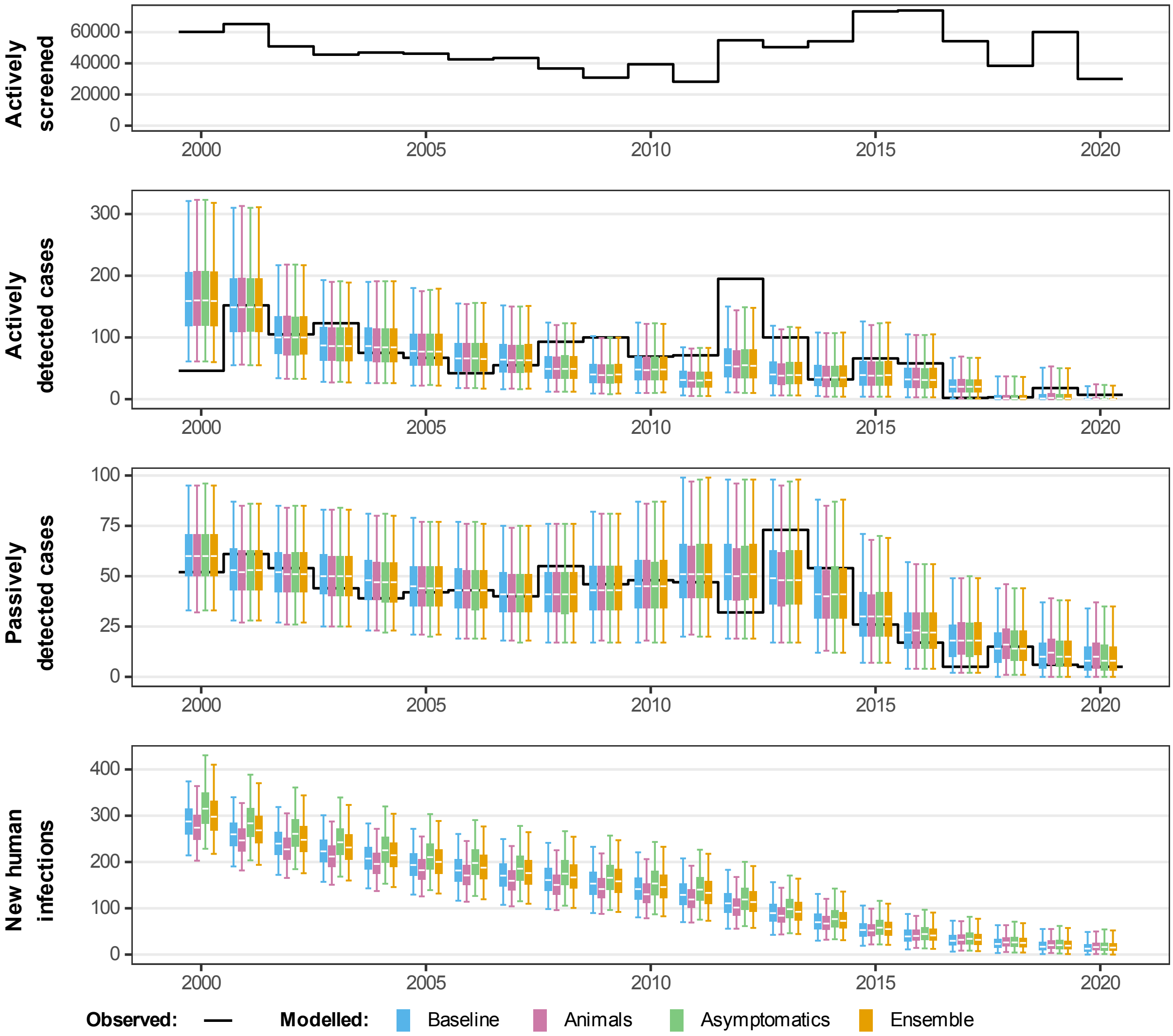


Fig 16: **Comparison of fits to data in Bagata.** The deterministic model was used to perform fitting and sampling was conducted by using the stochastic model with the fitted posterior distributions. Blue, pink and green box and whisker plots show the baseline model, model with animal transmission and asymptomatic model fits respectively. The orange boxes represent the ensemble model outputs. The central line of each box is the median, the box is the 50% credible interval (CI) and the whiskers show the 95% CI. Case data are shown as a black line. New infections are estimated through the model fit, however, there is no way to directly observe this so there are no corresponding observational data.

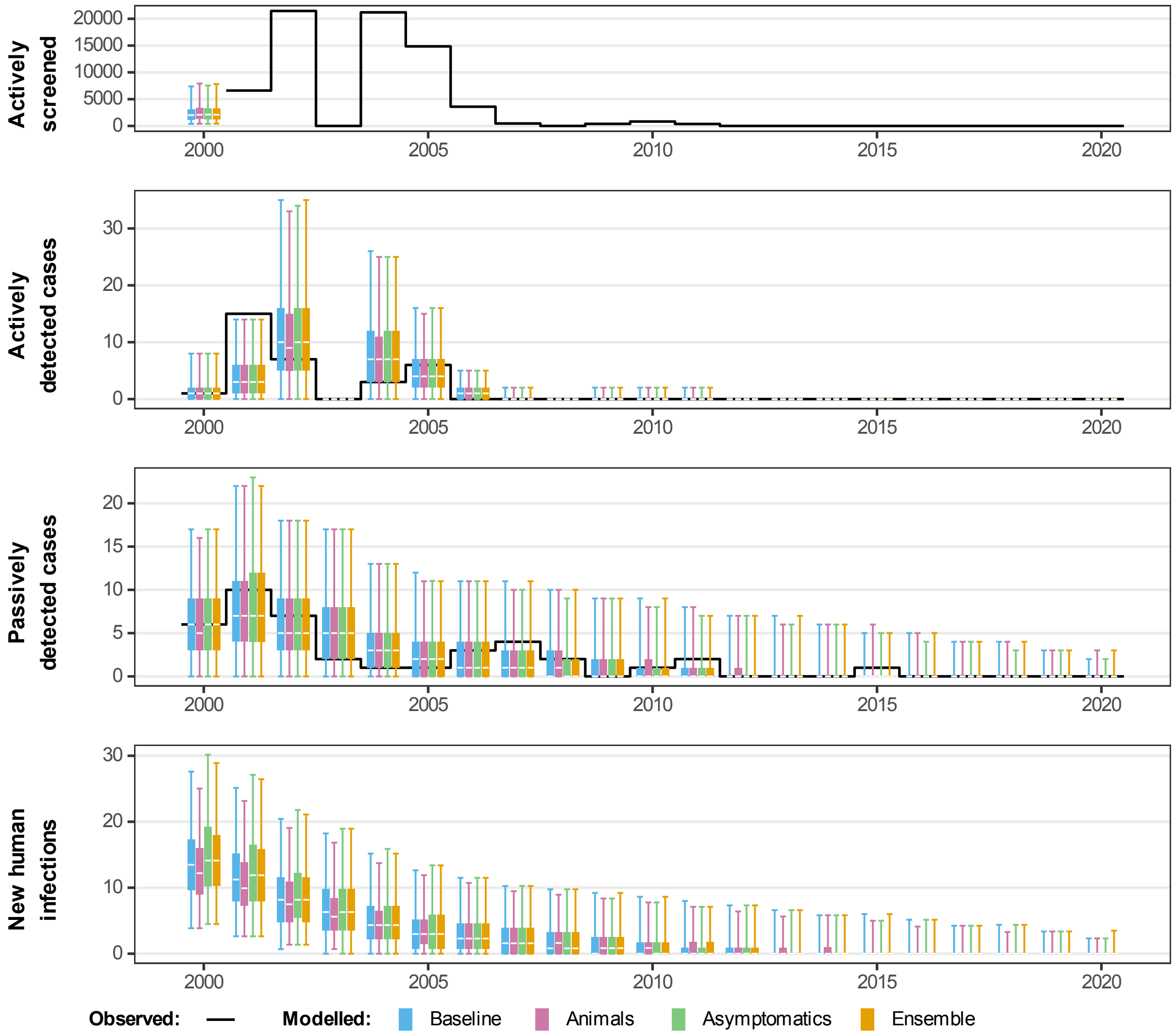


Fig 17: **Comparison of fits in Mbaya.** The deterministic model was used to perform fitting and sampling was conducted by using the stochastic model with the fitted posterior distributions. Blue, pink and green box and whisker plots show the baseline model, model with animal transmission and asymptomatic model fits respectively. The orange boxes represent the ensemble model outputs. The central line of each box is the median, the box is the 50% credible interval (CI) and the whiskers show the 95% CI. Case data are shown as a black line. New infections are estimated through the model fit, however, there is no way to directly observe this so there are no corresponding observational data.

#### Model projections

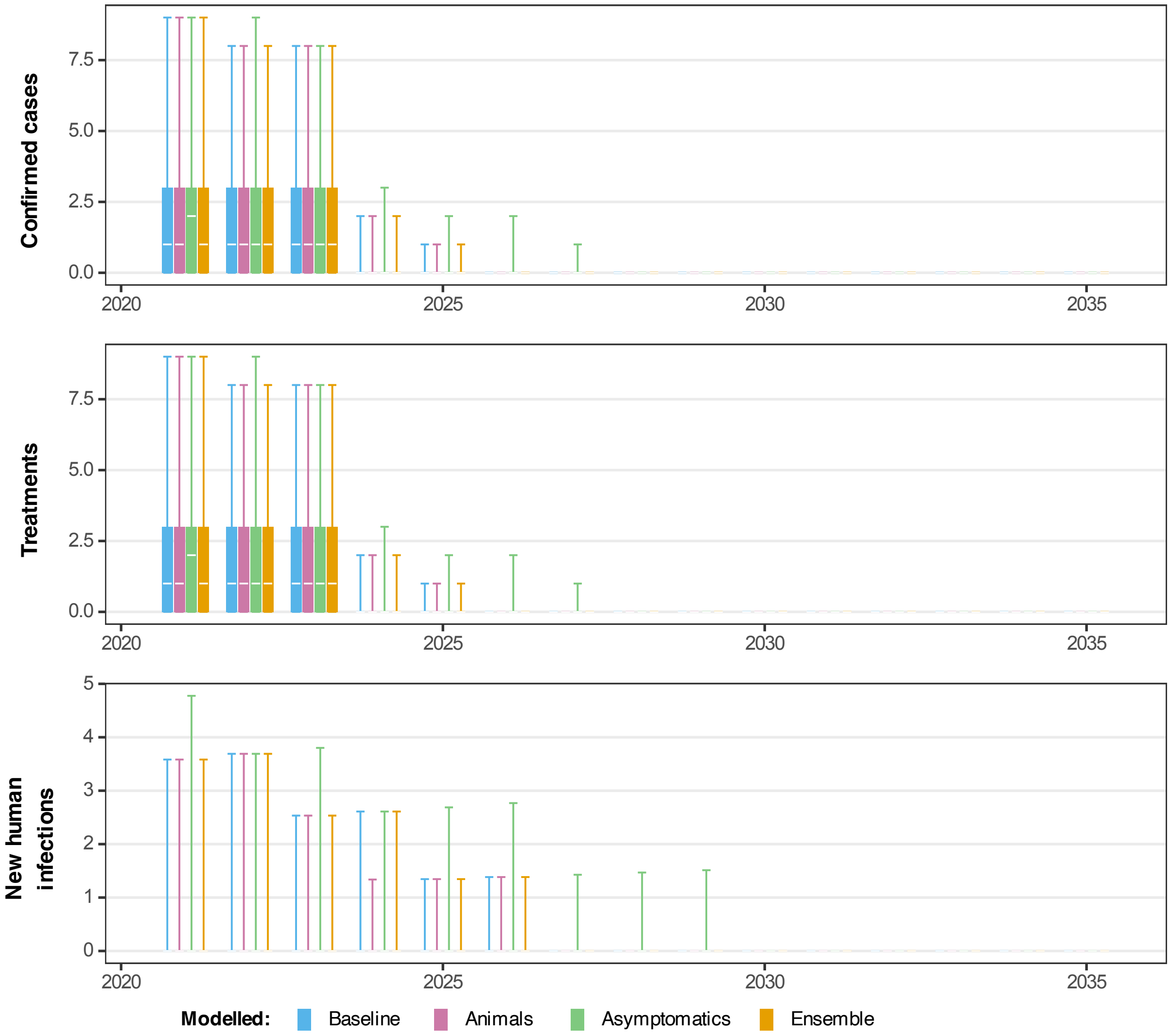


Fig 18: **Projected dynamics in Bominenge health zone in Equateur Nord coordination under Mean active screening strategy.** Comparing three model variants using the stochastic model including projections for 2021–2035 under a MeanAS strategy (using AS coverage for Bominenge from 2016–2020). Blue, pink, green and orange box and whisker plots show the baseline model, model with animal transmission, asymptomatic model and ensemble model projections respectively. The central line of each box is the median, the box is the 50% prediction interval (PI) and the whiskers show the 95% PI.

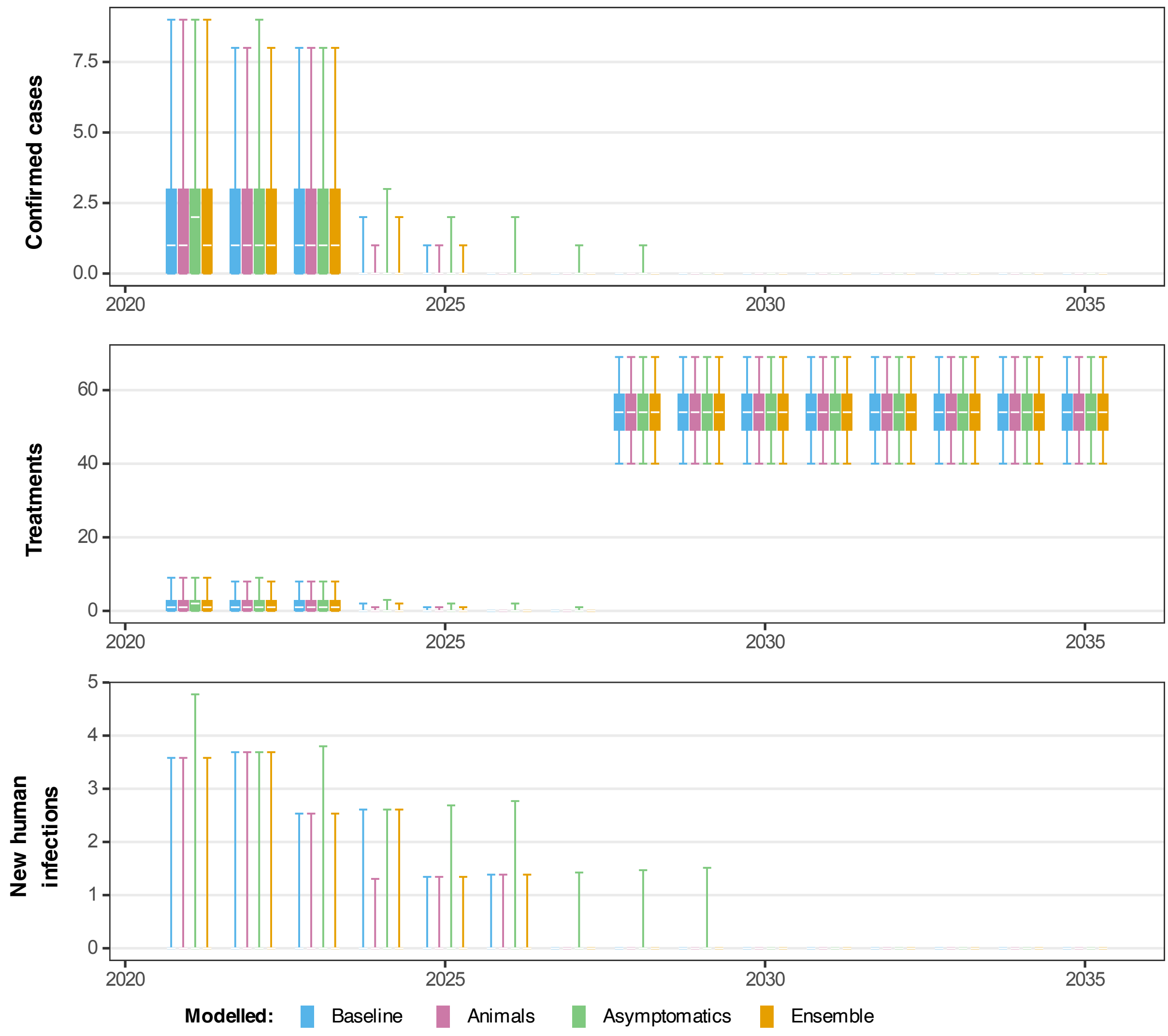


Fig 19: **Projected dynamics in Bominenge health zone in Equateur Nord coordination under Mean screen-and-treat strategy.** Comparing three model variants using the stochastic model including projections for 2021–2027 under a MeanAS strategy followed by MeanS&T at the same coverage level for 2028–2035 (using AS coverage for Bominenge from 2016–2020). Blue, pink, green and orange box and whisker plots show the baseline model, model with animal transmission, asymptomatic model and ensemble model projections respectively. The central line of each box is the median, the box is the 50% prediction interval (PI) and the whiskers show the 95% PI.

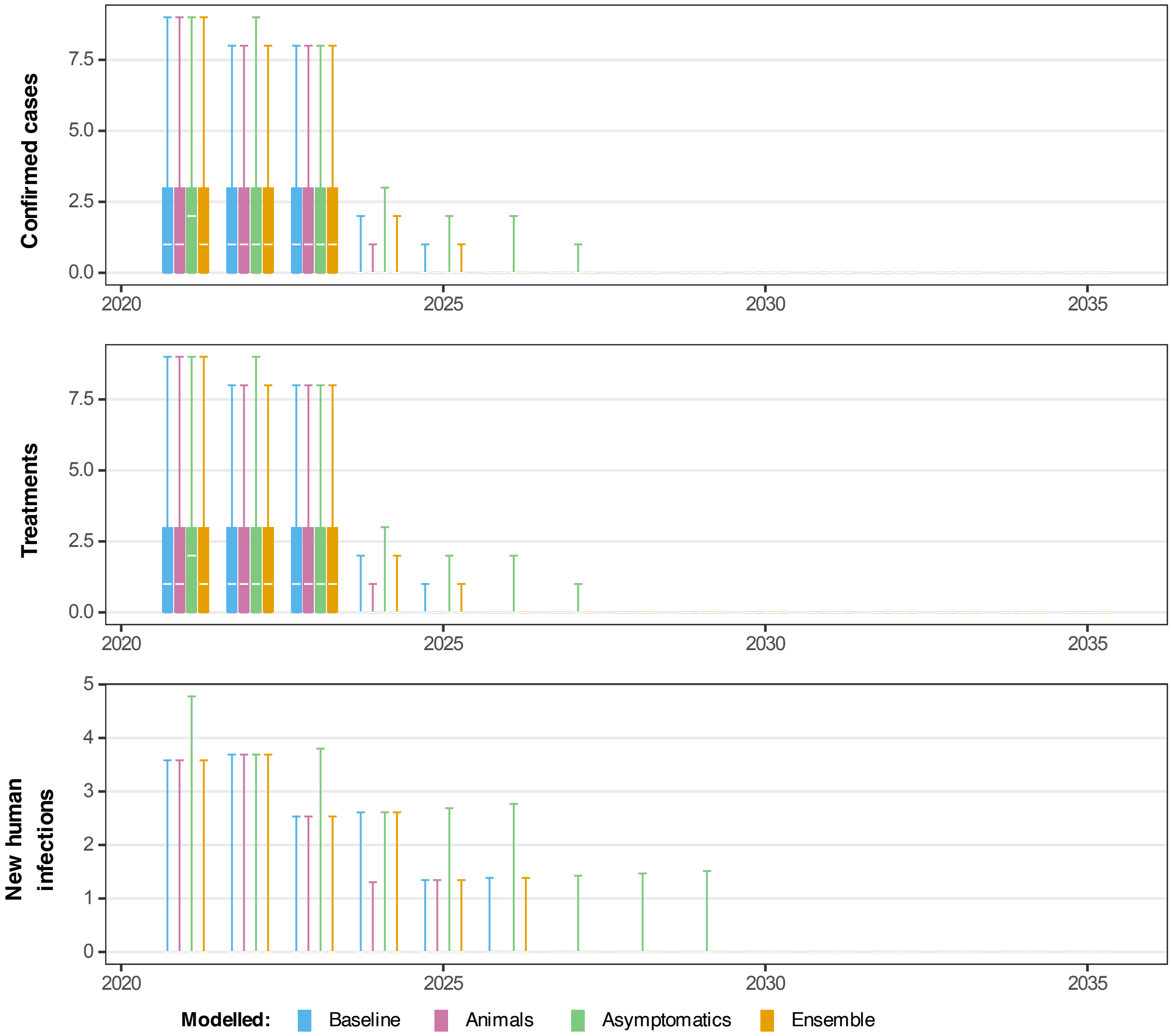


Fig 20: **Projected dynamics in Bominenge health zone in Equateur Nord coordination under Mean active screening strategy with vector control.** Comparing three model variants using the stochastic model including projections for 2021–2023 under a MeanAS strategy and 2024–2035 under a MeanAS+VC strategy with an assumed 80% reduction in tsetse population after 1 year (using AS coverage for Bominenge from 2016–2020). Blue, pink, green and orange box and whisker plots show the baseline model, model with animal transmission, asymptomatic model and ensemble model projections respectively. The central line of each box is the median, the box is the 50% prediction interval (PI) and the whiskers show the 95% PI.

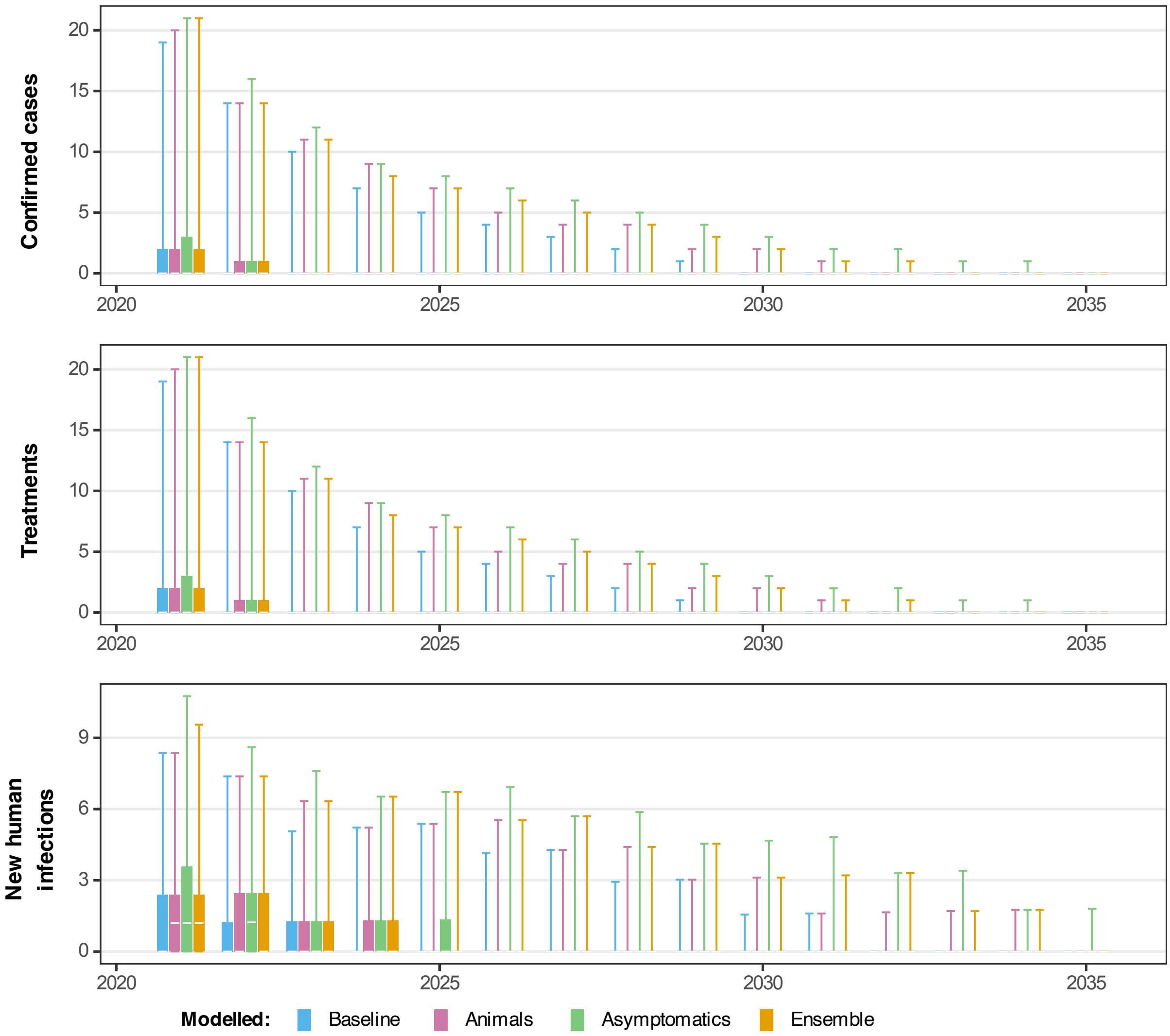


Fig 21: **Projected dynamics in Mosango health zone in Bandundu Sud coordination under Mean active screening strategy.** Comparing three model variants using the stochastic model including projections for 2021–2035 under a MeanAS strategy (using AS coverage for Mosango from 2016–2020). Blue, pink, green and orange box and whisker plots show the baseline model, model with animal transmission, asymptomatic model and ensemble model projections respectively. The central line of each box is the median, the box is the 50% prediction interval (PI) and the whiskers show the 95% PI.

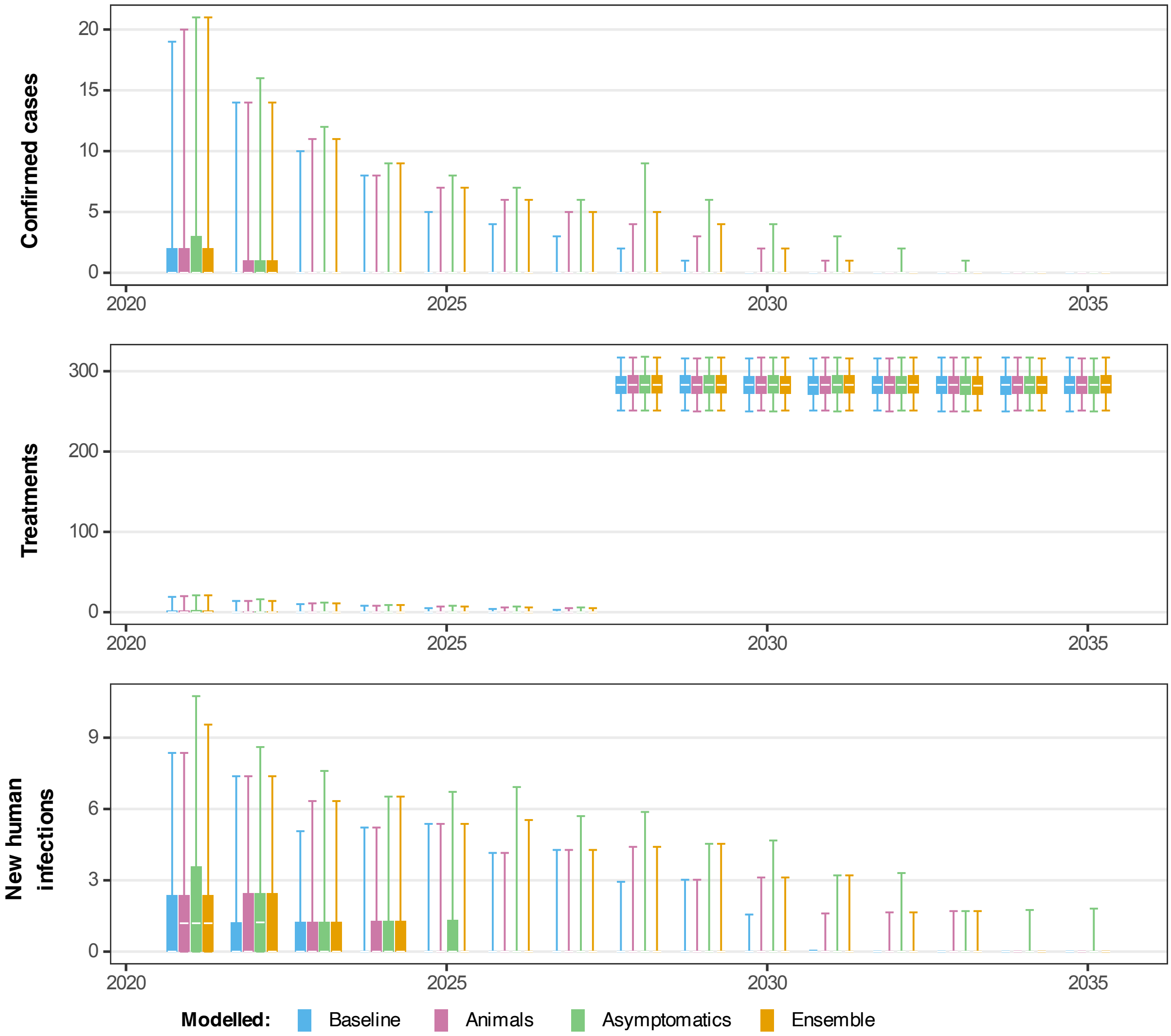


Fig 22: **Projected dynamics in Mosango health zone in Bandundu Sud coordination under Mean screen-and-treat strategy.** Comparing three model variants using the stochastic model including projections for 2021–2027 under a MeanAS strategy followed by MeanS&T at the same coverage level for 2028–2035 (using AS coverage for Mosango from 2016–2020). Blue, pink, green and orange box and whisker plots show the baseline model, model with animal transmission, asymptomatic model and ensemble model projections respectively. The central line of each box is the median, the box is the 50% prediction interval (PI) and the whiskers show the 95% PI.

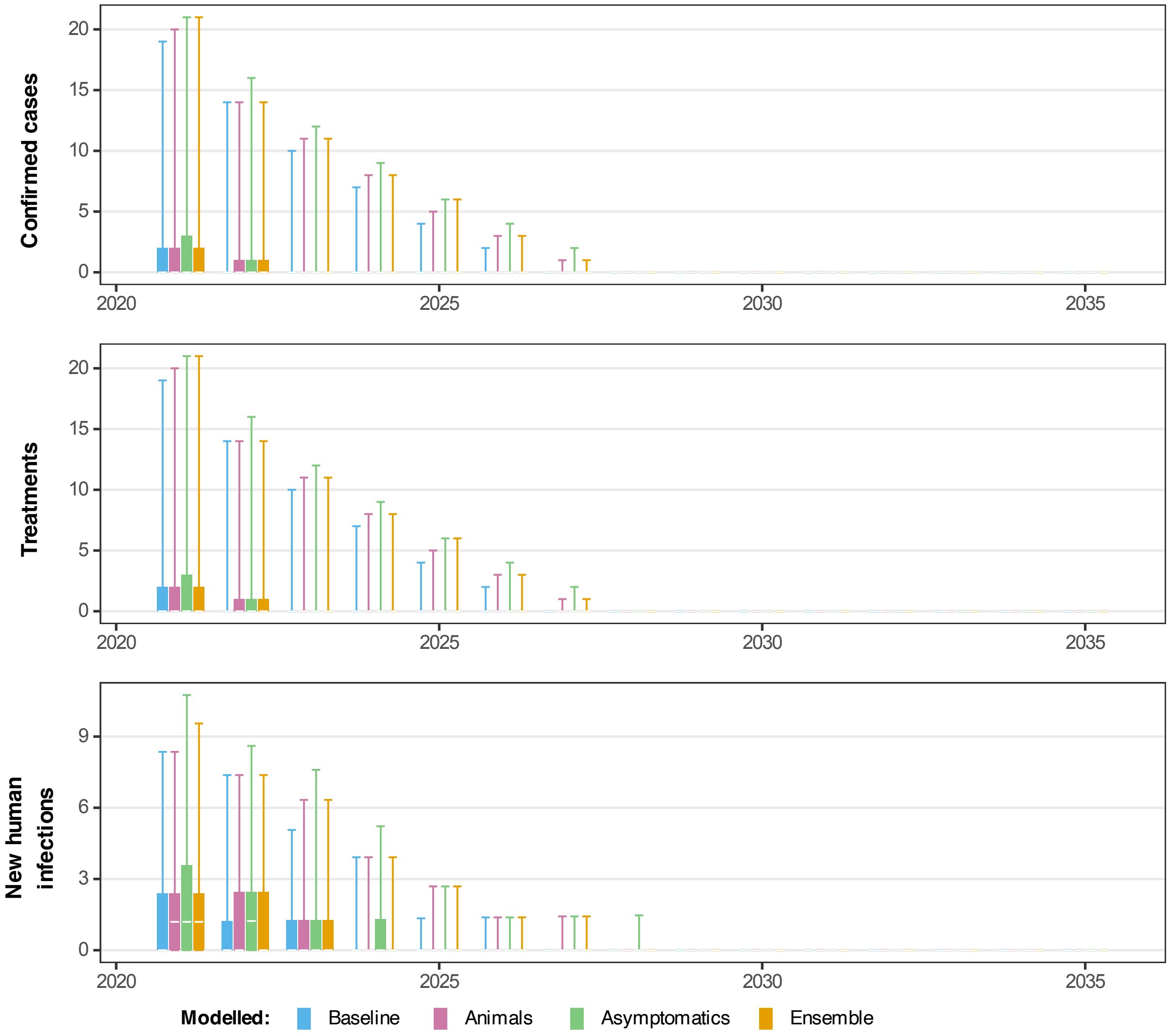


Fig 23: **Projected dynamics in Mosango health zone in Bandundu Sud coordination under Mean active screening strategy with vector control.** Comparing three model variants using the stochastic model including projections for 2021–2023 under a MeanAS strategy and 2024–2035 under a MeanAS+VC strategy with an assumed 80% reduction in tsetse population after 1 year (using AS coverage for Mosango from 2016–2020). Blue, pink, green and orange box and whisker plots show the baseline model, model with animal transmission, asymptomatic model and ensemble model projections respectively. The central line of each box is the median, the box is the 50% prediction interval (PI) and the whiskers show the 95% PI.

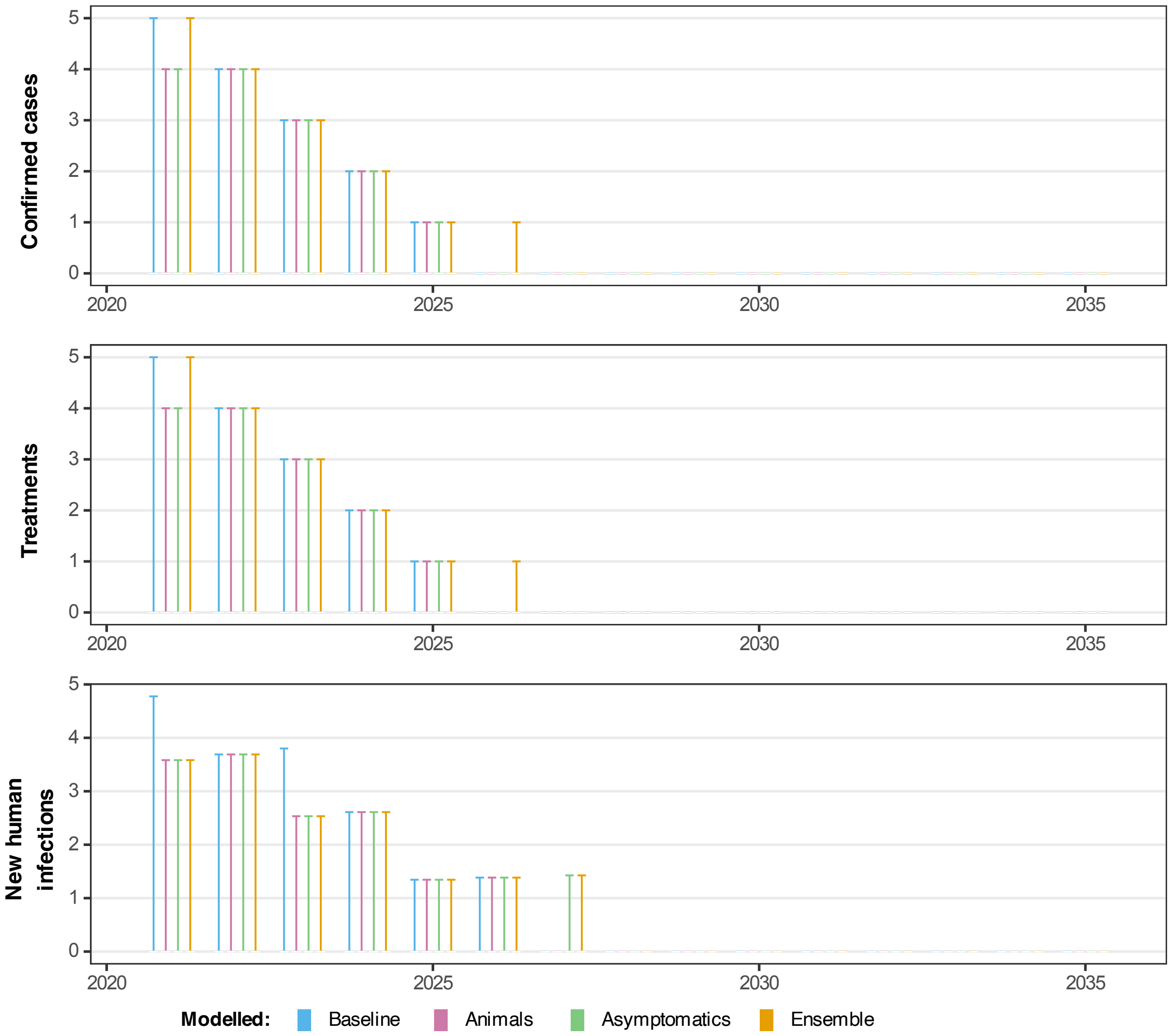


Fig 24: **Projected dynamics in Budjala health zone in Equateur Nord coordination under Mean active screening strategy.** Comparing three model variants using the stochastic model including projections for 2021–2035 under a MeanAS strategy (using AS coverage for Budjala from 2016–2020). Blue, pink, green and orange box and whisker plots show the baseline model, model with animal transmission, asymptomatic model and ensemble model projections respectively. The central line of each box is the median, the box is the 50% prediction interval (PI) and the whiskers show the 95% PI.

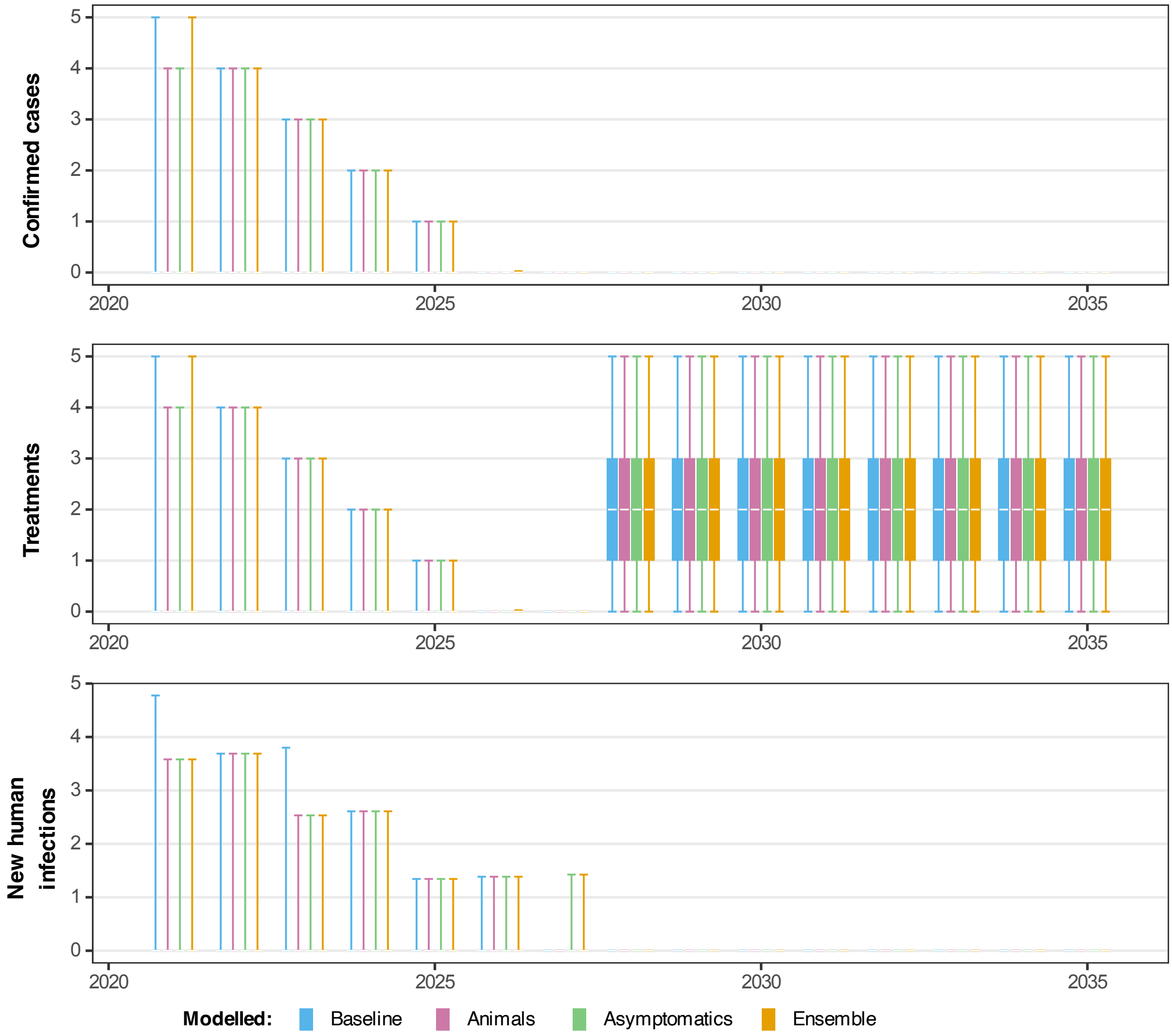


Fig 25: **Projected dynamics in Budjala health zone in Equateur Nord coordination under Mean screen-and-treat strategy.** Comparing three model variants using the stochastic model including projections for 2021–2027 under a MeanAS strategy followed by MeanS&T at the same coverage level for 2028–2035 (using AS coverage for Budjala from 2016–2020). Blue, pink, green and orange box and whisker plots show the baseline model, model with animal transmission, asymptomatic model and ensemble model projections respectively. The central line of each box is the median, the box is the 50% prediction interval (PI) and the whiskers show the 95% PI.

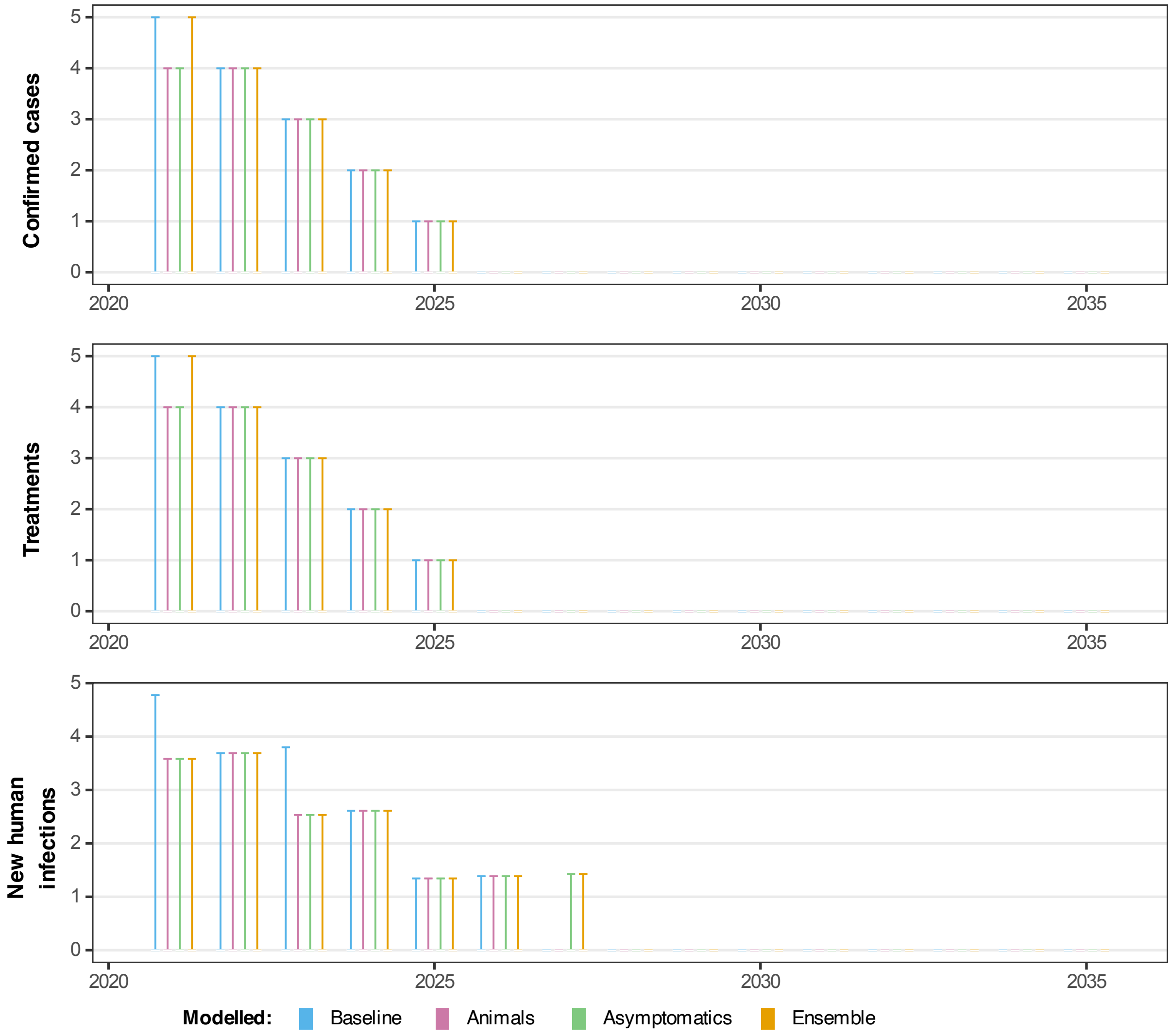


Fig 26: **Projected dynamics in Budjala health zone in Equateur Nord coordination under Mean active screening strategy with vector control.** Comparing three model variants using the stochastic model including projections for 2021–2023 under a MeanAS strategy and 2024–2035 under a MeanAS+VC strategy with an assumed 80% reduction in tsetse population after 1 year (using AS coverage for Budjala from 2016–2020). Blue, pink, green and orange box and whisker plots show the baseline model, model with animal transmission, asymptomatic model and ensemble model projections respectively. The central line of each box is the median, the box is the 50% prediction interval (PI) and the whiskers show the 95% PI.

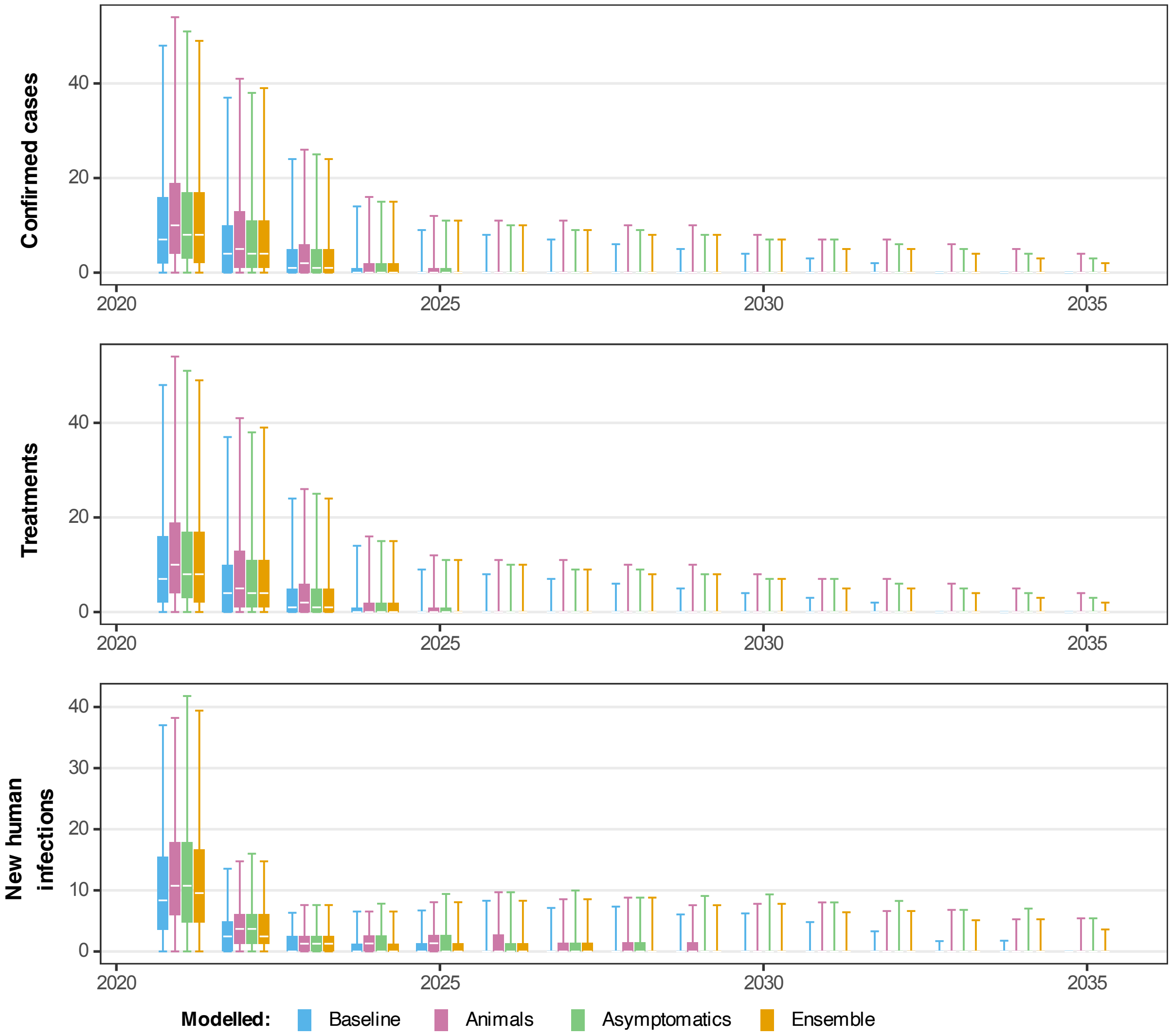


Fig 27: **Projected dynamics in Bagata health zone in Bandundu Nord coordination under Mean active screening strategy.** Comparing three model variants using the stochastic model including projections for 2021–2035 under a MeanAS strategy (using AS coverage for Bagata from 2016–2020). Blue, pink, green and orange box and whisker plots show the baseline model, model with animal transmission, asymptomatic model and ensemble model projections respectively. The central line of each box is the median, the box is the 50% prediction interval (PI) and the whiskers show the 95% PI.

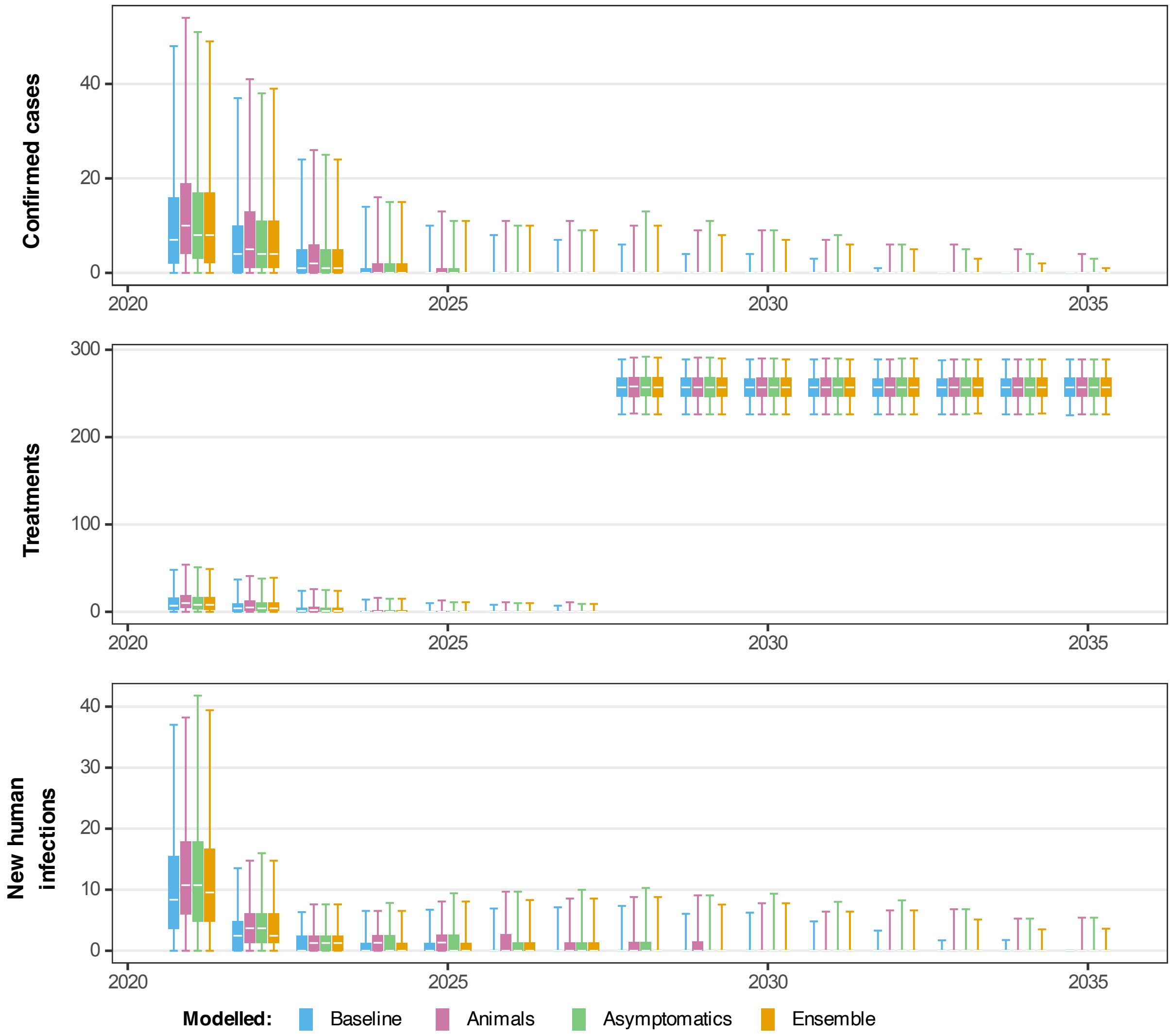


Fig 28: **Projected dynamics in Bagata health zone in Bandundu Nord coordination under Mean screen-and-treat strategy.** Comparing three model variants using the stochastic model including projections for 2021–2027 under a MeanAS strategy followed by MeanS&T at the same coverage level for 2028–2035 (using AS coverage for Bagata from 2016–2020). Blue, pink, green and orange box and whisker plots show the baseline model, model with animal transmission, asymptomatic model and ensemble model projections respectively. The central line of each box is the median, the box is the 50% prediction interval (PI) and the whiskers show the 95% PI.

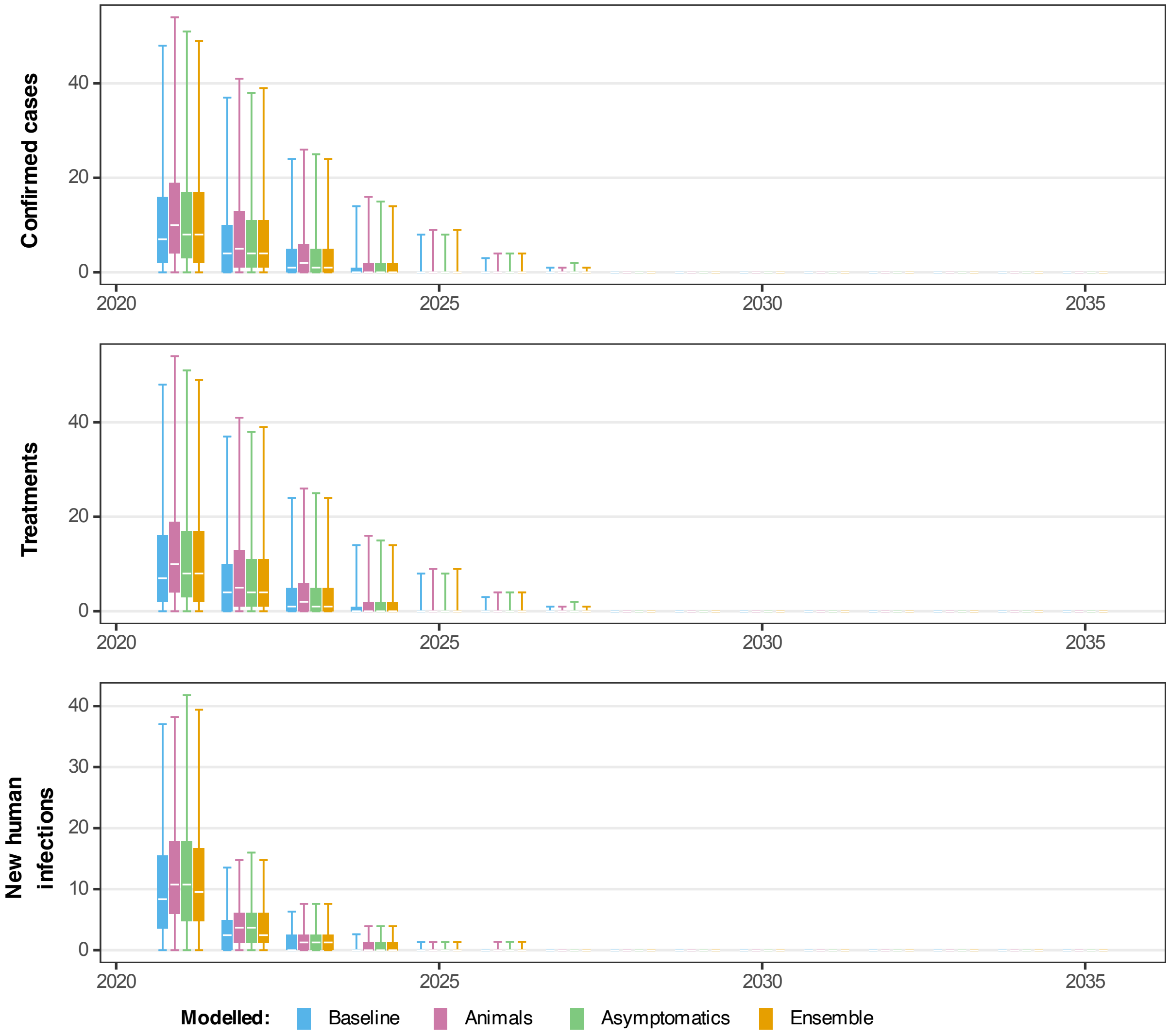


Fig 29: **Projected dynamics in Bagata health zone in Bandundu Nord coordination under Mean active screening strategy with vector control.** Comparing three model variants using the stochastic model including projections for 2021–2023 under a MeanAS strategy and 2024–2035 under a MeanAS+VC strategy with an assumed 80% reduction in tsetse population after 1 year (using AS coverage for Bagata from 2016–2020). Blue, pink, green and orange box and whisker plots show the baseline model, model with animal transmission, asymptomatic model and ensemble model projections respectively. The central line of each box is the median, the box is the 50% prediction interval (PI) and the whiskers show the 95% PI.

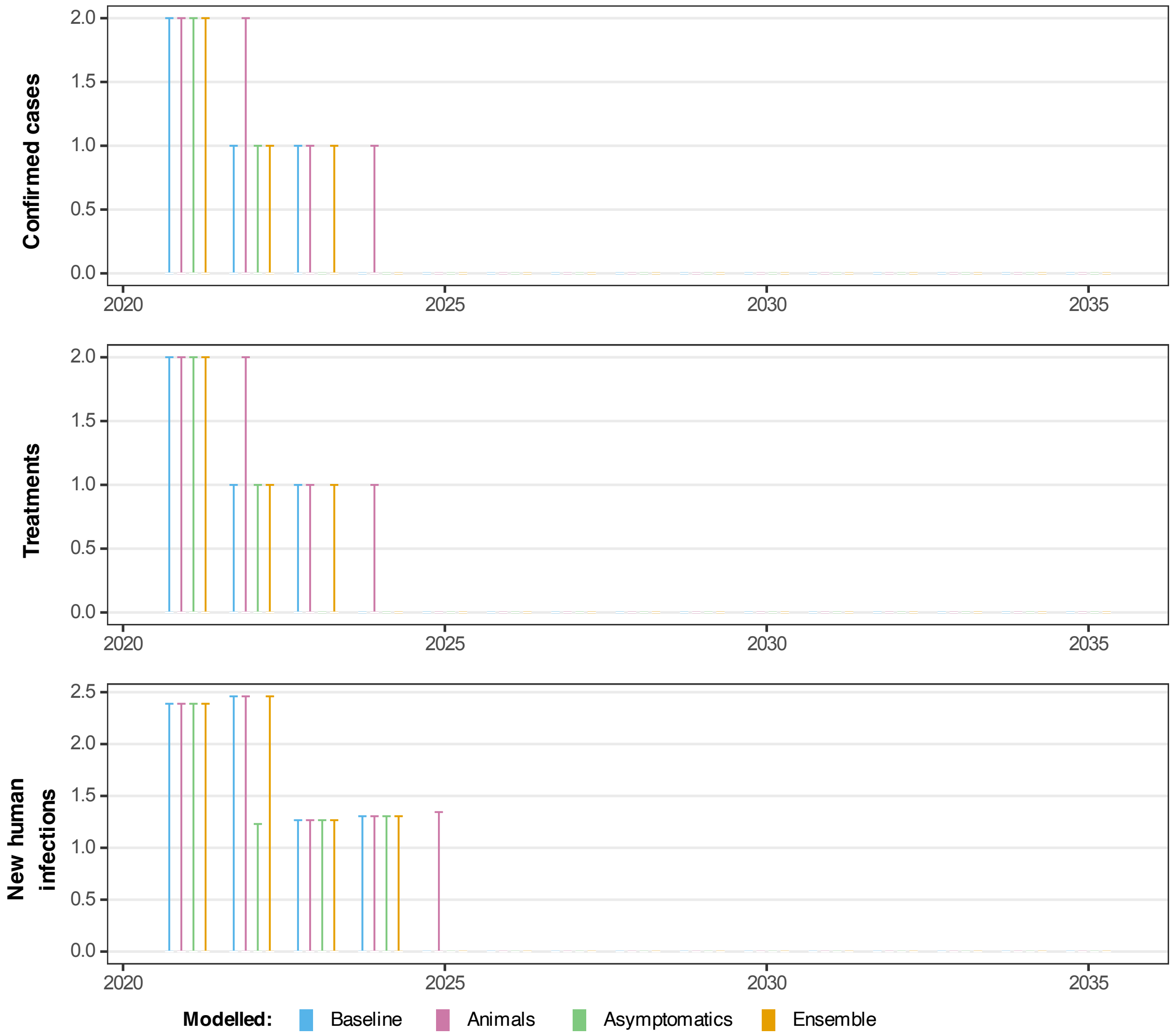


Fig 30: **Projected dynamics in Mbaya health zone in Equateur Nord coordination under Mean active screening strategy.** Comparing three model variants using the stochastic model including projections for 2021–2035 under a MeanAS strategy (using AS coverage for Mbaya from 2016–2020). Blue, pink, green and orange box and whisker plots show the baseline model, model with animal transmission, asymptomatic model and ensemble model projections respectively. The central line of each box is the median, the box is the 50% prediction interval (PI) and the whiskers show the 95% PI.

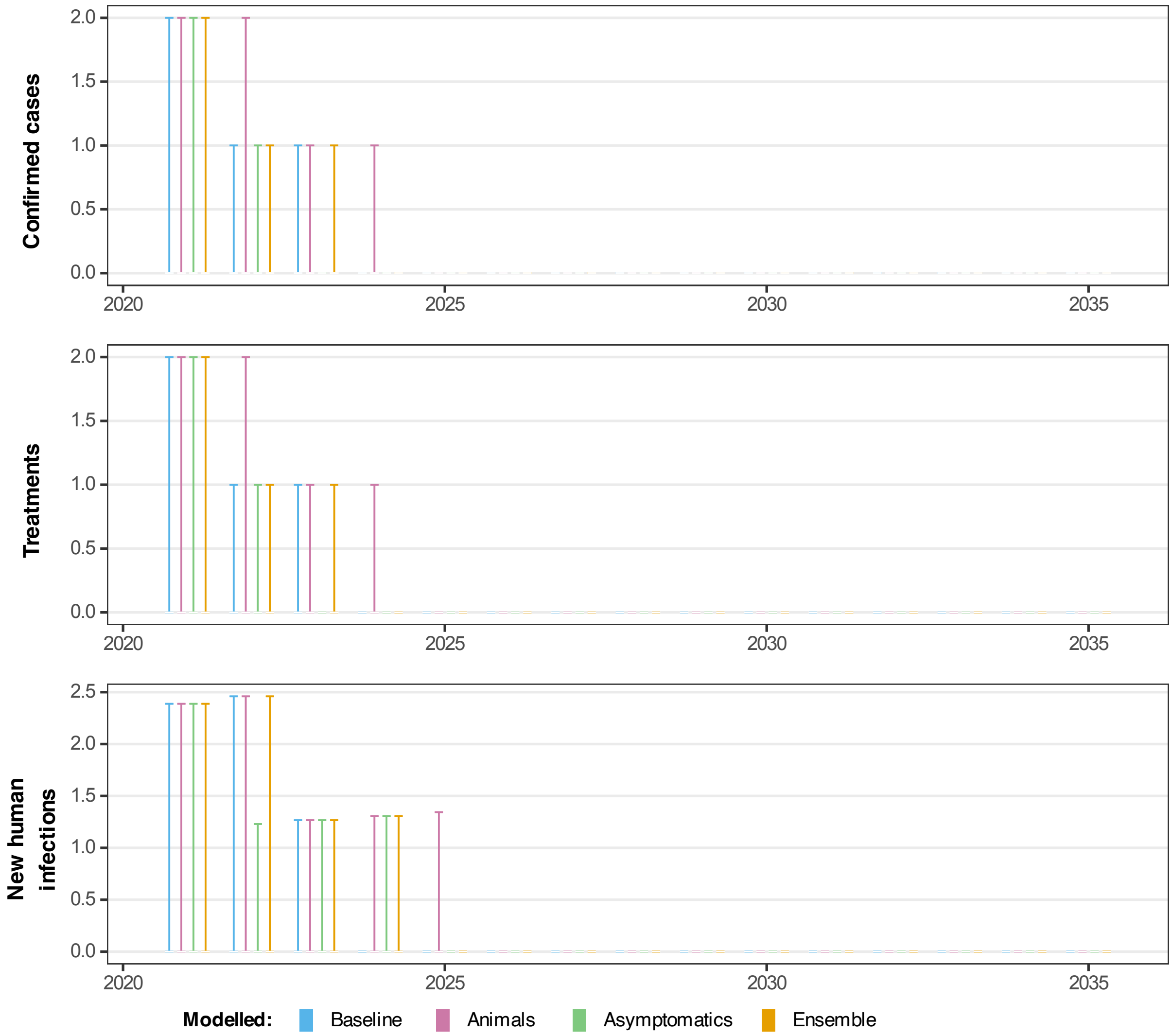


Fig 31: **Projected dynamics in Mbaya health zone in Equateur Nord coordination under Mean screen-and-treat strategy.** Comparing three model variants using the stochastic model including projections for 2021–2027 under a MeanAS strategy followed by MeanS&T at the same coverage level for 2028–2035 (using AS coverage for Mbaya from 2016–2020). Blue, pink, green and orange box and whisker plots show the baseline model, model with animal transmission, asymptomatic model and ensemble model projections respectively. The central line of each box is the median, the box is the 50% prediction interval (PI) and the whiskers show the 95% PI.

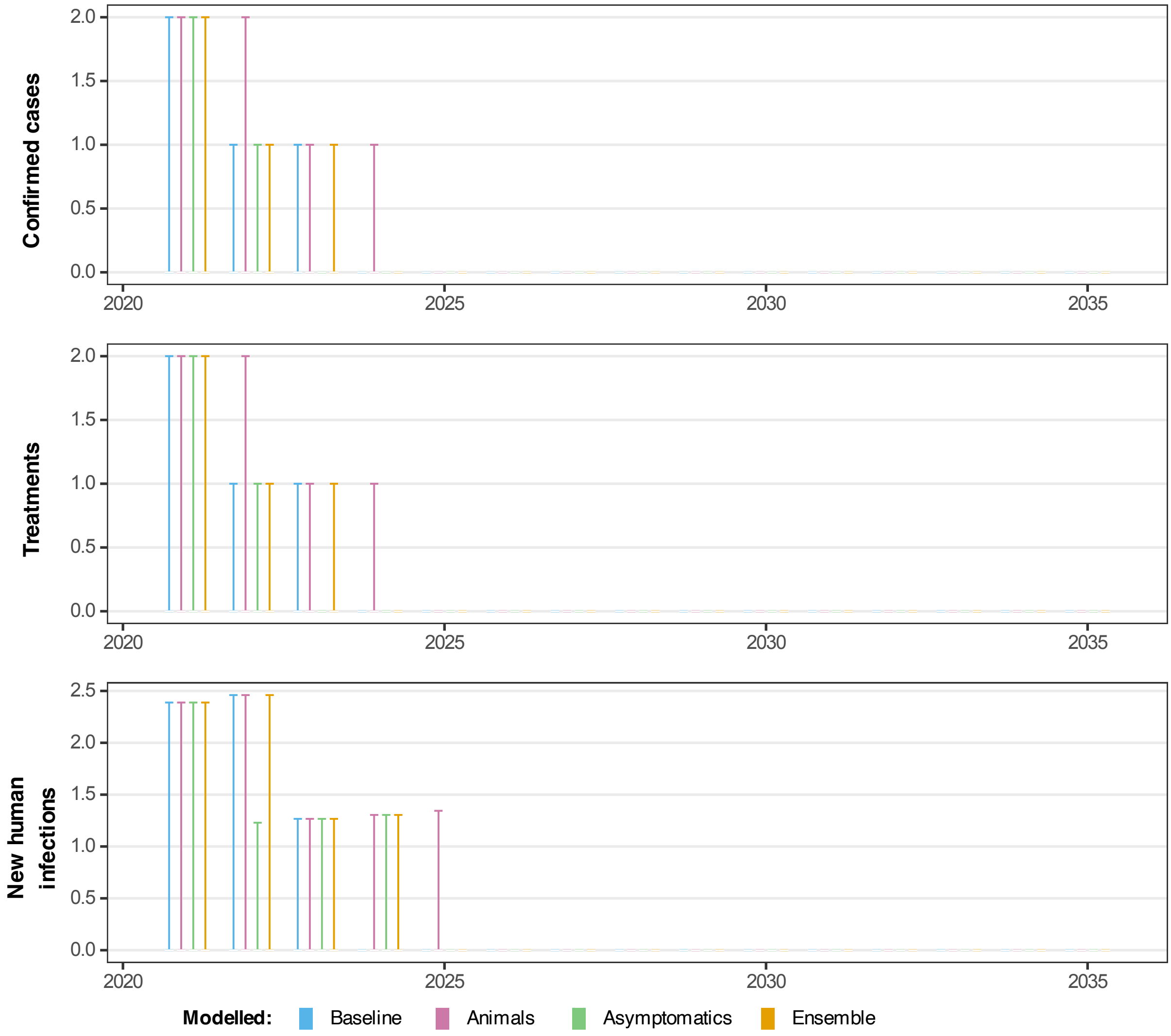


Fig 32: **Projected dynamics in Mbaya health zone in Equateur Nord coordination under Mean active screening strategy with vector control.** Comparing three model variants using the stochastic model including projections for 2021–2023 under a MeanAS strategy and 2024–2035 under a MeanAS+VC strategy with an assumed 80% reduction in tsetse population after 1 year (using AS coverage for Mbaya from 2016–2020). Blue, pink, green and orange box and whisker plots show the baseline model, model with animal transmission, asymptomatic model and ensemble model projections respectively. The central line of each box is the median, the box is the 50% prediction interval (PI) and the whiskers show the 95% PI.

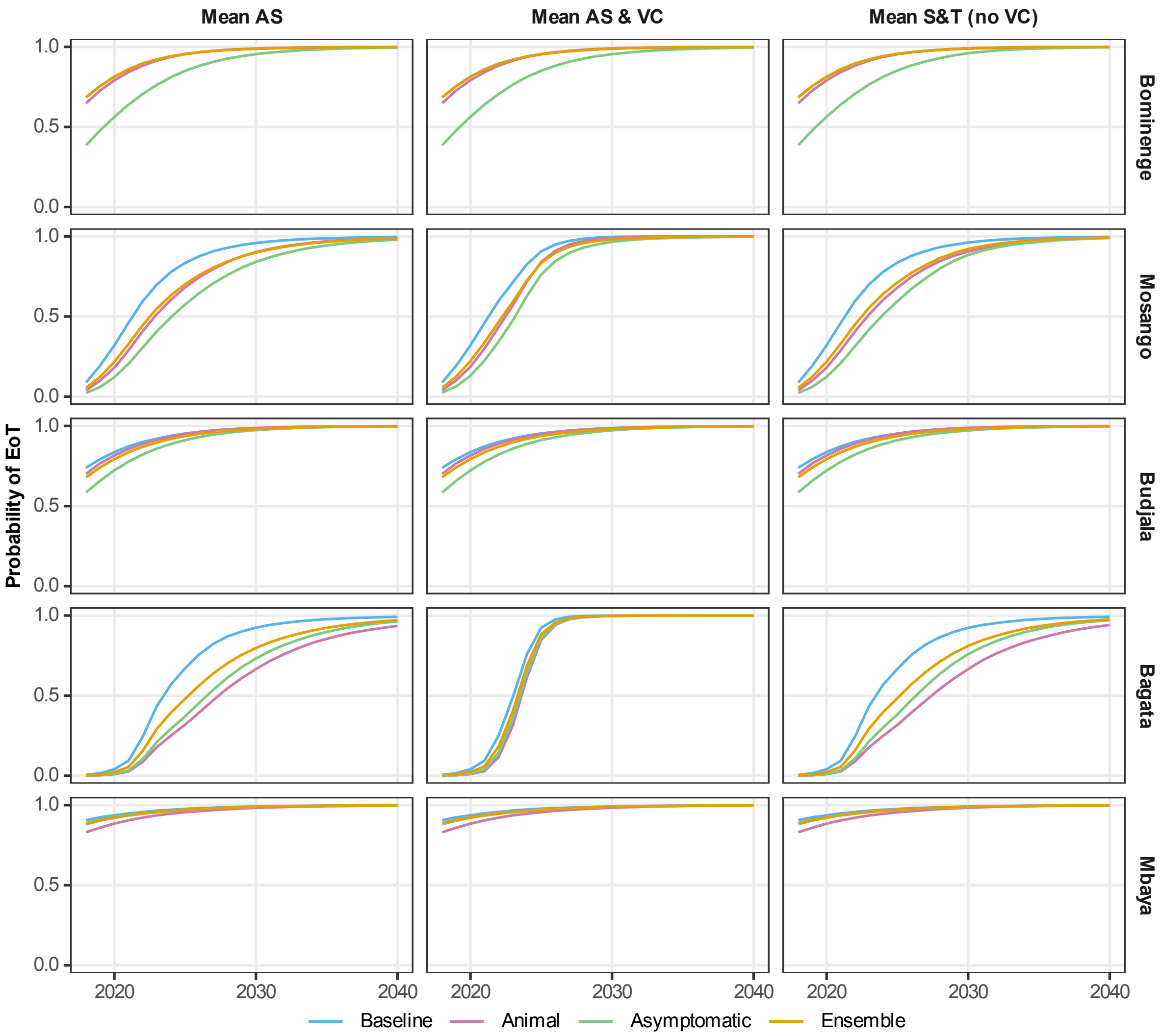


Fig 33: **Comparing the probability of EoT in each of the modelled health zones**, under each model variant and three different strategies. The blue, pink and green curves represent the model-estimated probability of EoT by each year, calculated by taking the number of realisations where there are no new infections to humans in or after that year until the end of the simulation and diving by the total number of realisations (20,000 for the baseline and animal models and 50,000 for the asymptomatic model) The ensemble results are given by the orange curve which is computed as the weighted average of the probability of EoT from the individual model variants to avoid statistical sampling variation. As per Table 2of the main text , for the second strategy with vector control (VC), we assume this novel intervention begins in 2024, and for the third strategy using screening-and-treat (S&T), we assume this novel intervention begins in 2028.

#### Summary of updates

The key updates presented in this study are highlighted below:

- This is the first time that the Warwick gHAT asymptomatic model variant (Model 9) has been fitted to data. Previously it was presented as part of a sensitivity analysis [1].
- The sequential Bayesian updating approach to fitting the asymptomatic model has not been used before in the fitting of a gHAT model to learn more about geographically invariant parameters.
- Previously model comparison of more than two Warwick gHAT models was done using weighting from the deviance information criteria (DIC), whereas here Bayes factors are used, extending the approach used by Crump et al. [10] to compare the baseline and animal models.
- No previous gHAT asymptomatic model fitting (from any group) has assessed whether the asymptomatic model is statistically likely compared to other model variants or quantified the impact of asymptomatic transmission on the predicted dynamics and elimination of transmission.
- This is the first time a gHAT model with the screen-and-treat strategy (where seropositive people receive treatment regardless of confirmation status) has been published by any group.

#### NTD PRIME criteria

To fulfil recommendations set out for good modelling practises for policy, we itemise below how each of the five key principles relating to communication, quality and relevance of analyses - known as Policy-Relevant Items for Reporting Models in Epidemiology of Neglected Tropical Diseases (PRIME-NTD) [3] have been met in Table E.

Table E: PRIME-NTD criteria fulfillment. How the NTD Modelling Consortium's "5 key principles of good modelling practice" have been met in the present study.

| Principle | What has been done to satisfy the principle? | Where in the manuscript is this described? |
|---|---|---|
| **1. Stakeholder engagement** | | |
| | This study is one in a series of modelling analyses for the DRC. The modelling team led the simulation and analysis work guided by members of the national sleeping sickness control programme in DRC (PNLTHA-DRC) – coauthors E Mwamba Miaka and S Chancy. PNLTHA-DRC have supported model development through the context of how the human case data used in the study were collected and how this has changed over time (via in-person meetings, online meetings and by email). | Authorship list |
| **2. Complete model documentation** | | |
| | Full model fitting code and documentation are available through OpenScienceFramework (OSF). The model is fully described in the main text and SI. | See Materials and Methods section in the main text, Supplementary Information (file S1) and at OSF (`https://osf.io/73ytc/`) |
| **3. Complete description of data used** | | |
| | The original data and how we aggregated the data for fitting were described in detail in the SI and also other model fitting papers for DRC from our group including a recent update. | See Supplementary Information, and Crump *et al.* [9] and [10] and Antillon et al. [2]. |
| **4. Communicating uncertainty** | | |
| | *Structural uncertainty*: The focus of this paper is to assess statistical support for three alternative gHAT model variants – one with no cryptic infections, one with animal transmission and one with self-curing asymptomatic humans. In previous studies, eight variants were considered [19, 14] and there was the highest support for "Model 4" (called baseline in the present study) and "Model 7" (animal transmission) which are examined here. The asymptomatic model has not been fitted or statistically compared previously. We show both fits and projections side by side to show model differences driven by this structural uncertainty<br>*Parameter uncertainty*: Parameter distributions are estimated by fitting to data. All model fits and projections include and propagate parameter uncertainty and include it in visual representations (either box and whisker plots or as probabilities, as appropriate) | *Structural uncertainty*: Materials and Methods section in the main text and all figures.<br>*Parameter uncertainty*: All main text figures and Supplementary Information (S1) figures |
| Continued on next page | | |

**Table E – continued from previous page**

| Principle | What has been done to satisfy the principle? | Where in the manuscript is this described? |
|---|---|---|
| | *Prediction uncertainty*: All predictions incorporate structural and parameter uncertainty. We show three strategies. | *Prediction uncertainty*: Main text and SI figures |
| **5. Testable model outcomes** | | |
| | Previous versions of this model have undergone validation exercises (data censoring) to examine the robustness of the predictive ability of the model [14, 18]. Whilst this was not performed here, the model is an updated version of those that have undergone validation, with updates based on the critical review of model fits as data for different regions or time periods (notable refinements also included here were made in the related analysis of Antillon et al. [2]). As more years of data become available in the future the model outputs shown here will be able to be compared to reported active and passive case data to test model predictions. | Main text and SI figures |

[9] R. E. Crump, C.-I. Huang, E. S. Knock, S. E. F. Spencer, P. E. Brown, E. Mwamba Miaka, C. Shampa, M. J. Keeling, and K. S. Rock. Quantifying epidemiological drivers of *gambiense* human African Try-

panosomiasis across the Democratic Republic of Congo. *PLOS Computational Biology*, 17:1–23, 01 2021. doi: 10.1371/journal.pcbi.1008532. URL https://doi.org/10.1371/journal.pcbi.1008532.

[16] OCHA Office for the Coordination of Humanitarian Affairs. *Journées Nationales de Vaccination (JNV) Activités de vaccination supplémentaire , RDC*, Accessed May 2016.

[24] World Health Organisation. Control and surveillance of human African trypanosomiasis. Technical Report 984, World Health Organisation, Nov. 2013.
